# Exploring parental prenatal influences on child health: a multicohort study and data visualisation tool

**DOI:** 10.1101/2025.08.17.25333431

**Authors:** Gemma C Sharp, Deborah A Lawlor, Kayleigh E Easey, Sampurna Kundu, Ahmed Elhakeem, Edward Hone, Rosemary R C McEachan, Maria C Magnus, Alexandra Havdahl, Caroline L Relton

**Affiliations:** School of Psychology, Faculty of Health and Life Sciences, University of Exeter, Exeter, UK; MRC Integrative Epidemiology Unit at the University of Bristol, Bristol, UK; Population Health Science, Bristol Medical School, University of Bristol, Bristol, UK; School of Psychological Science, University of Bristol, Bristol, UK; Research Software Engineers Group, University of Exeter, Exeter, UK; Bradford Institute for Health Research, Bradford Teaching Hospitals National Health Service Foundation Trust, Bradford, UK; Centre for Fertility and Health, Norwegian Institute of Public Health, Oslo, Norway; Nic Waals Institute, Lovisenberg Diaconal Hospital, Oslo, Norway; PsychGen Centre for Genetic Epidemiology and Mental Health, Division of Public Health and Prevention, Norwegian Institute of Public Health, Oslo, Norway; Center for Genetic Epidemiology and Mental Health, Norwegian Institute of Public Health, Oslo, Norway; PROMENTA Research Centre, Department of Psychology, University of Oslo, Oslo, Norway; London School of Hygiene and Tropical Medicine, London, UK

## Abstract

**Background:** The Developmental Origins of Health and Disease (DOHaD) hypothesis suggests that early life environmental exposures, especially during pregnancy, can impact long-term health. Research has largely relied on correlational evidence and has focused on maternal factors, with less attention given to paternal, postnatal, and broader social determinants. This focus could complicate efforts to determine the most effective strategies for improving population health.

**Methods:** Using harmonised data across four longitudinal cohort studies, we took a systematic approach to explore associations of parental prenatal health behaviours (smoking, alcohol, and caffeine consumption) and low socioeconomic position (SEP) with 72 child health-related outcomes (e.g. related to body size and composition, cognitive function, mental health, blood pressure, allergy, etc). Where possible, cohort estimates were meta-analysed, yielding a maximum sample size of over 230,000. We triangulated evidence of causality using different analytical approaches, including Mendelian randomization, negative controls (maternal-vs-paternal and during-vs-post-pregnancy comparisons), and dose-response analyses. This comprehensive set of analyses generated more than 594,000 effect estimates. We developed a web app, ‘EPoCH Explorer’ to visualise and share our results in an accessible format.

**Results:** We did not find strong evidence for widespread or large effects of parental health behaviours on child health and wellbeing. Only 6% of analyses had a Cohen’s D value >0.2 and FDR-adjusted P<0.05. In most analyses, the effect estimate was similar for mothers and partners, with 51% showing a larger effect for mothers and 49% for partners. Despite the lack of widespread associations, we found consistent evidence of association between maternal smoking and small for gestational age, higher childhood body mass index (BMI), depressive symptoms, and behavioural issues, while partner smoking was consistently associated with childhood BMI and social communication difficulties. Overall, we found stronger evidence of child outcomes being associated with low SEP than with health behaviours: 15% of results for low SEP had a Cohen’s D value >0.2 and FDR-P <0.05, compared to 6% for parental smoking, 3% for alcohol, and 0.4% for caffeine.

**Conclusion:** Our findings suggest that wider familial socioeconomic conditions may be a more important determinant of child health than specific parental health behaviours prenatally. Interventions to improve population health may be most effective if they target wider social inequalities. We encourage researchers to use EPoCH Explorer to prioritise associations for exploration in their own datasets, thus enabling replication and cross-context comparison to validate and extend the generalisability of our findings.

## Introduction

The Developmental Origins of Health and Disease (DOHaD) hypothesis posits that environmental exposures in early life, including those that occur prenatally, can affect long-term health (1). Traditionally, the field has focused on aspects of the maternal environment in and around pregnancy, including how a mother’s health behaviours might affect her fetus *in utero* (i.e. “maternal pregnancy effects”) (2). There has been much less of a focus on other types of exposure, including postnatal factors, paternal factors, and the wider social determinants of health (2–4). This makes it difficult to test (rather than merely reinforce) the DOHaD hypothesis, and to challenge implicit and deeply-held assumptions about the causal primacy of maternal pregnancy effects (2). It also restricts our ability to contextualise the evidence on maternal pregnancy effects to understand whether efforts to improve offspring health would be most effective if they were directed at mothers only or towards partners too, or beyond individuals – targeting wider social determinants. In addition, current evidence used to support guidelines about changes to behaviour around pregnancy is often based on correlational rather than causal evidence (3,5,6). Although the field necessarily relies largely on observational data, few studies have employed techniques that attempt to infer causal effects from such data. Even fewer have triangulated (7) evidence from multiple causal inference techniques to try to balance competing assumptions and improve confidence in causal conclusions. Finally, although copious prenatal (mostly maternal) exposure and offspring outcome combinations have been explored in the literature, sample sizes have often been small and differences in study design, populations, and definitions of variables can make direct comparisons and meta-analysis difficult (8–10).

The Exploring Prenatal influences on Childhood Health (EPoCH) study attempted to address these challenges. We took a systematic approach to explore associations between prenatal health behaviours (smoking, alcohol, and caffeine consumption) in both mothers and their partners, and multiple health outcomes of their children, across four longitudinal cohort studies with a combined maximum sample size of over 230,000. To enable exploration of causality, we also applied the principles of Mendelian Randomization (MR) by calculating associations between child health outcomes and parents’ genetic risk scores for each health behaviour (11), explored time- and dose-specific effects (12), and considered the equivalent exposure in the other parent, or outside of pregnancy, as a negative control (13). Each of these causal inference methods has its own sources of bias, but the biases differ across approaches. To enhance confidence in our causal inferences, we undertook triangulation (7) to compare evidence from multiple methods wherever possible, reducing the impact of any single source of bias on our conclusions.

To help contextualise evidence for effects of the health behaviours of individual parents against effects of wider social influences on health, we also explored parental socioeconomic position (SEP) as an exposure. We meta-analysed results from individual cohorts where possible, yielding results from over 594,000 analyses involving parental exposures at up to nine timepoints, 72 outcomes over four age ranges, and eight adjustment sets. We have made these data available to the research community through EPoCH Explorer, a web-app providing interactive and customisable data visualisations, interpretations, and downloads.

In this paper, we describe some high-level insights regarding the relative strength of evidence for effects of mother vs partners, and health behaviours vs socioeconomic factors. We illustrate how EPoCH Explorer can be used to visualise triangulated evidence and provide causal insights. For this illustration, we use the effect of maternal smoking on child externalising traits (i.e., disruptive, hyperactive, or aggressive behaviours (14)) as an example. Finally, we invite researchers to use EPoCH Explorer in their own studies, for example: to generate hypotheses for further detailed exploration of causality in their own datasets; to check for replication of their own findings in EPoCH cohorts; or to further increase sample size and generalisability through meta-analysis of EPoCH results with comparable results from their own datasets.

## Methods

### Cohorts

We used data from four cohorts: the Avon Longitudinal Study of Parents and Children (ALSPAC; United Kingdom, pregnancies in 1991-1992) (15,16), Born in Bradford (BiB; United Kingdom, pregnancies in 2007-2011) (17,18), Millennium Cohort Study (MCS; United Kingdom, pregnancies in 2000-2001) (19), and the Mother, Father, and Child Cohort Study (MoBa; Norway, pregnancies in 1999-2008) (20). These cohorts were chosen because they collected data on most of the exposures/outcomes of interest, and had multiple follow-ups during childhood. More information about each cohort is provided in **Supplementary File 1**.

### Participants

Within each cohort, we excluded any children from multiple pregnancies (see **Supplementary File 1** for sample sizes before and after exclusions). No exclusions were made based on any other variable, but all our analyses were complete case, meaning that parent-child pairs were excluded from analyses if they did not have complete data on the exposure, outcome, and covariates for any specific exposure-outcome analysis. Therefore, the sample size varies between different exposure-outcome associations. For all cohorts, partners were invited by the enrolled study mother. This person cannot always be assumed to be the biological father, and paternity information was not collected by every cohort. Therefore, we use the term ‘partner’ rather than ‘father’.

### Parental exposures

To maximise the relevance to public health policy, we focused on parental health behaviours that are potentially modifiable during pregnancy: tobacco smoking, alcohol consumption, and caffeine consumption. Where data were available (summarised in **Supplementary File 2**), we derived each exposure variable for both mothers and partners. For each health behaviour, we derived several variables based on the nature and timing of exposure, with definitions harmonised across the four cohorts. These were: active smoking, passive smoke exposure, alcohol consumption, episodes of binge drinking, caffeine from tea, coffee, cola, and any of those sources. We considered up to eight timepoints: preconception (in the three to six months before pregnancy), separately in each trimester of pregnancy, in the two years postnatally, at any time during pregnancy, and for active smoking only, early onset (started smoking before age 11) and ever in life. Where possible, we derived ordinal variables with four levels of exposure: heavy, moderate, light and none (for definitions, see **Supplementary File 1**). All other variables were binary (any vs no exposure), or for caffeine consumption only, continuous (mg/day).

In addition to health behaviours, we also explored parental SEP as an exposure class, indicative of the wider social determinants of health. We defined separate binary measures of SEP based on highest education level achieved and current occupational social class where available. These variables compared the group with the lowest SEP (cohort specific – see **Supplementary File 1**) to all other groups as the reference.

### Genetic risk scores for parental exposures

We had access to genetic data for mothers in ALSPAC, BiB and MoBa, and partners (fathers) in MoBa. We calculated genetic risk scores (GRS) for parents’ smoking, alcohol, and coffee consumption for use in a Mendelian randomization framework to explore unconfounded potential causal effects of each health behaviour (21). GRS were calculated using PLINK V1.9, using SNPs identified in large scale GWAS of number of aggregated alcoholic drinks consumed per week (22), number of cups of coffee consumed per day (23), and four smoking-related traits: age of initiation or regular smoking, ever vs never been a regular smoker, cigarettes per day, smoking cessation (current vs former smoker) (22). Scores were weighted by the effect estimates reported in the corresponding GWAS and converted to standard deviation units (z scores) to facilitate comparing effects across different scores. The F-statistics for associations between GRS instruments and relevant exposures are shown in **Supplementary File 1**.

### Child outcomes and age stages

Where data were available (**Supplementary File 2**), we explored a broad range of child outcomes over five health domains: body size and adiposity; psychosocial and cognitive; immunological; serum biomarkers; and blood pressure. Outcomes were measured at up to six different timepoints depending on the outcome and data availability in each cohort: at delivery, first year, age 1-2 years, age 3-4 years, age 5-7 years, age 8-11 years. In addition, for some outcomes (e.g. asthma) it was appropriate to develop a variable to describe the outcome occurring or being diagnosed “any time in childhood” (up to age 11). All continuous outcomes were converted to standard deviation units (z scores) so that they could be meta-analysed across different cohorts and visualised on the same scale. To facilitate interpretation of estimates, we also generated binary variables for birthweight, BMI and some psychosocial and cognitive outcomes, based on commonly used clinical thresholds. Outcome variables are described in detail in **Supplementary File 1**.

### Covariates

The definition and harmonisation method for all covariates is described in detail in **Supplementary File 1**.

Where data were available, we derived harmonised variables for potential confounders of the relationship between parental health behaviours and child health (based on the established definition that confounders must be known or plausible causes of both the exposure and outcome): parents’ ethnicity (Asian, Black, white, mixed or other); parents’ age at conception or delivery (years); parents’ highest education (ordered categories, as defined in each cohort); parents’ occupational social class (ordered categories, as defined in each cohort); mother’s parity (nulliparous vs parity >=1).

We also generated harmonised variables for potential mediators (i.e. variables that potentially sit on the causal pathway between an exposure and an outcome) to include in a sensitivity analysis for some exposure-outcome combinations: gestational age at delivery (weeks); birthweight (kg); child’s caffeine consumption before age 2 (where available); child’s exposure to passive smoke before age 2 (where available).

Finally, all models were adjusted for sex (male, female) and age of the child at outcome measurement (years) to increase the precision of the estimate.

### Statistical analysis in each cohort

We conducted linear (for continuous outcomes) and logistic (for binary outcomes) multivariable regression models for each possible exposure/outcome combination, running up to eight models per exposure/outcome combination. These models and their rationales are outlined in **Table 1;** further details and the directed acyclic graphs (DAGs) are provided in **Supplementary File 3**. Where a cohort did not collect data on a particular covariate, it was omitted from the adjustment set for that cohort.

**Table 1.** Regression models that were run for each cohort.

| **Model number and name** | **Rationale** | **Adjustment set where the exposure is a health behaviour (i.e. smoking, alcohol, or caffeine)** | **Adjustment set where the exposure is socioeconomic position** | **Adjustment set where the exposure is a genetic risk score (GRS)** |
| --- | --- | --- | --- | --- |
| Model 1a: minimal – unadjusted for co-parent | Explores associations with maximum power and minimum selection, while still controlling for important covariates. | Child's sex, child's age at outcome measurement*, parent-of-interest's ethnicity | Child's sex, child's age at outcome measurement*, parent-of-interest's ethnicity | N/A |
| Model 1b: minimal – adjusted for co-parent | Additionally controls for the co-parent's exposure | Model 1a + *co-parent's exposure* | Model 1a + *co-parent's exposure* | N/A |
| Model 2a: standard – unadjusted for co-parent | Controls for key potential confounders | Child's sex, child's age at outcome measurement*, parent-of-interest's ethnicity, *age at conception/delivery*, *parity*<sup>†</sup>, *education*, *occupation*, *smoking/alcohol/caffeine*<sup>§</sup> | Child's sex, child's age at outcome measurement*, parent-of-interest's ethnicity, *age at conception/delivery*, *parity*<sup>†</sup> | Child's sex, child's age at outcome measurement*, parent-of-interest's age at DNA collection, 10 PCs |
| Model 2b: standard – adjusted for co-parent | Additionally controls for the co-parent's exposure | Model 2a + *co-parent's exposure* | Model 2a + *co-parent's exposure* | Model 2a + *co-parent's GRS* |
| Model 3a: standard with time-specific exposure – unadjusted for co-parent | Model 3 is only run when the exposure was measured during or post-pregnancy at repeat timepoints (see **Supplementary File 3**). It controls for correlation with previous timepoints to achieve a more time-specific estimate | Child's sex, child's age at outcome measurement*, parent-of-interest's ethnicity, *age*, *parity*<sup>†</sup>, *smoking/alcohol/caffeine*<sup>§</sup>, *the main exposure in previous timepoints* | N/A | N/A |
| Model 3b: standard with time-specific exposure – adjusted for | Additionally controls for the co-parent's exposure | Model 3a + *co-parent's exposure* | N/A | N/A |

| co-parent |  |  |  |  |
| --- | --- | --- | --- | --- |
| Model 4a: standard – unadjusted for co-parent & adjusted for potential mediators | Explores the direct effect of the exposure on the outcome by controlling for factors that (most likely) occur after the exposure and could sit on the causal pathway to the outcome | Child's sex, child's age at outcome measurement*, parent-of-interest's ethnicity, age, parity<sup>†</sup>, smoking/alcohol/caffeine<sup>§</sup>, *gestational age at delivery, birthweight, child's passive smoking, child's caffeine consumption* | Child's sex, child's age at outcome measurement *, parent-of-interest's ethnicity, age, parity<sup>†</sup>, *gestational age at delivery, birthweight, child's passive smoking, child's caffeine consumption* | Child's sex, child's age at outcome measurement*, parent-of-interest's age at DNA collection, 10 PCs, *child's GRS* |
| Model 4b: standard – adjusted for co-parent & adjusted for potential mediators | Additionally controls for the co-parent's exposure | Model 4a + *co-parent's exposure* | Model 4a + *co-parent's exposure* | Model 4a + *co-parent's GRS* |

*\*child's age was only included where there was variation in age within an outcome timepoint; <sup>†</sup>parity was only included where the parent-of-interest was the mother; <sup>§</sup>the two health behaviours that were not the main exposure were included; PCs: ancestry-informed principal components derived from genetic data. Italic text denotes variables that are new compared to the previous model.*

The minimally-adjusted models (models 1a/1b) included parent-of-interest’s ethnicity because this was a particularly important source of variation in health behaviours, especially for Born in Bradford, which had high ethnic heterogeneity. We considered the standard set of confounder adjusted models (models 2a/2b and 3a/3b) to be our main models.

The mediator-adjusted models 4a and 4b were run as a sensitivity analysis and results should be interpreted with caution because adjusting for mediators on the causal pathway can introduce collider bias (24). Mediators were included only where the outcome was measured *after* the potential mediator (e.g., child’s exposure to passive smoke postnatally was not included in models where the outcome was birthweight).

To allow exploration of sex-specific effects (which are supported by some DOHaD studies (25)), all models were additionally run stratified by sex of the child. We did not compare results between the sexes and the results of these models are not discussed further in this paper, but full results are available for others to explore and download via EPoCH Explorer.

### Processing of cohort results

To uphold the privacy of participants we removed results from analyses where the total sample size was <20, or any binary exposure or outcome group had n<5. We also removed results with an implausible effect estimate, which likely arose due to the model failing to converge (see **Supplementary File 3**).

### Meta-analysis

Where more than one cohort contributed results to an exposure/outcome combination, we estimated the weighted average effect across cohorts by inverse-variance weighted meta-analysis using the metafor package in R. Fixed-effects meta-analysis was used, because this is an appropriate method when there are a small number of studies (26). Heterogeneity was assessed using I^2^ and the heterogeneity P-value, and visually via forest plots.

### Processing of final results

We generated results from 594,326 analyses, which included a mix of regression results from single cohorts (where only one cohort had the required data for that analysis) and meta-analysis results (where two or more cohorts had the required data). P-values for each model were then corrected for multiple testing using a false discovery rate (FDR) of 5%. To aid visual comparisons between binary and continuous outcome results, we converted the log odds ratios for binary outcomes to Cohen’s D (27) (standardised mean difference) which is a unitless measure of effect (equation provided in **Supplementary File 3**). Effect estimates are presented in their raw format (i.e. not as Cohen’s D) in the full set of results.

### Causal inference approaches and triangulation

Our analyses allowed us to apply multiple causal inference approaches to strengthen causal interpretation of associations: 1) multivariable regression adjusting for potential confounders; 2) applying the principles of MR by instrumenting parental health behaviours with GRS (11); 3) employing co-parent’s exposure as a negative control to account for shared familial and environmental confounding (13); 4) using postnatal exposure as a negative control for pregnancy exposure (13); 5) exploring dose-response relationships where possible (12). Detailed methodology, including the assumptions and source of bias for each of these approaches, is outlined in **Supplementary File 3**. Causal evidence was evaluated based on predefined criteria (Table 2). Triangulation (7) was conducted for each exposure-outcome combination by summarising the number of analyses that provided causal evidence. When the mother was the parent-of-interest, strongest causal evidence was defined as support from five agreeing lines of evidence. When the partner was the parent-of-interest, we excluded the parental comparison negative control from this count – since the underlying assumptions are less likely to hold in this context (see discussion in Supplementary File 3) – and therefore defined strongest causal evidence as support from four agreeing lines of evidence.

**Table 2.** Causal inference approaches and criteria for evidence of a causal effect during pregnancy.

| Method | Exposure or exposure comparisons | Model or model comparisons | Criteria for evidence of a causal effect |
| --- | --- | --- | --- |
| Multivariable regression (MVR) | Health behaviour* ever in pregnancy | Model 2b | FDR-adjusted $P < 0.05$ |
| Mendelian randomization (MR) | Genetic risk score for health behaviour<sup>†</sup> | Model 2a (see Supplementary File 3 for justification) | $P < 0.05$ and directionally consistent with the equivalent observational effect identified in the MVR analysis |
| Parental comparison negative control<br>*(assumptions are more likely to hold when the mother is the parent-of-interest)* | Health behaviour ever in pregnancy | Model 2b for parent-of-interest vs equivalent model 2b for co-parent | $P < 0.05$ and directionally consistent after adjustment for the co-parent's exposure, with a greater absolute effect estimate for the parent-of-interest than the co-parent |
| Postnatal exposure negative control | Health behaviour ever in pregnancy vs health behaviour in the first two postnatal years | Model 2b for pregnancy exposure vs model 3b for postnatal exposure | A greater absolute effect estimate for the exposure during pregnancy than the exposure postnatally (with the postnatal estimate having been adjusted for the exposure during pregnancy) |
| Dose response | Heavy levels of the health behaviour during pregnancy vs light levels of the health behaviour during pregnancy | Model 2b | Any exposure timepoint during pregnancy (i.e. first, second, or third trimester) with an effect estimate that is greater for heavy compared to light doses of the exposure |

*\* We select the 'main' health behaviour definition for these analyses, i.e. active smoking, any alcohol consumption, and caffeine consumption from any source (rather than passive smoking, binge drinking, or caffeine from tea/coffee/cola). <sup>†</sup> For smoking, which has multiple GRSs, we use the GRS for smoking initiation because it has the strongest association with the phenotype.*

### Comparison to low SEP

For each outcome, we compared the estimated effect of each health behaviour in pregnancy (ever vs never) to the estimated effect of either measure of low SEP (i.e., based on education or occupation; lowest group vs higher groups). For these analyses, we used model 2a (i.e. the standard adjustment set) to ensure that the health behaviour model was adjusted for SEP while maximising power and minimising selection by not adjusting for the co-parent’s health behaviour. If the association between low SEP and the outcome is similar to, or greater than, the association between the health behaviour and the outcome, then the effect of the health behaviour might reflect residual confounding by SEP. In this case, the health behaviour may not be a specific or strong causal factor but rather part of a broader pattern of disadvantage associated with low SEP.

### EPoCH Explorer

To share, visualise, and draw inferences from our results, we developed EPoCH Explorer, an RShiny web app, which can be accessed at https://gcsharp.shinyapps.io/EPoCH/. Via EPoCH Explorer, users can view cohort data summaries and generate data visualisations of regression and meta-analysis summary statistics. Verbal interpretations of results are provided, as well as a causal inference report that presents all information needed to triangulate and contextualise the causal evidence for any exposure specified by the user. EPoCH Explorer also allows users to download the full set of summary statistics for EPoCH.

### Reproducibility

All analysis code is available online: https://github.com/gcsharp/EPoCH_analysis and has been developed iteratively and checked (see Supplementary File 3) to improve reproducibility. Access to individual level data is governed by each cohort, but all summary statistics are available via EPoCH Explorer https://gcsharp.shinyapps.io/EPoCH/. This paper is written in accordance with the STROBE guidelines for reporting of observational studies (checklist in **Supplementary File 4**).

## Results

### Sample sizes

Without stratification by sex, sample sizes ranged from 35 to 232,139 (mean = 18,554) and varied according to the model, the availability of data across cohorts, and missing values for participants within cohorts. For our standard models 2a and 2b (unstratified by sex, adjusted for the standard set of covariates), the number of analyses involving more than one cohort was 8955/29146 (31%) for model 2a and 7760/28171 (28%) for model 2b. See **Supplementary File 5** for a detailed summary of the number of cohorts and sample sizes for each model. The maximum sample size decreased by up to 22% after adjustment for a standard set of confounders (i.e. from model 1-2), but only up to 3.1% after further adjustment for potential mediators (i.e. from model 2 to model 4). It is not possible to make a direct comparison with model 3 because this model was only run for health behaviours in the second trimester, third trimester, or postnatally, and total sample sizes were lower for these exposure variables.

### Sample characteristics

Figure 1 summarises the percentage of the sample (pooled across cohorts where multiple cohorts contributed) that were in the exposed group for each binary exposure variable. Mothers self-reported lower levels of smoking and alcohol consumption than partners did, particularly during pregnancy. For caffeine consumption, differences between parents and pregnancy status were less apparent. Less than 23% of parents were categorised in the lowest SEP groups. In all cohorts, low SEP was correlated with high smoking (and caffeine consumption except in MoBa). In all cohorts except BiB, low SEP was weakly correlated with low alcohol consumption (**Supplementary File 5**). In general, cohorts had prioritised collecting data on mothers, so more variables were available (**Supplementary File 2**), and the data were more complete compared to data on partners. **Supplementary File 6** contains a detailed breakdown of sample characteristics by cohort.

**Figure 1.**
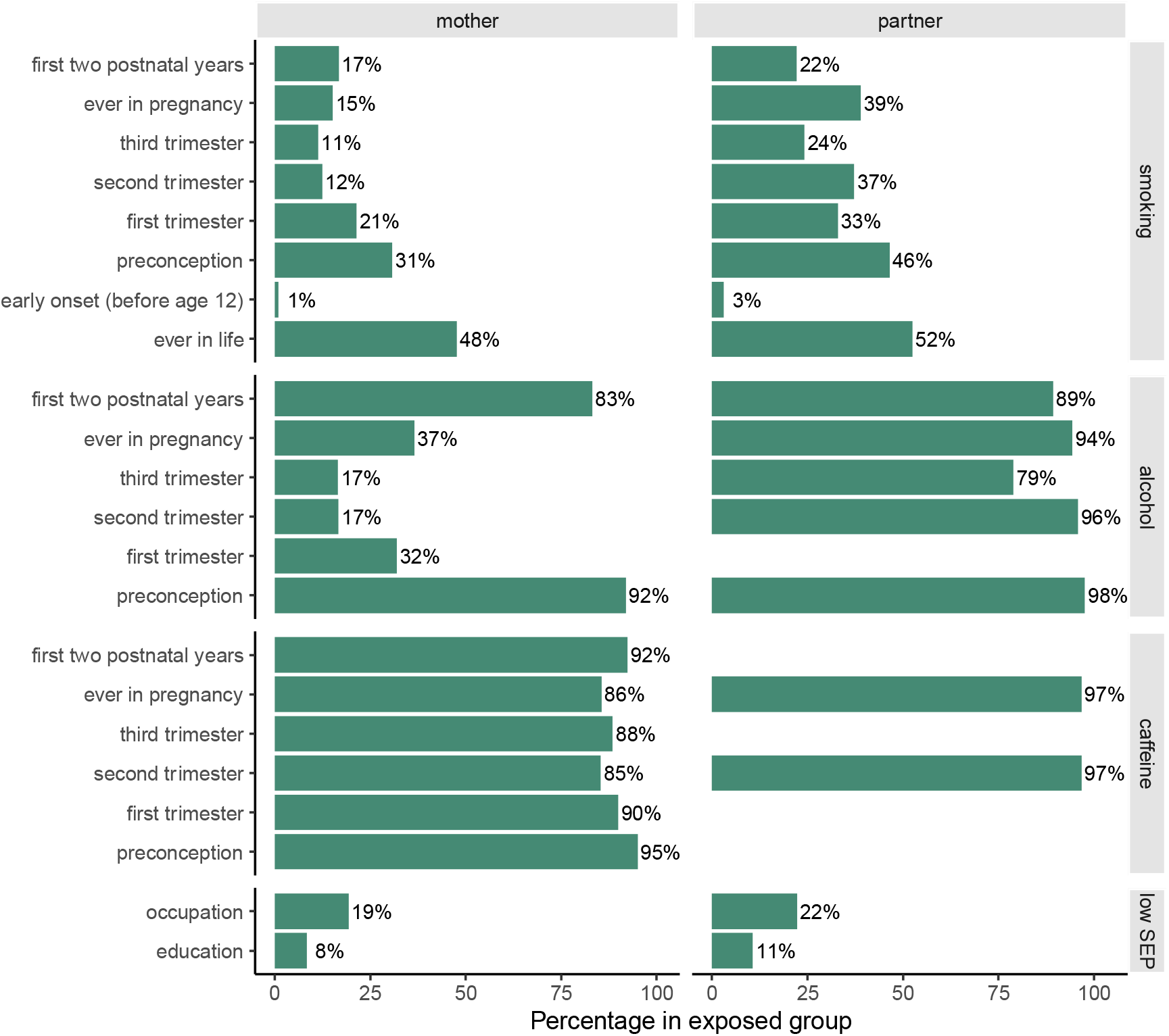
Summary of parental exposures in EPoCH. Percentages are the exposed sample pooled across all contributing cohorts, divided by the total non-missing sample pooled across those cohorts. SEP = socioeconomic position.

### General trends

Across all models (i.e. models 1a to 4b across all health behaviours, GRS, SEP, and outcomes), effect estimates were generally small (absolute Cohen’s D<0.2) and P-values large (FDR-adjusted P >0.05). In our standard models, the percentage of results with D>0.2 and FDR-P<0.05 (i.e. strong statistical evidence of a larger-than-small associations) was 6% before adjustment for the co-parent’s exposure (model 2a), and 3% after (model 2b).

We did not find widespread evidence of much stronger effects in mothers than partners: after mutual adjustment (model 2b), 51% of analyses had a larger effect estimate for mothers than partners, and 52% had a smaller p-value for mothers than partners. The percentage of model 2b results with D>0.2 and FDR-P<0.05 was 7% for mothers, compared to 5% for partners.

We observed some variation between exposure and outcome classes (**Figure 2** and **Supplementary File 5**). Across all outcomes, the strongest associations (D>0.2 and FDR-P<0.05) were seen with low SEP. In model 2b (standard adjustment with adjustment for the exposure in the co-parent), 15% of the results for low SEP had D>0.2 and FDR-P<0.05, compared to 6% for smoking, 3% for alcohol, and 0.4% for caffeine. Across all exposures, strongest associations were seen with psychosocial and cognitive outcomes (especially when the exposure was low SEP, smoking, or partner’s alcohol consumption) and body size and composition (especially when the exposure was smoking). For smoking, caffeine, and SEP, we saw a consistent pattern: a slightly higher proportion of strong associations for mothers compared to partners. In contrast, for alcohol, the proportion of strong associations was higher for partners. Notably, the direction of the partner association was often opposite to that seen for mothers (particularly for psychosocial outcomes), suggesting a potential protective effect of partner’s alcohol consumption during pregnancy. However, associations with the partner alcohol GRS (**Supplementary File 5**; F-statistic for instrument strength = 10 (**Supplementary File 1**)) were mostly null, indicating that this apparent protective effect could be explained by residual or unmeasured confounding. We found no clear patterns according to the timing of exposure or age at which outcomes were assessed (**Supplementary File 5**).

**Figure 2.**
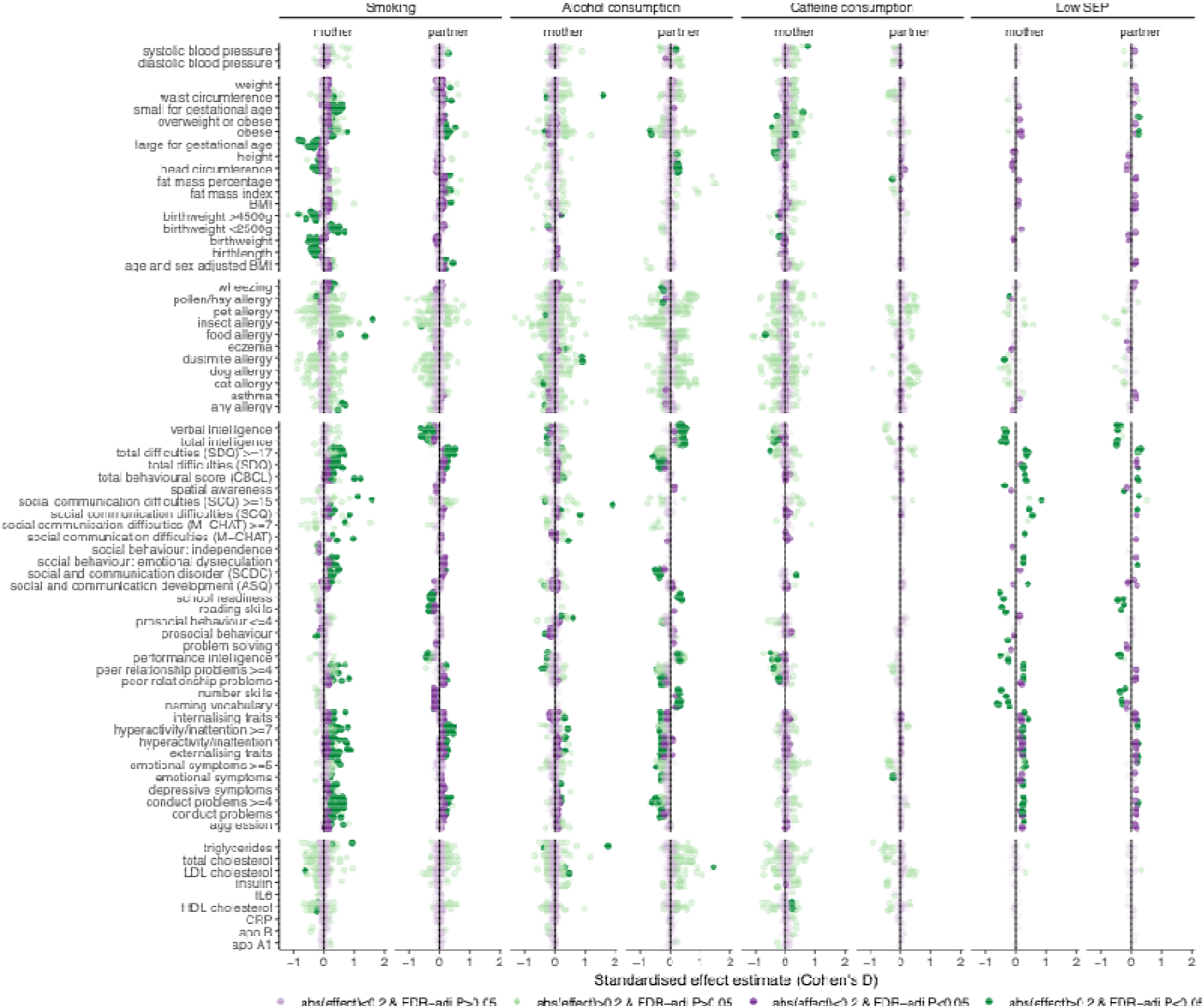
Summary of results by exposure and outcome class for the standard model with adjustment for co-parent’s exposure (model 2b). Dark green indicates the strongest evidence (FDR<0.05) of a larger-than-small effect (D>0.2).

### Triangulation of evidence for causal effects in pregnancy

In model 2b (standard adjustment with adjustment for the exposure in the co-parent), the number of analyses where the exposure was a health behaviour that occurred *during* pregnancy was 10,564 for mothers and 6049 for partners. Of these, 1099 (10%) and 585 (9%), respectively, had FDR-P<0.05. **Figure 3** illustrates triangulation of causal evidence for each of these associations. Note that a lack of evidence does not indicate evidence *against* a causal effect during pregnancy; it may reflect limited data availability or differences in assumptions across contexts. For most exposure-outcome combinations, the direction of association is consistent between the equivalent MVR and MR analyses, but there are instances for caffeine consumption where this is not true, indicating potential residual confounding in the observational (MVR) estimates (or bias caused by relatively weak instruments – see **Supplementary File 1** for F-statistics).

**Figure 3.**
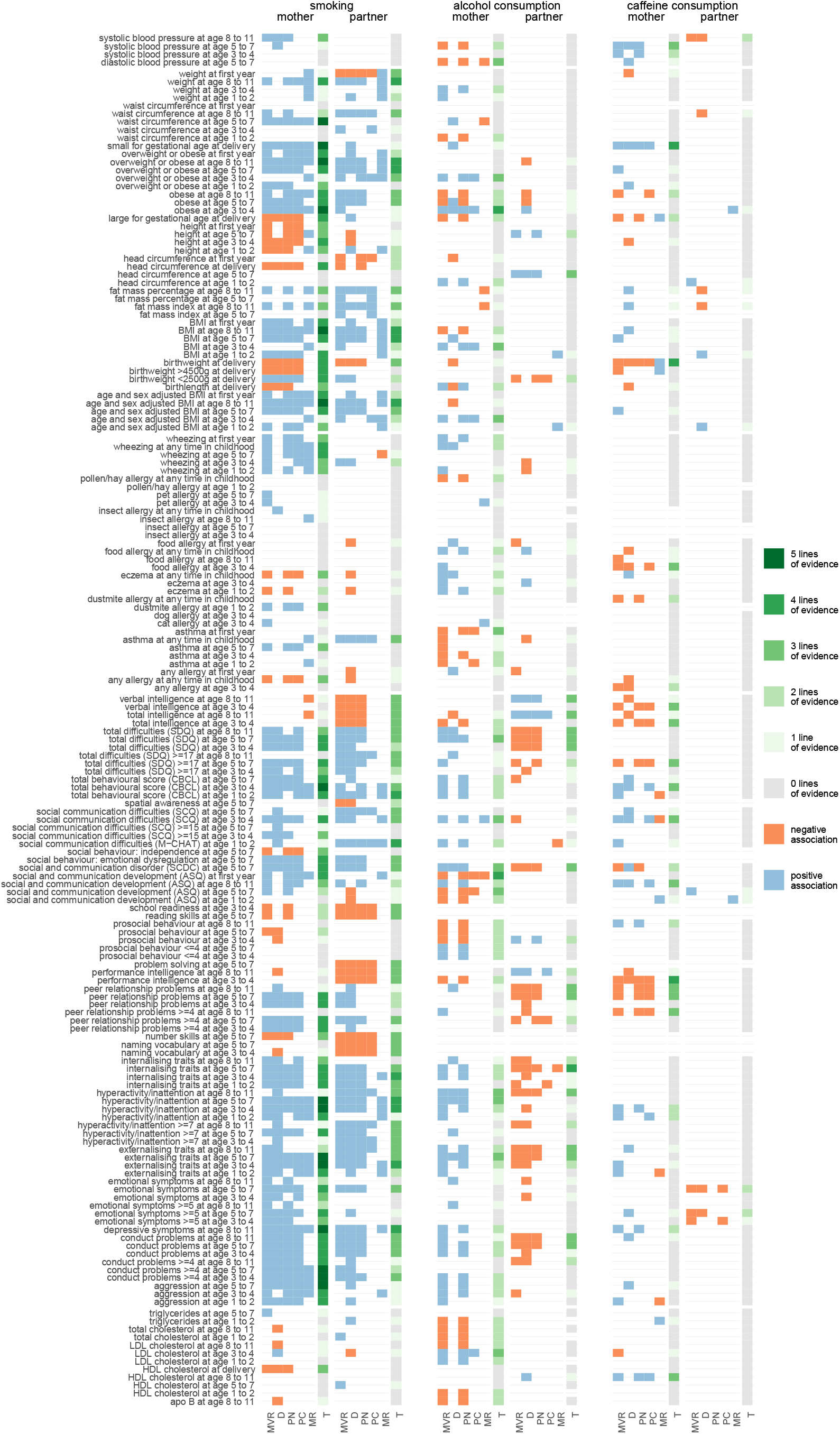
Triangulation of evidence for effects of parental health behaviours during pregnancy. MVR = multivariable regression (model 2b); PC = parental comparison negative control (model 2b); MR = mendelian randomisation genetic risk score (model 2a); D = dose response (model 2b); PN = postnatal comparison negative control (model 3b); T = triangulation summary indicating the number of lines of evidence across different analytical approaches that agree on the direction of association and have P<0.05.

For mothers, we found five agreeing lines of evidence across analytical approaches for an effect of smoking during pregnancy on: small for gestational age (SGA) at delivery; waist circumference at age 5-7, BMI and overweight/obese at age 8-11; conduct problems, externalising traits, and hyperactivity/inattention at ages 3-4 and 5-7; aggression at age 5-7; total behavioural issues at age 3-4; and depressive symptoms at age 8-11. It should be noted that, when adjusting for child’s and co-parent’s GRS (i.e. Model 4b) in the MR analysis (which can be an appropriate strategy to reduce genetic confounding and collider bias(28) – see Supplementary File 3 for a discussion), P-values were >0.05 for the body size and composition outcomes, possibly due to low statistical power. The body size and composition outcomes were all more strongly associated with maternal smoking than with low SEP (i.e., model 2a MVR effect estimates were further from the null). However, the psychosocial outcomes were more strongly associated with low SEP than maternal smoking.

For partners, we found four agreeing lines of evidence for an effect of smoking during pregnancy on BMI at age 5-7, BMI, overweight/obese, and weight at age 8-11; internalising traits at age 1-2 and 5-7; total behavioural issues at age 1-2, and social communication difficulties at age 1-2. We also found four agreeing lines of evidence for an effect of partner’s alcohol consumption during pregnancy on internalising traits. Additional adjustment for child’s and co-parent’s GRS (Model 4b) resulted in P-values >0.05 for the body size and composition outcomes and the associations with alcohol consumption. All identified outcomes were more strongly associated with low SEP than with partner health behaviours.

### Example of using EPoCH Explorer to triangulate causal evidence

Figure 4 shows screenshots from EPoCH Explorer to illustrate how a researcher might use the app to refine and assess evidence for (as an example) a hypothesis about prenatal influences on child psychosocial outcomes. To select an outcome of interest, the researcher could start by generating a Manhattan plot of the minimally-adjusted (model 1a) results for psychosocial traits, which would reveal that the strongest associations are between maternal smoking during pregnancy and child externalising traits at age 5-7. To explore this hypothesis further, they could generate a causal inference report (example in **Supplementary File 7**), which would show that: (1) the MVR association persists after adjusting for potential confounders; (2) the estimate for smoking initiation GRS aligns with the MVR estimate; (3) the maternal estimate is larger than the partner’s; (4) the estimate is larger for smoking during pregnancy than postnatally, (5) a dose-response relationship exists. These five lines of evidence support a causal effect of maternal smoking in pregnancy on externalising traits at age 5-7 years. The effect estimate for maternal smoking ever in pregnancy (model 2a β=0.21 (95% confidence interval 0.18 to 0.25, P=1.4×10^−31^) is smaller than the effect estimates for low SEP (education β=0.30 [95% CI=0.26 to 0.34, P=2×10^−46^], occupation β=0.26 [95% CI=0.21 to 0.31, P=1.5×10^−23^]), which suggests the effect may not be specific to smoking, but part of a broader pattern of disadvantage. The researcher could then perform a ‘Deep Dive’ to explore (for model 2b) cohort sample sizes, summary statistics, a forest plot, and a verbal interpretation of the findings. This shows that three cohorts (ALSPAC, MCS, MoBa) contributed to the meta-analysis, with a sample size of 40,014 (n smokers = 2971; 7%), and low heterogeneity (I^2^ = 23.4; heterogeneity P-value = 0.27). The researcher could also use EPoCH Explorer to investigate the impact of stratifying by sex, or adjusting for mediators (i.e. comparing to model 4). Finally, the researcher could download all results to analyse offline and/or to compare to their own data.

**Figure 4.**
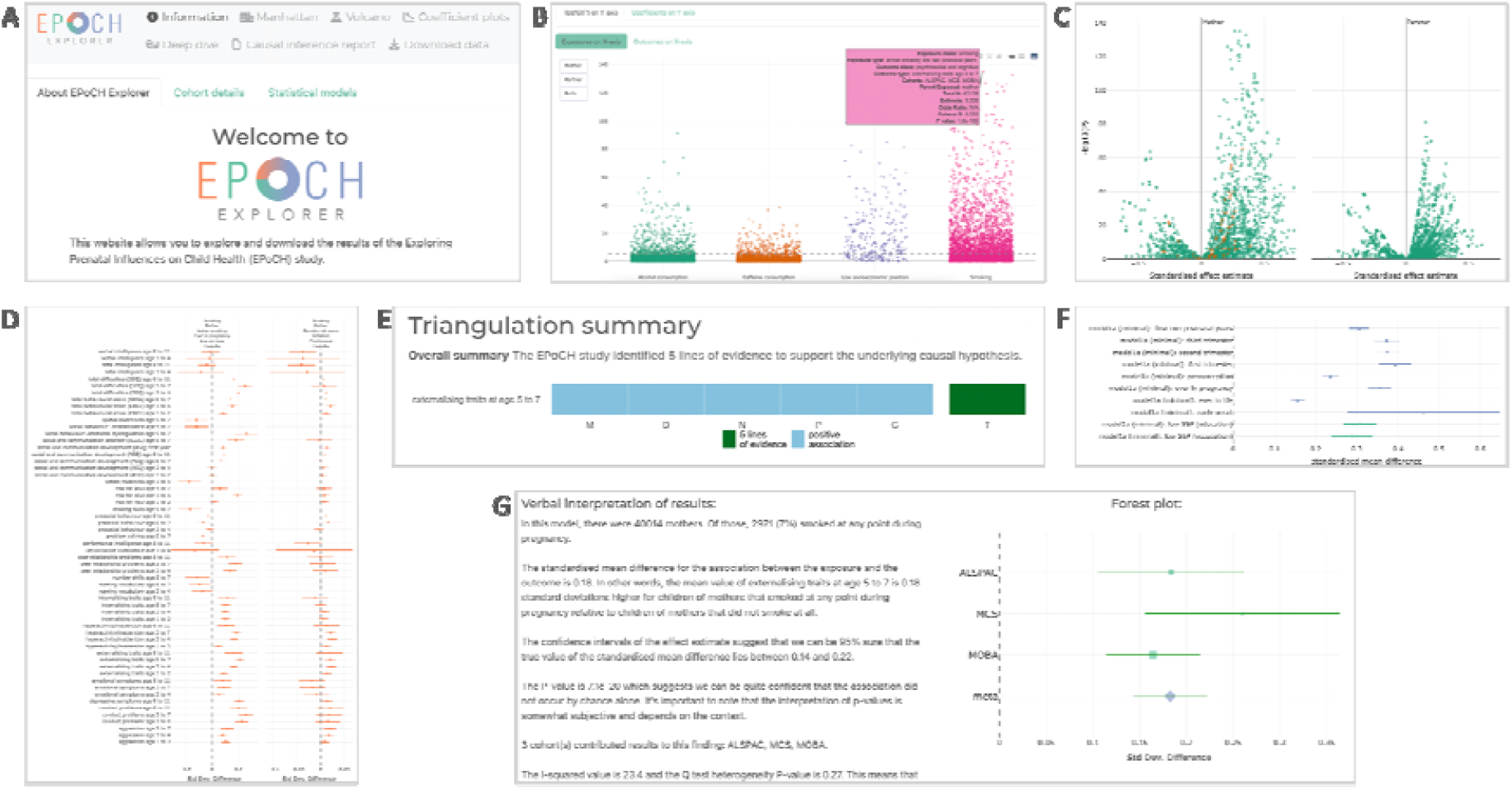
Screenshots from EPoCH Explorer. A. The navigation bar and top of the welcome page; B. An interactive Manhattan plot showing the distribution of P-values by exposure class for all psychosocial outcomes; C. An interactive volcano plot comparing maternal to partner estimates for smoking and psychosocial outcomes; D. Two coefficient plots comparing observational and genetic estimates for maternal smoking and continuous psychosocial outcomes; E. The triangulation summary taken from a causal inference report for maternal smoking and child externalising traits at age 5-7; F. A coefficient plot comparing estimates of smoking and low SEP on this outcome; G. A ‘Deep Dive’ summary of the results from model 2B, including a verbal interpretation of results and an interactive forest plot. EPoCH Explorer can be accessed at: https://gcsharp.shinyapps.io/EPoCH/

## Discussion

### Summary of results

In this systematic exploration – of four types of exposure in both parents at up to nine timepoints, on 69 child outcomes at up to four ages – we did not find strong evidence for widespread or large magnitude effects of parental health behaviours on child health and wellbeing. We did find consistent evidence for parental smoking on a small number of offspring outcomes, including maternal smoking increasing small for gestational age, partner smoking increasing social communication difficulties, and both parents increasing childhood BMI, depressive symptoms, and behavioural issues. In general, we found stronger evidence of association with low SEP compared to health behaviours, suggesting that wider familial socioeconomic conditions may be a more important determinant of child health than specific parental health behaviours prenatally.

Our findings support calls for DOHaD research to broaden the focus from maternal (mostly intrauterine) exposures to paternal and wider societal influences (2,3,29–31). This follows increasing evidence of paternal influences on child health, as outlined in recent systematic reviews in humans and/or animals (32–36). These reviews noted the high risk of bias in the included studies, including inadequate adjustment for maternal exposures (and therefore assortative mating (37)). We have previously shown that, mirroring what we see for humans, there is a paucity of animal research on paternal effects in the DOHaD literature (2).

The widespread association we identified between low SEP and poorer offspring health outcomes agrees with the large body of research underscoring the significant impact of socioeconomic factors on child health, including adiposity and behavioural difficulties (38– 40).

Our finding of higher partner alcohol consumption being associated with psychosocial and cognitive outcomes is in line with the well-established J-shape relationship between alcohol consumption and health, whereby light to moderate alcohol consumption is associated with better health outcomes due to socioeconomic confounding (41). Therefore, the positive associations between partner alcohol and child psychosocial outcomes (in our fully adjusted models, i.e. after adjustment for SEP) are likely to be explained by residual confounding by SEP. We hypothesise that the maternal estimates are not affected in the same way because mothers reduce their alcohol consumption during pregnancy to a much larger degree than partners. Indeed, before pregnancy, the trends for partners and mothers were more similar (**Supplementary File 5** Figure 2).

Our evidence of effects of maternal smoking on small for gestational age agrees with a large body of literature, including studies that have applied approaches to infer causality (42–44). The possible effect of maternal smoking on childhood BMI corroborates some but not all previous studies: despite strong evidence of association from multivariable regression analyses (10), studies that have used a parental negative control design (13) or Mendelian randomization (45) have yielded null results. The association between maternal smoking and depressive symptoms in children is less well characterised, but a study using a parental negative control design found little evidence for a causal effect on depressive symptoms or internalising traits (46), and another found that a more thorough adjustment for parental psychosocial factors attenuated the association with internalising traits to the null (47). Additionally, an MR study in UK Biobank (48), and a multicohort study using a negative control and cross-cohort design (49) have both found limited support for a causal effect on *adult* depression. Previous studies on maternal smoking and behavioural issues have shown mixed results. A systematic review (50) found that 13 out of 19 studies reported a positive association with ADHD, but six studies concluded the association was due to confounding. Large genetically informed studies, including a meta-analysis of over one million participants, provide no evidence for a causal association between maternal smoking during pregnancy and offspring ADHD (51). Evidence from sibling-comparison, parental-comparison, and polygenic score analyses suggests that observed associations are largely explained by genetic and familial confounding, and that paternal smoking may not be a valid negative control for assessing intrauterine effects of maternal smoking on ADHD and related traits (52). Finally, our finding of consistent evidence for an effect of partner smoking on social communication difficulties corroborates a previous study showing an association with autistic traits (53), but contrasts another showing no association (54). Neither study applied approaches to infer causality. Differences between our results and those of previous studies could be explained by differences in definition, measurement, and timing of exposures/outcomes, adjusted covariates, statistical power, and populations studied.

### Strengths

The EPoCH study takes a novel, ambitious, and comprehensive approach to systematically explore potential causal effects of maternal and partner prenatal exposures and child outcomes at different timepoints. The use of prospective longitudinal population-based data minimises recall bias and enables associations to be examined over time. By harmonising data across cohorts, we were able to maximise comparability. For some analyses, the sample size was very large supporting precise estimates.

We focus our analyses on potentially modifiable parental health behaviours, and importantly, provide a useful, free, open resource for the research community (the EPoCH Explorer web app). The interactive data visualisations enable researchers to explore our results, and generate and assess evidence for their own hypotheses. It allows users to download the full set of results for use in their own analyses, thereby facilitating further meta-analysis and comparisons to other populations. The causal inference report and ‘Deep Dive’ verbal interpretation of results includes explanations that make EPoCH Explorer a valuable resource for students of epidemiology and others interested in understanding the data.

### Limitations

Despite our attempt to harmonise variables across studies, differences in availability of data meant that up to two thirds of our analyses were conducted within a single cohort. Therefore, the sample size was relatively small for some analyses (range 35 to 256,387, mean = 17,326). The generalisability of our findings is also constrained by the low ethnic and socioeconomic diversity across the cohorts, except for BiB. It is unclear whether our findings apply to populations and settings outside of high-income countries in Northern Europe.

Some of the variation in sample size across analyses arises due to missing data within collected variables. Due to the number and complexity of our analyses, we chose to use a complete-case approach and did not employ multiple imputation techniques. The exclusion of participants with incomplete data may have introduced bias, particularly if the data are not missing at random. Additionally, as with all self-report data, there may be recall and measurement errors that could affect validity.

The inclusion of non-biological partners can be considered a strength (e.g., our approach is inclusive of same-sex parents, maximises sample size, and is beneficial for controlling for shared household confounding), but it may not fully describe the partner effect if the mechanism is hypothesised to operate via paternal genetics, sperm (*e*.*g*., *de novo* genetic mutations, sperm function, epigenetics, cytoplasmic factors) or other constituents of semen. However, we were limited by the lack of information about paternity status for some cohorts. The inclusion of non-biological partners could mean that mothers are more genetically related to study children than partners are. Although this might bias partner estimates (in either direction depending on exposure/outcome), sensitivity analyses have shown that the bias is minimal, even with implausibly high simulated rates of non-paternity (55). Similarly, higher rates of missing data for partners relative to mothers has the potential to introduce bias, particularly as we have previously shown that those partners who are recruited tend to be of a higher SEP than those who are not (56). However, our previous simulation study provides confidence that any resulting selection bias on the partner or partner-adjusted maternal estimate was likely to be small (56).

EPoCH focused on outcomes in childhood, and we did not examine miscarriage or still birth. Given that our exposures could increase risk of these outcomes, our analyses may be affected by live-birth bias (57), a type of selection bias induced by restricting to live births. This is a potential limitation of many birth and child cohort studies. Fetal loss is difficult to study in ALSPAC or BiB, where the low number of cases result in insufficient statistical power, and impossible to study in MCS, because only families with a child around nine months-old were recruited.

Finally, some GRSs were only weakly associated with the behaviour they were intended to instrument (see **Supplementary File 1** for F-statistics). Deriving our GRSs using the top SNPs from previous GWAS helps ensure specificity, but it is dependent on the power in the original GWAS. We used the largest GWASs available at the time of analysis for smoking, alcohol, and caffeine consumption to maximise power, but it is possible that using a wider subset of SNPs would have generated GRSs that captured more variance in these health behaviours (but with a likely trade off on specificity). Future research should conduct sensitivity analyses to test the assumptions of MR (58).

### Implications for future research and practice

Our findings suggest that there is no strong scientific rationale to focus exclusively on maternal influences, and future DOHaD studies should broaden their scope to study data on partners, postnatal exposures, and the wider social determinants of health. Partners are often overlooked in current preconception care policies and little guidance or supportive policies are available for them, or targeted at both parents together (59). Behaviours such as smoking, caffeine, and alcohol consumption are socially patterned and often occur alongside other factors that may elevate the risk of poor health and psychosocial wellbeing in children. Interventions to address social inequalities (for example, improving access to healthcare and ensuring good quality housing) are expected to improve childhood health outcomes (60–62).

## Conclusion

We found limited evidence for large effects of parental prenatal health behaviours on childhood health and wellbeing, and stronger evidence of association with low SEP. This suggests that wider familial socioeconomic conditions may be a more important determinant of child health than specific parental health behaviours prenatally.

Our findings highlight the need for better data and more research into paternal/partner and wider societal or structural determinants of health.

## Supporting information

Supplementary File S1: Cohort Information

Supplementary File S2: Data Availability

Supplementary File S3: Statistical analysis details

Supplementary File S4: STROBE checklist

Supplementary File S5: Supplementary Results

Supplementary File S6: Cohort Variable Distributions

Supplementary File S7: Example from EPoCH Explorer

## Data Availability

Individual level data from each cohort cannot be shared under the terms of each cohorts data sharing agreement (but is available to researchers directly from the cohorts themselves), but all summary statistics from our analyses are available via EPoCH Explorer: https://gcsharp.shinyapps.io/EPoCH/.

## Funding and acknowledgements

Thank you to all participants who contributed to the cohort studies. See **Supplementary File 1** for cohort-specific funding and acknowledgements. EPoCH was funded by a grant from the Medical Research Council to GCS (MR/S009310/2). DAL and AE are supported by the Medical Research Council Integrative Epidemiology Unit (MRC IEU) at the University of Bristol (MC UU 00032/05, MC UU 00032/06), and GCS, KE, and CR were also supported by the MRC IEU when part of this work was conducted. The funders had no role in the design or execution of the study. MCM is supported by the Research Council of Norway through its Centers of Excellence funding scheme (project No 262700). AH is supported by the Research Council of Norway (#336085), the South-Eastern Norway Regional Health Authority (#2020022, #2018059, #2922083), and the European Union’s Horizon Europe Research and Innovation programme (FAMILY; #101057529, and Marie Skłodowska-Curie grant ESSGN #101073237).

## References

1. Barker DJP. The origins of the developmental origins theory. J Intern Med. 2007 May;261(5):412–7.

2. Sharp GC, Schellhas L, Lawlor DA. Time to cut the cord: recognizing and addressing the imbalance of DOHaD research towards the study of maternal pregnancy exposures. J Dev Orig Health Dis. 2019 Oct 1;10(5):509–12.

3. Sharp GC, Lawlor DA, Richardson SS. It’s the mother!: How assumptions about the causal primacy of maternal effects influence research on the developmental origins of health and disease. Soc Sci Med. 2018 Sep;213:20–7.

4. Ben-Shlomo Y, Cooper R, Kuh D. The last two decades of life course epidemiology, and its relevance for research on ageing. Int J Epidemiol. 2016 Aug 1;45(4):973–88.

5. Gage SH, Munafò MR, Davey Smith G. Causal Inference in Developmental Origins of Health and Disease (DOHaD) Research. Annu Rev Psychol. 2016 Jan 4;67(1):567–85.

6. Elliott HR, Sharp GC, Relton CL, Lawlor DA. Epigenetics and gestational diabetes: a review of epigenetic epidemiology studies and their use to explore epigenetic mediation and improve prediction. Diabetologia. 2019 Dec 1;62(12):2171–8.

7. Lawlor DA, Tilling K, Davey Smith G. Triangulation in aetiological epidemiology. Int J Epidemiol. 2017 Jan 20;dyw314.

8. Mamluk L, Edwards HB, Savović J, Leach V, Jones T, Moore THM, et al. Low alcohol consumption and pregnancy and childhood outcomes: time to change guidelines indicating apparently ‘safe’ levels of alcohol during pregnancy? A systematic review and meta-analyses. BMJ Open. 2017 Jul;7(7):e015410.

9. Oken E, Levitan EB, Gillman MW. Maternal smoking during pregnancy and child overweight: systematic review and meta-analysis. Int J Obes. 2008 Feb;32(2):201–10.

10. Magalhães EIDS, Sousa BAD, Lima NP, Horta BL. Maternal smoking during pregnancy and offspring body mass index and overweight: a systematic review and meta-analysis. Cad Saúde Pública. 2019;35(12):e00176118.

11. Burgess S, Thompson SG. Use of allele scores as instrumental variables for Mendelian randomization. Int J Epidemiol. 2013 Aug;42(4):1134–44.

12. Hill AB. The environment and disease: association or causation. Proc R Soc Med. 1965 May;58(5):295–300.

13. Smith GD. Negative Control Exposures in Epidemiologic Studies. Epidemiology. 2012 Mar;23(2):350–1.

14. Liu J. Childhood Externalizing Behavior: Theory and Implications. J Child Adolesc Psychiatr Nurs. 2004 Jul;17(3):93–103.

15. Boyd A, Golding J, Macleod J, Lawlor DA, Fraser A, Henderson J, et al. Cohort Profile: The ‘Children of the 90s’—the index offspring of the Avon Longitudinal Study of Parents and Children. Int J Epidemiol. 2013 Feb;42(1):111–27.

16. Fraser A, Macdonald-Wallis C, Tilling K, Boyd A, Golding J, Davey Smith G, et al. Cohort Profile: The Avon Longitudinal Study of Parents and Children: ALSPAC mothers cohort. Int J Epidemiol. 2013 Feb 1;42(1):97–110.

17. Wright J, Small N, Raynor P, Tuffnell D, Bhopal R, Cameron N, et al. Cohort Profile: The Born in Bradford multi-ethnic family cohort study. Int J Epidemiol. 2013 Aug 1;42(4):978– 91.

18. McEachan RRC, Santorelli G, Watmuff A, Mason D, Barber SE, Bingham DD, et al. Cohort Profile Update: Born in Bradford. Int J Epidemiol. 2024 Feb 14;53(2):dyae037.

19. Connelly R, Platt L. Cohort Profile: UK Millennium Cohort Study (MCS). Int J Epidemiol. 2014 Dec 1;43(6):1719–25.

20. Magnus P, Birke C, Vejrup K, Haugan A, Alsaker E, Daltveit AK, et al. Cohort Profile Update: The Norwegian Mother and Child Cohort Study (MoBa). Int J Epidemiol. 2016 Apr;45(2):382–8.

21. Davey Smith G, Hemani G. Mendelian randomization: genetic anchors for causal inference in epidemiological studies. Hum Mol Genet. 2014 Sep 15;23(R1):R89–98.

22. Liu M, 23andMe Research Team, HUNT All-In Psychiatry, Jiang Y, Wedow R, Li Y, et al. Association studies of up to 1.2 million individuals yield new insights into the genetic etiology of tobacco and alcohol use. Nat Genet. 2019 Feb;51(2):237–44.

23. The Coffee and Caffeine Genetics Consortium, Cornelis MC, Byrne EM, Esko T, Nalls MA, Ganna A, et al. Genome-wide meta-analysis identifies six novel loci associated with habitual coffee consumption. Mol Psychiatry. 2015 May;20(5):647–56.

24. Munafò MR, Tilling K, Taylor AE, Evans DM, Davey Smith G. Collider scope: when selection bias can substantially influence observed associations. Int J Epidemiol. 2018 Feb 1;47(1):226–35.

25. Gilbert JS, Nijland MJ. Sex differences in the developmental origins of hypertension and cardiorenal disease. Am J Physiol-Regul Integr Comp Physiol. 2008 Dec;295(6):R1941–52.

26. Borenstein M, Hedges LV, Higgins JPT, Rothstein HR. A basic introduction to fixed-effect and random-effects models for meta-analysis. Res Synth Methods. 2010 Apr;1(2):97– 111.

27. Cohen J. Statistical power analysis for the behavioral sciences. Second edition. Hillsdale, NJ: Lawrence Erlbaum Associates, Publishers; 1988. 1 p.

28. Lawlor D, Richmond R, Warrington N, McMahon G, Davey Smith G, Bowden J, et al. Using Mendelian randomization to determine causal effects of maternal pregnancy (intrauterine) exposures on offspring outcomes: Sources of bias and methods for assessing them. Wellcome Open Res. 2017 Feb 14;2:11.

29. Braun JM, Messerlian C, Hauser R. Fathers Matter: Why It’s Time to Consider the Impact of Paternal Environmental Exposures on Children’s Health. Curr Epidemiol Rep. 2017 Mar;4(1):46–55.

30. Ferraro ZM, Adamo KB. Maternal, Paternal, and Societal Efforts Are Needed to “Cure” Childhood Obesity. Mayo Clin Proc. 2015 Apr;90(4):555.

31. Soubry A. POHaD: why we should study future fathers. Environ Epigenetics [Internet]. 2018 Apr 1 [cited 2024 Aug 20];4(2). Available from: https://academic.oup.com/eep/article/doi/10.1093/eep/dvy007/4987171

32. Oldereid NB, Wennerholm UB, Pinborg A, Loft A, Laivuori H, Petzold M, et al. The effect of paternal factors on perinatal and paediatric outcomes: a systematic review and meta-analysis. Hum Reprod Update. 2018 May 1;24(3):320–89.

33. Easey KE, Sharp GC. The impact of paternal alcohol, tobacco, caffeine use and physical activity on offspring mental health: a systematic review and meta-analysis. Reprod Health. 2021 Dec;18(1):214.

34. Carter T, Schoenaker D, Adams J, Steel A. Paternal preconception modifiable risk factors for adverse pregnancy and offspring outcomes: a review of contemporary evidence from observational studies. BMC Public Health. 2023 Mar 16;23(1):509.

35. Sharp GC, Lawlor DA. Paternal impact on the life course development of obesity and type 2 diabetes in the offspring. Diabetologia. 2019 Oct 1;62(10):1802–10.

36. Rutkowska J, Lagisz M, Bonduriansky R, Nakagawa S. Mapping the past, present and future research landscape of paternal effects. BMC Biol. 2020 Dec;18(1):183.

37. Madley-Dowd P, Rai D, Zammit S, Heron J. Simulations and directed acyclic graphs explained why assortative mating biases the prenatal negative control design. J Clin Epidemiol. 2020 Feb;118:9–17.

38. Reiss F. Socioeconomic inequalities and mental health problems in children and adolescents: A systematic review. Soc Sci Med. 2013 Aug;90:24–31.

39. Ogden CL, Lamb MM, Carroll MD, Flegal KM. Obesity and socioeconomic status in children and adolescents: United States, 2005-2008. NCHS Data Brief. 2010 Dec;(51):1–8.

40. Poulain T, Vogel M, Sobek C, Hilbert A, Körner A, Kiess W. Associations Between Socio-Economic Status and Child Health: Findings of a Large German Cohort Study. Int J Environ Res Public Health. 2019 Feb 26;16(5):677.

41. Andréasson S. Alcohol and J-Shaped Curves. Alcohol Clin Exp Res [Internet]. 1998 May [cited 2024 Aug 20];22(S7). Available from: https://onlinelibrary.wiley.com/doi/10.1111/j.1530-0277.1998.tb04391.x

42. Tyrrell J, Huikari V, Christie JT, Cavadino A, Bakker R, Brion MJA, et al. Genetic variation in the 15q25 nicotinic acetylcholine receptor gene cluster (CHRNA5–CHRNA3–CHRNB4) interacts with maternal self-reported smoking status during pregnancy to influence birth weight. Hum Mol Genet. 2012 Dec 15;21(24):5344–58.

43. Rice F, Harold GT, Boivin J, Hay DF, van den Bree M, Thapar A. Disentangling prenatal and inherited influences in humans with an experimental design. Proc Natl Acad Sci U S A. 2009 Feb 17;106(7):2464–7.

44. Brand JS, Gaillard R, West J, McEachan RRC, Wright J, Voerman E, et al. Associations of maternal quitting, reducing, and continuing smoking during pregnancy with longitudinal fetal growth: Findings from Mendelian randomization and parental negative control studies. Minelli C, editor. PLOS Med. 2019 Nov 13;16(11):e1002972.

45. Yang Q, Millard LAC, Davey Smith G. Proxy gene-by-environment Mendelian randomization study confirms a causal effect of maternal smoking on offspring birthweight, but little evidence of long-term influences on offspring health. Int J Epidemiol. 2020 Aug 1;49(4):1207–18.

46. Dolan CV, Geels L, Vink JM, Van Beijsterveldt CEM, Neale MC, Bartels M, et al. Testing Causal Effects of Maternal Smoking During Pregnancy on Offspring’s Externalizing and Internalizing Behavior. Behav Genet. 2016 May;46(3):378–88.

47. Lavigne JV, Hopkins J, Gouze KR, Bryant FB, LeBailly SA, Binns HJ, et al. Is Smoking During Pregnancy a Risk Factor for Psychopathology in Young Children? A Methodological Caveat and Report on Preschoolers. J Pediatr Psychol. 2011 Jan 1;36(1):10–24.

48. Sallis HM, Wootton RE, Davey Smith G, Munafò MR. Proxy gene-by-environment Mendelian randomization study of the association between cigarette smoking during pregnancy and offspring mental health. Int J Epidemiol. 2023 Oct 5;52(5):1350–9.

49. Taylor AE, Carslake D, de Mola CL, Rydell M, Nilsen TIL, Bjørngaard JH, et al. Maternal Smoking in Pregnancy and Offspring Depression: a cross cohort and negative control study. Sci Rep. 2017 Oct 3;7(1):12579.

50. Haan E, Westmoreland KE, Schellhas L, Sallis HM, Taylor G, Zuccolo L, et al. Prenatal smoking, alcohol and caffeine exposure and offspring externalizing disorders: a systematic review and meta-analysis. Addiction. 2022 Oct;117(10):2602–13.

51. Gustavson K, Ystrom E, Stoltenberg C, Susser E, Surén P, Magnus P, et al. Smoking in Pregnancy and Child ADHD. Pediatrics. 2017 Feb;139(2):e20162509.

52. Havdahl A, Wootton RE, Leppert B, Riglin L, Ask H, Tesli M, et al. Associations Between Pregnancy-Related Predisposing Factors for Offspring Neurodevelopmental Conditions and Parental Genetic Liability to Attention-Deficit/Hyperactivity Disorder, Autism, and Schizophrenia: The Norwegian Mother, Father and Child Cohort Study (MoBa). JAMA Psychiatry. 2022 Aug 1;79(8):799.

53. Kim B, Ha M, Kim YS, Koh YJ, Dong S, Kwon HJ, et al. Prenatal exposure to paternal smoking and likelihood for autism spectrum disorder. Autism. 2021 Oct;25(7):1946–59.

54. Costa ADA, Almeida MTC, Maia FA, Rezende LFD, Saeger VSDA, Oliveira SLN, et al. Transtorno do espectro do autismo e o uso materno e paterno de medicamentos, tabaco, álcool e drogas ilícitas. Ciênc Saúde Coletiva. 2024;29(2):e01942023.

55. Lawlor DA, Timpson NJ, Harbord RM, Leary S, Ness A, McCarthy MI, et al. Exploring the Developmental Overnutrition Hypothesis Using Parental–Offspring Associations and FTO as an Instrumental Variable. Ludwig D, editor. PLoS Med. 2008 Mar 11;5(3):e33.

56. Easey KE, Gkatzionis A, Millard LAC, Tilling K, Lawlor DA, Sharp GC. Challenges in using data on fathers/partners to study prenatal exposures and offspring health. J Dev Orig Health Dis. 2024;15:e25.

57. Snowden JM, Bovbjerg ML, Dissanayake M, Basso O. The curse of the perinatal epidemiologist: inferring causation amidst selection. Curr Epidemiol Rep. 2018 Dec;5(4):379–87.

58. Burgess S, Davey Smith G, Davies NM, Dudbridge F, Gill D, Glymour MM, et al. Guidelines for performing Mendelian randomization investigations: update for summer 2023. Wellcome Open Res. 2023 Aug 4;4:186.

59. Shawe J, Delbaere I, Ekstrand M, Hegaard HK, Larsson M, Mastroiacovo P, et al. Preconception care policy, guidelines, recommendations and services across six European countries: Belgium (Flanders), Denmark, Italy, the Netherlands, Sweden and the United Kingdom. Eur J Contracept Reprod Health Care. 2015 Mar 4;20(2):77–87.

60. Bambra C, Gibson M, Sowden A, Wright K, Whitehead M, Petticrew M. Tackling the wider social determinants of health and health inequalities: evidence from systematic reviews. J Epidemiol Community Health. 2010 Apr 1;64(4):284–91.

61. Kirkbride JB, Anglin DM, Colman I, Dykxhoorn J, Jones PB, Patalay P, et al. The social determinants of mental health and disorder: evidence, prevention and recommendations. World Psychiatry. 2024 Feb;23(1):58–90.

62. Marmot M, Allen J, Boyce T, Goldblatt P, Morrison J. Health Equity in England: The Marmot Review 10 Years On [Internet]. Institute of Health Equity; 2020. Available from: health.org.uk/publications/reports/the-marmot-review-10-years-on

