## Supplementary File S1: Cohort Information for "Exploring parental prenatal influences on child health: a multicohort study and data visualisation tool"

This file contains detailed information about the cohorts that contributed to EPoCH, and how variables were defined in each.

### Cohorts

#### Avon Longitudinal Study of Parents and Children (ALSPAC)

Cohort description: ALSPAC is an ongoing population-based birth cohort study that recruited pregnant women residing in Avon, UK with expected dates of delivery between 1st April 1991 and 31st December 1992 (1,2). The core sample consisted of 14,541 pregnancies resulting in 14,062 live births and 13,988 children who were alive at the age of 1 Of the original 14,541 initial pregnancies, 338 were from a woman who had already enrolled with a previous pregnancy, meaning 14,203 unique mothers were initially enrolled in the study. Partners were invited to complete questionnaires by the mothers at the start of the study and they were not formally enrolled at that time. Participants have been regularly followed up through clinic visits and questionnaires. Ethics approval for the study was obtained from the ALSPAC Ethics and Law Committee and the Local Research Ethics Committees. Consent for biological samples has been collected in accordance with the Human Tissue Act (2004). The study website (http://www.bristol.ac.uk/alspac/researchers/our-data/) contains details of all the data that is available through a fully searchable data dictionary and variable search tool.

Cohort acknowledgements: We are extremely grateful to all the families who took part in this study, the midwives for their help in recruiting them, and the whole ALSPAC team, which includes interviewers, computer and laboratory technicians, clerical workers, research scientists, volunteers, managers, receptionists and nurses.

Cohort funding: The UK Medical Research Council and Wellcome (Grant ref: 217065/Z/19/Z) and the University of Bristol provide core support for ALSPAC. This publication is the work of the authors and Gemma Sharp will serve as guarantor for the contents of this paper. A comprehensive list of grants funding is available on the ALSPAC website (http://www.bristol.ac.uk/alspac/external/documents/grant-acknowledgements.pdf).

#### Born in Bradford (BiB)

Cohort description: BiB recruited pregnant women residing in Bradford, UK between 2007 and 2011, and who planned to give birth at the Bradford Royal Infirmary (3). After being provided with initial study information at their first antenatal clinic appointment, pregnant women were largely recruited at a routine oral glucose tolerance test (OGTT) at 24-28 weeks’ gestation. Women with pre-existing diabetes were not invited for the routine OGTT. Around 80% of pregnant women in Bradford, without pre-existing diabetes, attended for an OGTT, and about 80% of these participated in BiB, resulting in the recruitment of 12,453 women with 13,776 pregnancies resulting in 13,740 live births. Ethical approval for BiB was obtained from the Bradford Research Ethics Committee. Mothers were not asked to report fathers/partners behaviours. Father/partner data was collected directly via self-report, through invitations to participate if they attended the OGTT or subsequent antenatal appointment with the pregnant study mother, or postnatally when a member of the study team visited the family home during follow-up visits.

Cohort acknowledgements:

Born in Bradford is only possible because of the enthusiasm and commitment of the children and parents in BiB. We are grateful to all the participants, health professionals, schools and researchers who have made Born in Bradford happen.

Cohort funding:

BiB was supported by a joint grant from the UK Medical Research Council (MRC) and UK Economic and Social Science Research Council (ESRC) (MR/N024391/1); the British Heart Foundation (CS/16/4/32482); a Wellcome Infrastructure Grant (WT101597MA); the National Institute for Health Research under its Applied Research Collaboration for Yorkshire and Humber (NIHR200166). The National Institute for Health Research Clinical Research Network provided research delivery support for this study. The views expressed in this publication are those of the authors and not necessarily those of the National Institute for Health Research or the Department of Health and Social Care.

#### Millennium Cohort Study (MCS)

Cohort description: MCS is a UK nationally representative cohort study that recruited parents of children born between 1^st^ September 2000 and 31^st^ August 2001 (for England and Wales), and between 24^th^ November 2000 and 11^th^ January 2002 (for Scotland and Northern Ireland) (4). The children had to be alive and living in the UK at age 9 months. Eligible children were identified using government child benefit records, which has almost universal coverage. Certain subgroups were intentionally over-sampled (children living in disadvantaged areas, children of ethnic minority backgrounds, and children growing up in the smaller nations of the UK) to help ensure that typically hard to reach populations were adequately represented. In the first sweep, 18,827 children from 18,552 families were recruited. Ethics approval for the study was obtained from an NHS Research Ethics Committee (MREC) and informed consent was obtained from parents, as well as from the children themselves as they grow up.

Cohort acknowledgements:

We are grateful to the Centre for Longitudinal Studies (CLS), UCL Social Research Institute, for the use of these data and to the UK Data Service for making them available, and to the children and families who take part in the study. Neither CLS nor the UK Data Service bear any responsibility for the analysis or interpretation of these data.

Cohort funding:

The Millennium Cohort Study is supported primarily by the UK Economic and Social Research Council with co-funding from a consortium of UK government departments.

#### Norwegian Mother, Father and Child Cohort Study (MoBa)

Cohort description: MoBa is a prospective population-based pregnancy cohort study conducted by the Norwegian Institute of Public Health (5). It recruited pregnant women from all over Norway from 1999 to 2008. Invitations were sent to women in 277,702 pregnancies, the participation rate of pregnant women was 41%. The total cohort includes approximately 95,200 mothers, 75,200 fathers and 114,500 children. Blood samples were obtained from both parents during pregnancy and from mothers and children (umbilical cord) at birth (6). We used the genotype data quality-controlled by MoBaPsychGen (7). As well as through data linkage to the the Medical Birth Registry (MBRN), a national health registry containing information about all births in Norway, parents and their children have been regularly followed up through questionnaires. MoBa is regulated by the Norwegian Health Registry Act. The current study was approved by the Regional Committee for Medical and Health Research Ethics of South/East Norway (2016/1702).

Cohort acknowledgements: The Norwegian Mother, Father and Child Cohort Study is supported by the Norwegian Ministry of Health and Care Services and the Ministry of Education and Research. We are grateful to all the participating families in Norway who take part in this on-going cohort study. The MoBa analyses were performed on the TSD (Tjeneste for Sensitive Data) facilities, owned by the University of Oslo, operated and developed by the TSD service group at the University of Oslo, IT Department (USIT), with resources provided by Sigma2 - the National Infrastructure for High-Performance Computing and Data Storage in Norway.

Cohort funding: For generating high-quality genomic data, we thank the Norwegian Institute of Public Health (NIPH), the HARVEST collaboration, the NORMENT Centre at the University of Oslo, the Center for Diabetes Research at the University of Bergen, deCODE Genetics, the Research Council of Norway, the South-Eastern and Western Norway Regional Health Authorities, the ERC AdG, Stiftelsen KG Jebsen, the Trond Mohn Foundation, and the Novo Nordisk Foundation.

### Sample selection

Within each cohort, we excluded any children from multiple pregnancies. Figure S1 shows how the final sample sizes included in our analyses were reached.

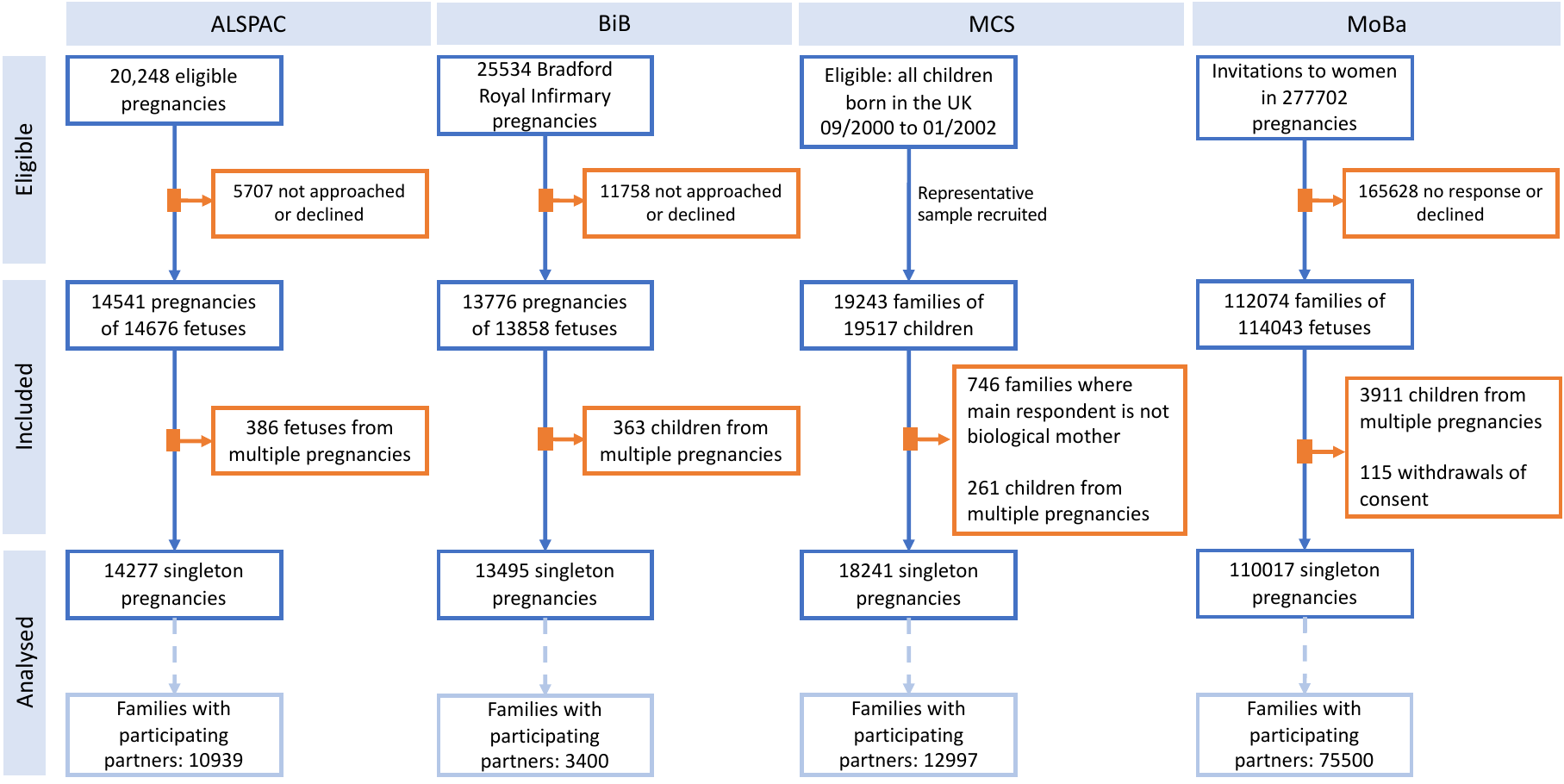

Figure S1. Flowcharts describing how final sample sizes were obtained.

### Harmonisation

Measures across cohorts and between maternal and paternal measures were derived the same way where the data allowed. Continuous measures were standardized into z-scores. Supplementary File S2 shows the availability of variables across cohorts.

#### Exposures

Where the data were available, we derived each exposure variable for both mothers and partners. For each health behaviour, we derived several variables based on the nature and timing of exposure, to enable us to explore associations with more granularity, and to generate variables harmonised across the four cohorts.

Exposure times: Preconception (up to 3 months before birth), first trimester (up to end of 12 weeks), second trimester (week 13-26), third trimester (week 27+), postnatally (birth-1000 days), unless stated otherwise.

##### Smoking

**ALSPAC**

Mothers were asked during three pregnancy questionnaires about their smoking behaviours during pregnancy, in the three months prior to pregnancy (preconception), and historically (e.g., whether they’d ever been a regular smoker, and the age at which they had their first cigarette). They were asked how long they spent in smoky rooms on weekends and weekdays (passive smoke exposure). They were also asked about postnatal smoking in two questionnaires before the child reached age 2.

Partners were asked in one pregnancy questionnaire about their smoking behaviours during pregnancy, and historically. They were not asked specifically about smoking in the three months prior to pregnancy, just about smoking “at the start” of pregnancy. This meant it was not possible to distinguish between preconception and first trimester. We chose to label the ‘start of pregnancy’ variable as first trimester, while acknowledging that many partners would not have changed their smoking behaviours a lot between pre-conception and the first trimester. Partners were also asked about postnatal smoking in two questionnaires before the child reached age 2.

Mothers were also asked to report on their partner’s age at smoking initiation and smoking around the start of pregnancy. In these cases, we derived two variables: one for self-report and one for mother-report. We found good agreement between the two, but the data were more complete for self-report. For our analyses, we derived composite variables, with self-report as the primary variable and missing values assigned with the mother-report value where it existed.

For one or both parents, we derived:

- Ordered categorical measures: none = 0, light = 1-9, moderate = 10-19, heavy = 20+. For postnatal smoking, two questionnaires applied to a single timepoint, so we took the maximum value across the timepoints.
- Binary measures of ever vs never smoked during/around pregnancy were derived from these ordered categorical measures: always reported ‘none’ (or none and missing) vs reported light/moderate/heavy at least once.
- Binary measure for ever smoking up until the birth (ever in life): yes vs no.
- Binary measure for smoking before the age of 12: yes vs no.
- Binary measure for passive smoke exposure: no = <1 hour per day and yes = 1+ hour per day.

**BIB**

Mothers were asked about their smoking behaviours in the baseline questionnaire. They were asked whether they were a current, former, or never smoker, the age at which they started smoking, the number of cigarettes they smoked per day in the three months before pregnancy (preconception), in the first trimester of pregnancy, and since the fourth month of pregnancy. The wording of this question meant it was not possible to distinguish between second and third trimester, so we derived the same variable for both. Mothers were also asked if they had been exposed to passive smoke during pregnancy.

Partners were asked about their smoking behaviours in their baseline questionnaire. They were asked whether they were a current, former, or never smoker, and the number of cigarettes they smoked “nowadays”, which would have been the third trimester.

For one or both parents, we derived:

- Ordered categorical measures: none = 0, light = 1-9, moderate = 10-19, heavy = 20+
- Binary measures of ever vs never smoked during/around pregnancy were derived from these ordered categorical measures: always reported ‘none’ (or none and missing) vs reported light/moderate/heavy at least once.
- Binary measure for ever smoking up until birth (ever in life): yes vs no.
- A binary measure for smoking before the age of 12: yes vs no.
- Passive smoke exposure: no = “no” or <1 hour per day and yes = “yes”.

**MCS**

Mothers AND partners were asked about their smoking behaviours in the baseline interview, which was after the birth of the child. They were asked whether they were a current, former, or never smoker, what they smoked, how many they smoked ‘just before pregnancy’ (preconception), whether their smoking habits changed during pregnancy, when, and how much they smoked per day after the change.

For one or both parents, we derived:

- Ordered categorical measures: none = 0, light = 1-9, moderate = 10-19, heavy = 20+ cigarettes per day. These were derived for specific trimesters and postnatally using data from the questions on smoking before pregnancy, when smoking habits changed during pregnancy, and how much was smoked after the change, and smoking now (at interview).
- Binary measures of ever vs never smoked during/around pregnancy were derived from these ordered categorical measures: always reported ‘none’ (or none and missing) vs reported light/moderate/heavy at least once.
- Binary measure for ever smoking up until the interview (ever in life): yes vs no.

**MoBa**

Mothers were asked about smoking behaviours at multiple stages during the pregnancy, preconception and postnatal. They were asked about their current smoking status, frequency of smoking, and how many cigarettes they smoke. Also, they were asked if they smoked before getting pregnant, and smoking patterns during pregnancy.

Partners were also asked about their smoking behaviour preconception, during pregnancy at different stages. They were asked about current smoking status, frequency of smoking, and how many cigarettes they smoke. In addition, mothers were asked to report on their partner’s smoking behaviours preconception, and in the second and third trimester. In this case, we derived composite binary variables defining “no” as BOTH mother AND self-report smoking ==”no”, and “yes” as EITHER mother OR self-report smoking == “yes”.

Derived for Mothers

- Ordered categorical measures: none = 0, light = 1-9, moderate = 10-19, heavy = 20+ cigarettes per day at 3 months preconception, second trimester, third trimester, and postnatally.
- Binary measures (yes/no) of ever smoked up to pregnancy, ever smoked <=age 11, smoked 3 months preconception, any smoking in second trimester, third trimester, any time in pregnancy, passive smoke in pregnancy, and postnatally.

Derived for Partners

- Ordered categorical measures of number of cigarettes per day (none, light: 1-4, light to moderate: 5-9, moderate to heavy: 10-19, heavy: 20+) at preconception, second trimester, third trimester (mother report only), any time in pregnancy, postnatally.
- Binary measures (yes/no) of any smoking ever up to birth, maternal report of any smoking ever up to birth (yes/no), preconception, second trimester, second trimester (mother report), third trimester (mother report only), anytime in pregnancy, postnatally.

##### Alcohol consumption

**ALSPAC**

Mothers were asked about their alcohol consumption during three pregnancy questionnaires: before current pregnancy, during pregnancy, in the three months prior to pregnancy (preconception). They were asked about whether they consume alcohol and how much do they consume currently. During the pregnancy they were asked about whether in the past month they consumed the equivalent of 2 pints of beer, 4 glasses of wine or 4 pub measures of spirit, and the frequency during the month (none, 1-2 days, 3-4 days, 5-10 days, more than 10 days and every day). Also, during pregnancy the mothers were asked how much they usually drink during a day and the type of drinks (beer/lager in pints, wine by glasses, spirits by pub measures, and other alcoholic beverages by pub measures). In the second trimester, they were asked “how much were you drinking at around the time you first felt the baby move, or if you haven't felt the baby move yet, the last 2 weeks". People usually feel the baby kick between 16 and 25 weeks (first trimester), so we can pretty safely assume this question relates to the second trimester. The alcohol consumption behaviour was also asked in three of the questionnaires postnatally before the child reached age 2 (at 8 weeks, 8 months, and 21 months postnatally).

Partners were asked to fill questionnaires during the pregnancy of the mother where they reported how often they drank alcohol pre-conception (in the three months prior to pregnancy) and in the second and third trimester (none, 1-2 days, 3-4 days, 5-10 days, more than 10 days and every day). Also, in the second trimester, they were asked how many days in the past month they consumed the equivalent of 2 pints of beer, 4 glasses of wine or 4 pub measures of spirit, and the frequency during the month (none, 1-2 days, 3-4 days, 5-10 days, more than 10 days and every day), and the type of alcohol they drank often (wine, beer/lager, sherry/port, gin/whisky/vodka/brandy, other drinks, don’t drink at all). The alcohol consumption behaviour was also asked in three of the questionnaires postnatally before the child reached age 2 (at 8 weeks, 8 months, and 21 months postnatally).

The mothers were also asked about their partners alcohol drinking behaviour ever during pregnancy with the following categories: never drinks alcohol, very occasionally (less than once a week), occasionally (at least once a week), drinks 1-2 glasses (nearly everyday), drinks 3-9 glasses (everyday), drinks at least 10 glasses a day and don’t know. In these cases, we derived two variables: one for self-report and one for mother-report. We found good agreement between the two, but the data were more complete for self-report. For our analyses, we derived composite variables, with self-report as the primary variable and missing values assigned with the mother-report value where it existed.

Derived for Mothers

- Ordered categorical measures of number of drinks per day (none, light: < 1 glass per week, moderate: 1+ glasses per week but not daily, heavy: daily drinking with at least 1 glass per day) at preconception, first trimester, second trimester, third trimester, postnatally.
- Binary measures (yes/no) of any alcohol at preconception, first trimester, second trimester, third trimester; any binge drinking (2 pints of beer or equivalent in past month) in first trimester, third trimester, anytime in pregnancy, postnatally; and binge drinking (yes/no) during the first trimester, third trimester or at any point during the pregnancy.

Derived for Partners

- Ordered categorical measures of number of drinks per day (none, light: < 1 glass per week), moderate: 1+ glasses per week but not daily, heavy: at least 1 per day) at preconception (3 months prior to pregnancy), second trimester, third trimester, postnatally.
- Binary measures (yes/no) of any alcohol preconception, second trimester, third trimester, any time in pregnancy, postnatally; and binge drinking (yes/no 4+ units of alcohol) in the second trimester.

**BiB**

Mothers were asked about their alcohol behaviours in the baseline questionnaire. They were asked whether they drank alcohol during pregnancy, three months before pregnancy (preconception), and post pregnancy up to two years. The frequency of alcohol consumption was reported as drinking every day, nearly every day, 1-4 times/week, 1-3 times a month, rarely and never.

Partners were also asked in the baseline questionnaire whether they drink alcohol and what was their weekly average no. of unit's consumption, in the third trimester.

Derived for Mothers

- Ordered categorical measures of number of drinks per day (none, light: < 1 glass per week, moderate: 1+ glasses per week but not daily, heavy: at least 1 per day) at preconception, first trimester, second trimester (first and second trimester measures are identical due to how/when the question was asked), third trimester.
- Binary measures (yes/no) of any alcohol at preconception, first trimester, second trimester, third trimester (first and second trimester measures are identical); any binge drinking (5 or more drinks on any occasion) in first trimester, second, third trimester (first and second trimester measures are identical), anytime in pregnancy.

Derived for Partners

- Ordered categorical measures of number of drinks per day (none, light: < 1 glass per week), moderate: 1+ glasses per week but not daily, heavy: at least 1 per day) third trimester.
- Binary measures (yes/no) of third trimester, any time in pregnancy (third trimester measure is identical to any time in pregnancy)

**MCS**

Mothers AND partners were asked about their alcohol behaviours in the baseline interview, which was after the birth of the child. They were asked about how frequently they drank alcohol (every day, 5-6 times per week, 3-4 times per week, 1-2 times per week, 1-2 times per month, less than once a month; never), whether their drinking pattern was different during pregnancy, and units of alcohol consumption (half pint of beer, a glass of wine, or a single measure of spirit or liqueur).

Derived for Mothers:

- Ordered categorical measures of number of drinks per day (none, light: <1 glass per week, moderate: 1+ glass per week but not daily, heavy: daily drinking or over 1-2 glasses per day) at any time in pregnancy, postnatally.
- Binary measures (yes/no) of any alcohol at anytime during pregnancy, postnatally in first two years.

Derived for Partners:

- Ordered categorical measures of number of drinks per day (none, light: <1 glass per week, moderate: 1+ glass per week but not daily, heavy: daily drinking or over 1-2 glasses per day) at any time in pregnancy, postnatally.
- Binary measures (yes/no) of any alcohol in first two postnatal years.

**MoBa**

Mothers were asked about their alcohol behaviour at preconception, at multiple stages during the pregnancy, and after pregnancy. They were asked if they ever consumed alcohol (yes/no), if they consumed alcohol three months before pregnancy (pre-conception), during pregnancy, along with the frequency of consumption (approximately 6-7 times a week, approximately 4-5 times a week, approximately 2-3 times a week, approximately once a week, approximately 1-3 times a month, less than once a month and never). They were also asked about the type of alcohol they usually consumed (light beer, beer, red wine, white wine, low alcohol sodas, fortified wines, spirits), if they consumed 5 units or more at least once in the last three months before pregnancy and during pregnancy along with the frequency of it (several times per week, once a week, 1-3 times a month, less than once a month, never).

Partners were also asked about their alcohol behaviours at preconception and second trimester. They were also asked if they ever consumed alcohol (yes/no), if they consumed alcohol six months before pregnancy (pre-conception), during pregnancy, along with the frequency of consumption (approximately 6-7 times a week, approximately 4-5 times a week, approximately 2-3 times a week, approximately once a week, approximately 1-3 times a month, less than once a month and never). They were also asked if they consumed 5 units or more at least once in the last six months before pregnancy and during pregnancy along with the frequency of it (several times per week, once a week, 1-3 times a month, less than once a month, never).

Derived for Mothers

- Ordered categorical measures of usual number of alcohol units per day (none; light: less than once per month, approximately 1-3 times a month, approximately once a week; moderate: approximately 2-3 times a week, approximately 4-5 times a week; heavy: approximately 6-7 times a week) at preconception, first trimester, second trimester, third trimester, postnatally.
- Binary measures (yes/no) of any alcohol at preconception, first trimester, second trimester, third trimester, anytime during pregnancy; any binge drinking (5+ units or more at least once: several times a week/once per week/less than once a month/ 1-3 times per month, versus, never) at preconception, in first trimester, second trimester, third trimester, anytime in pregnancy, postnatally.

Derived for Partners

- Ordered categorical measures of usual number of alcoholic units when drink (one; light: less than once per month, approximately 1-3 times a month, approximately once a week; moderate: approximately 2-3 times a week, approximately 4-5 times a week; heavy: approximately 6-7 times a week) at preconception (3 months prior to pregnancy), second trimester, postnatally.
- Binary measures (yes/no) of any alcohol at preconception, second trimester, anytime during pregnancy; any binge drinking (5+ units or more at least once: several times a week/once per week/less than once a month/ 1-3 times per month, versus, never) at preconception, second trimester and anytime during pregnancy.

##### Caffeine consumption

**ALSPAC**

Mothers and partners were asked how many cups of tea (herbal tea not included), cola and coffee they drink in a day, ever during pregnancy and postnatally.

Derived for Maternal:

- Continuous measures of (mg given of caffeine content) coffee cups per day (1 cup: 57mg), tea (1 cup: 27mg), cola (1 cup: 20mg), total caffeine drinks, in first trimester, second trimester, third trimester, postnatally; average caffeine per day in pregnancy (cups per day).
- Binary measures (yes/no) of any caffeine in first trimester, second trimester, third trimester, anytime in pregnancy, postnatally.
- Ordinal categorical measure of total caffeine levels (none, light: less than 200mg, moderate: 200-400mg and heavy: 400mg or more, derived from the continuous measures of caffeine per week) in first trimester, second trimester, third trimester, postnatally, ever in pregnancy.

Derived for Partner:

- Continuous measures of (mg given of caffeine content per cup) cups per day of coffee (1 cup: 57mg), tea (1 cup: 27mg), cola (1 cup: 20mg), total caffeinated drinks, in second trimester and ever in pregnancy. It was derived as from any caffeine source and from each source.
- Binary measures (yes/no) of any caffeine in second trimester, any caffeine in pregnancy – same as second trimester measure.
- Ordinal categorical measure of total caffeine levels (none, light: less than 200mg, moderate: 200-400mg and heavy: 400mg or more, derived from the continuous measures of caffeine per week) in second trimester, any caffeine during pregnancy – same as second trimester measure.

**BiB**

Mothers were asked in the baseline questionnaire on how many cups of tea (includes caffeinated Kashmiri, herbal teas as well), cola and coffee (includes instant and filter coffees) they drink in a day and week, during the past 4 weeks of pregnancy and during a usual day.

Derived for Maternal:

- Continuous measures of (mg given of caffeine content) coffee cups per day (1 cup: 57mg), tea (1 cup: 27mg), cola (1 cup: 20mg), total caffeine drinks, in third trimester; average caffeine per day in pregnancy (cups per day) (third trimester measures are identical to average caffeine per day in pregnancy)
- Binary measures (yes/no) of any caffeine in third trimester, anytime in pregnancy.
- Ordinal categorical measure of total caffeine (none, light: less than 200mg, moderate: 200-400mg and heavy: 400mg or more, derived from the continuous measures of caffeine per week) in third trimester and ever in pregnancy.

**MCS**

Caffeine data not available.

**MoBa**

Mothers and partners were asked pre-conception and in second trimester of pregnancy on how frequently they consumed tea, cola (includes sugar and sugar free ones too) and coffee (includes instant and filter coffees)-seldom/never, 1-6 glass per week, 1 glass per day, 2-3 glasses per day, 4 glasses or more per day.

The caffeine content in mg were measured as 1 coffee cup: 57mg, 1 tea cup: 27mg, 1 cola cup: 20mg. Seldom or never is considered as 0 cups per day, 1-6 glasses per week is 0.43 cups per day (3 cups/7 days=0.43pd), 1 glass is 1 cup per day, 2-3 glasses are 2.5 cups per day and 4 glasses or more is taken as 4 cups per day.

Derived for Maternal:

- Binary measures (yes/no) of any caffeine in preconception, second trimester, anytime in pregnancy.
- Ordinal categorical measure of total caffeine (none, light: less than 200mg, moderate: 200-400mg and heavy: 400mg or more, derived from the continuous measures of caffeine per week) in preconception, second trimester and ever in pregnancy.

Derived for Partner:

- Binary measures (yes/no) of any caffeine in second trimester and ever in pregnancy.
- Ordinal categorical measure of total caffeine (none, light: less than 200mg, moderate: 200-400mg and heavy: 400mg or more, derived from the continuous measures of caffeine per week) in second trimester and ever in pregnancy.

##### Socioeconomic position

We defined socioeconomic position using highest educational qualification/level achieved and current occupation at the time of pregnancy. We derived for all the cohorts as a three or four level categorical variable.

**ALSPAC**

Mothers reported their and their partner’s education levels and occupation.

- Mother education (CSE/none = 0, O-level or vocational = 1, A-Level = 2, Degree = 3)
- Partner education (CSE/none = 0, O-level or vocational = 1, A-Level = 2, Degree = 3)
- Mother occupation (semi-skilled or unskilled=0, skilled manual= 1, skilled non-manual= 2, professional or managerial/technical= 3)
- Partner occupation (semi-skilled or unskilled= 0, skilled manual= 1, skilled non-manual= 2, professional or managerial/technical= 3).

**BiB**

Mothers reported their education levels as well as partner’s education levels and occupation.

- Mother education (<5 GCSE equivalent= 0, 5 GCSE equivalent= 1, A-level equivalent= 2, Higher than A-level=3)
- Partners education (<5 GCSE equivalent= 0, 5 GCSE equivalent= 1, A-level equivalent= 2, Higher than A-level=3)
- Partner occupation (semi-skilled or unskilled= 0, skilled manual=1, skilled non-manual= 2, professional or managerial= 3).

**MCS**

Mothers reported their and their partner’s education levels and occupation.

- Mother education (none= 0, O level or GCSE= 1, A level= 2, degree/HE diploma= 3)
- Partner education (none= 0, O level or GCSE= 1, A level= 2, degree/HE diploma= 3)
- Mother occupation (semi-routine or routine= 0, low supervisory and technical= 1, intermediate and small employer or self-employed= 2, professional or managerial= 3)
- Partner occupation (semi-routine or routine= 0, low supervisory and technical= 1, intermediate and small employer or self-employed= 2, professional or managerial= 3)

**MoBa**

Mothers reported their own and their partner’s education levels around week 15 of pregnancy according to six categories, which were harmonised with other cohorts into three levels as follows:

0 = 9-year elementary education

1 = Further education – vocational, Further education 1-2 years, Further education 3 years - (general studies, sixth form)

2 = Higher education (university/college), over 4 years, Higher education (university/college), up to and including 4 years

Mothers also reported their own and their partners’ occupation (employed; self-employed; military; unemployed; student/apprentice; domestric tasks; inactive/other), but we could not harmonise this information with occupation as reported by the other cohorts, so we omitted parental occupation from MoBa.

#### Outcomes

Data on child outcomes were collected at various follow-ups, either via questionnaires or interviews with the primary caregiver(s), study clinics, or via linkage to other data (e.g. obstetric birth records in ALSPAC). Depending on the outcome and data availability, we generated variables for the following ages:

- At birth/delivery
- In the first year of life (>0 to <1 years)
- Childhood stage 1 (around age 2; >=1 to <3)
- Childhood stage 2 (around age 4; >=3 to <5)
- Childhood stage 3 (around age 6; >=5 to <8)
- Childhood stage 4 (around age 9; >=8 to <11)

All continuous outcomes were converted to standard deviation units (z scores) so that they could be meta-analysed across different cohorts and visualised on the same scale.

##### Body size and composition

###### Birth length (cm)

**ALSPAC:** Study-preferred crown-heel length extracted from obstetrics records or measured by ALSPAC; **BiB**: height at 0 months taken from various sources (e.g. primary care, eClipse study); **MCS:** not available; **MoBa:** Not available in our dataset.

###### Birthweight (g)

**ALSPAC:** Study-preferred birthweight extracted from obstetrics records or measured by ALSPAC; **BiB**: Taken from routine healthcare records; **MCS:** parent reported birthweight in kg converted to g; **MoBa:** extracted from the Medica Birth Registry of Norway

###### Birthweight <2500g

Derived for all cohorts as a binary variable <2500g vs >=2500g.

###### Birthweight >4500g

Derived for all cohorts as a binary variable <=4500g vs >4500g.

###### Large for gestational age

**ALSPAC, BiB, and MoBa:** Birth weight centiles were calculated using UK WHO 1990 growth standards for ALSPAC and BiB, and unstandardised centiles for MoBa. LGA was defined as z score >=90^th^ centile. **MCS:** not available.

###### Small for gestational age

**ALSPAC, BiB, and MoBa:** Birth weight centiles were calculated using UK WHO 1990 growth standards for ALSPAC and BiB, and unstandardised centiles for MoBa. LGA was defined as z score <=10^th^ centile. **MCS:** not available.

###### Height

**ALSPAC:** Height was measured using a Harpenden stadiometer. Derived as a continuous variable for height in cm. **BiB and MCS**: Height was measured using Leicester Height Measure (Seca digital scales). **MoBa**: Mother’s reported the child’s height. Derived as a continuous variable for height in cm.

###### Weight

**ALSPAC**: Weight was measured using Tanita body fat analyzer and weighing scales. **BiB**: Measured using standard weighing scales. **MoBa**: Mothers reported the child’s weight. **MCS**: Measured using Tanita BF-522 scales. Derived as a continuous variable for weight in kg.

###### Body Mass Index (BMI) (kg/m^2^) – not standardised

Derived for all cohorts as a continuous variable using weight (kg) / height (m)^2.

###### Body Mass Index (BMI) (kg/m^2^) – adjusted for age and sex

BMI was adjusted for age at measurement and sex and standardised according to the UK WHO 1990 growth standards using the childsds R package.

###### Overweight/obese

Overweight was defined as an age and sex standardised BMI value >1 (vs a ‘normal weight’ value <=1).

###### Obese

Overweight was defined as an age and sex standardised BMI value >2 (vs a ‘normal weight’ value <=1).

###### Fat mass index (FMI)

**ALSPAC:** total body fat mass measured via Dual-energy X-ray Absorptiometry (DXA) **and MCS:** Not available. **BiB**: Fat mass was measured by bioimpedance. Derived for all cohorts as a continuous variable, by dividing the fat mass by height squared **MoBa**: not available.

###### Fat mass percentage

**ALSPAC and BiB** Derived as a continuous variable using fat mass and total body weight as described above (fat mass (kg)/body weight (kg))*100. MCS: body fat percentage calculated by bioimpedance scales. **MoBa**: not available.

###### Head circumference

**ALSPAC:** The head circumference was measured using a taut tape just after birth and during each clinic visit. Derived as a continuous variable for head circumference in cm. **BiB:** Measured right after birth using Lasso-o tapes. The measurements were part of routine clinical check-ups and were recorded in the hospital’s e-record systems. Derived as a continuous variable for head circumference in cm. **MoBa**: Measured within 72 hours of birth, using a fibre glass tape against and fixed ruler. Derived as a continuous variable for head circumference in cm. **MCS**: not available.

###### Waist circumference

**ALSPAC:** The waist circumference was measured using a flexible tape measure, in the midpoint between the lowest rib and iliac crest, by touching the skin but not compressing it. The measurement was done in mm. Derived as a continuous variable in cm. **BiB:** Measured at the level of navel after birth. Derived as a continuous variable in cm. **MCS:** Measured using a non-elasticated SECA tape, midpoint between costal margin and iliac crest, and measurement taken to nearest mm. Derived as a continuous variable in cm. **MoBa:** not available

##### Immunological outcomes

###### Any allergy

Reported by mothers and derived for all cohorts as a binary variable whether having any particular allergy (as defined by the other allergy variables described below) or not.

###### Cat allergy

**ALSPAC**: Reported by mothers and was derived as a binary variable (yes/no). **BiB, MCS and MoBa**: not available.

###### Dog allergy

**ALSPAC**: Reported by mothers and was derived as a binary variable (yes/no). **BiB, MCS and MoBa**: not available.

###### Pet allergy

**ALSPAC:** Reported by mothers and derived from combining cat and dog allergies, as a binary variable (yes/no). **MoBa**: Reported by mothers at stage 1 according to whether the child had tested positive for animal allergy, and derived as a binary variable (yes/no). **BiB and MCS**: not available.

###### Dustmite allergy

**ALSPAC:** Reported by mothers as allergy to dust (stage 2, 3, 4). **MoBa**: Reported by mothers as a positive test for mite allergy (stage 1). **BiB and MCS**: not available.

###### Food allergy

**ALSPAC, BiB and MoBa:** Reported by mothers and derived for these cohorts as a binary variable (yes/no). **MCS**: not available

###### Insect allergy

**ALSPAC**: Reported by mothers and derived as a binary variable (yes/no). **BiB, MCS and MoBa**: not available

###### Pollen/hay allergy

**ALSPAC, BiB:** Pollen allergy was reported by mothers and derived for these cohorts as a binary variable (yes/no). **MCS**: Hay allergy was reported by mothers and derived as a binary variable (yes/no) derived as a binary variable. **MoBa:** reported by mothers as a positive test for pollen allergy.

###### Asthma

**ALSPAC**: Mothers reported if the child had asthma or taken medication for asthma in past year or if doctor said if the child ever had asthma. **BiB, MCS and MoBa**: reported by mothers if child ever had (recent) asthma or not. Where possible we derived stage-specific binary variables (ever in this stage vs never in this stage), as well as binary variables to describe whether the child had ever had asthma at any of the timepoints (ever vs never in childhood).

###### Eczema

**ALSPAC**: Mothers reported if the child had eczema or an eczema-like rash. **MoBa**: mothers reported whether the child had atopic eczema or another type of eczema now, in the past, or never. **BiB:** mothers reported whether the child had ever had eczema. **MCS:** parents reported whether the child had ever had eczema up to age 5. Where possible we derived stage-specific binary variables (ever in this stage vs never in this stage), as well as binary variables to describe whether the child had ever had eczema at any of the timepoints (ever vs never in childhood).

###### Wheezing

**ALSPAC:** mothers reported whether the child had wheezing or whistling on the chest since 6 months, before 12 months, after 18 months, or in the past year. **BiB:** mothers reported whether the child had ever had wheezing. **MCS:** Parents were asked whether the child had wheezing in the past 12 months. **MoBa:** Mothers were asked whether the child had wheezing now, in the past, or never. Where possible we derived stage-specific binary variables (ever in this stage vs never in this stage), as well as binary variables to describe whether the child had ever had wheezing at any of the timepoints (ever vs never in childhood).

##### Psychosocial and cognitive outcomes

*Note: only the measures considered to be measuring the same constructs were harmonised for cross-cohort meta-analysis. The remaining measures were analysed as separate outcomes. Some outcomes were binarized according to commonly used clinical thresholds because they were strongly skewed. Cohort instrument documentation includes more detail about each measure:* [*ALSPAC*](https://www.bristol.ac.uk/alspac/researchers/our-data/)*,* [*BiB*](https://borninbradford.github.io/datadict/)*,* [*MCS*](https://cls.ucl.ac.uk/cls-studies/millennium-cohort-study/?_gl=1*mtpzty*_up*MQ..*_ga*MTQzMjczMTc4LjE3NTQ4MTAxNTg.*_ga_EYRQV4V0KV*czE3NTQ4MTAxNTckbzEkZzAkdDE3NTQ4MTAxNTckajYwJGwwJGgw)*,* [*MoBa*](https://www.fhi.no/globalassets/dokumenterfiler/studier/den-norske-mor-far-og-barn--undersokelsenmoba/instrumentdokumentasjon/instrument-documentation-synthese.pdf.)*.*

###### Social communication difficulties and repetitive behaviours (M-CHAT and SCQ)

**MoBa**: Measured by the Modified Checklist for Autism in Toddlers (M-CHAT) scale for stage 1 and Social Communication Questionnaire (SCQ) for stages 2 and 3, to screen for early signs of autism. **ALSPAC, BiB and MCS:** Not available

###### Social communication difficulties and repetitive behaviours over the clinical threshold

**MoBa**: Derived as a binary variable with MCHAT scale cut off set at 7, (<7=’0’ & ≥7=’1’) and the SCQ set at 15 (<15=0; >=15=1). **ALSPAC, BiB and MCS:** Not available

###### Communication development (ASQ)

**MoBa:** Measured by the ages and stages questionnaire (ASQ) to assess the child’s social and communication development across all stages. **ALSPAC, BiB and MCS:** Not available.

###### Social and communication development (SCDC)

**ALSPAC**: Measured by the Skuse Social Communication Scores (SCDC) to assess the social and communication abilities of the child at all stages. **BiB, MCS, and MoBa**: Not available

###### Aggression

**MoBa:** Measured using aggression subscale of the Child Behavioural Checklist (CBCL). **ALSPAC, BiB, and MCS**: Not available

###### Conduct problems

**ALSPAC and MCS**: Measured by the conduct problems subscale of the strengths and difficulties questionnaire (SDQ) which is a behavioural screening tool and was completed by parents. **BiB and MoBa:** Not available

###### Conduct problems >=4

**ALSPAC and MCS**: Derived as a binary variable by setting the cutoff for conduct problems scores at 4 (<4=’0’ & ≥4=’1’) according to <https://sdqinfo.org/py/sdqinfo/c0.py>. **BiB and MoBa:** Not available.

###### Depressive symptoms

**ALSPAC and MoBa**: Measured by the Mood and Feelings Questionnaire (MFQ) completed by the parent. **BiB and MCS:** Not available.

###### Emotional symptoms

**ALSPAC and MCS**: Measured by the emotional symptoms subscale of the strengths and difficulties questionnaire (SDQ). **BiB and MoBa:** Not available

###### Emotional symptoms >=5

**ALSPAC and MCS**: Derived as a binary variable by setting the cutoff for emotional symptoms scores at 5 (<5=’0’ & ≥5=’1’) according to <https://sdqinfo.org/py/sdqinfo/c0.py>. **BiB and MoBa:** Not available.

###### Externalising traits

**ALSPAC and MCS**: Measured by the strengths and difficulties questionnaire (SDQ) with combination of the subscales: hyperactivity and conduct problems. **BiB:** Not available. **MoBa**: Measured using the Child Behaviour CheckList (CBCL) pre-school version short scale in MoBa for externalising behaviour, which is a combination of the scales for attention problems and aggressive behaviours.

###### Hyperactivity/inattention

**ALSPAC and MCS**: Measured by the hyperactivity subscale of the strengths and difficulties questionnaire (SDQ). Derived as a continuous variable based on the scores. **BiB:** Not available. **MoBa**: Measured using the Child Behaviour CheckList (CBCL) pre-school version short scale in MoBa using only the attention problems subscale.

###### Hyperactivity/inattention >=7

**ALSPAC and MCS**: Derived as a binary variable by setting the SDQ cutoff for hyperactivity scores at 7 (<7=’0’ & ≥7=’1’) according to <https://sdqinfo.org/py/sdqinfo/c0.py>. **BiB and MoBa:** Not available.

###### Internalising traits

**ALSPAC and MCS**: Measured by the strengths and difficulties questionnaire (SDQ) internalising scale, which is a combination of subscales Emotional symptoms and Peer problems. Derived as a continuous variable based on the scores. **BiB:** Not available. **MoBa**: Measured using the internalising behavior scale from the Child Behaviour CheckList (CBCL) pre-school version short scale in MoBa.

###### Peer relationship problems

**ALSPAC and MCS**: Measured by the peer problems subscales of the strengths and difficulties questionnaire (SDQ). **BiB and MoBa:** Not available

###### Peer relationship problems >=4

**ALSPAC and MCS**: Derived as a binary variable by setting the cutoff for peer problems scores at 4 (<4=’0’ & ≥4=’1’) according to <https://sdqinfo.org/py/sdqinfo/c0.py>. **BiB and MoBa:** Not available.

###### Prosocial behaviour

**ALSPAC, MCS and MoBa**: Measured by the prosocial behaviour subscale of the strengths and difficulties questionnaire (SDQ). **BiB:** Not available.

###### Prosocial behaviour <=6

**ALSPAC, MCS and MoBa**: Derived as a binary variable by setting the cutoff for prosocial behaviour scores at 6 (<6=’0’ & ≥6=’1’) according to <https://sdqinfo.org/py/sdqinfo/c0.py>. **BiB:** Not available.

###### Social behaviour: emotional dysregulation

**MCS**: Measured by the emotional dysregulation subscale of the Child Social Behaviour Questionnaire (CSBQ). **ALSPAC, BiB and MoBa**: Not available

###### Social behaviour: independence

**MCS**: Measured by the independence subscale of the Child Social Behaviour Questionnaire (CSBQ). **ALSPAC, BiB and MoBa**: Not available.

###### Total behavioural score (CBCL)

**MoBa**: Measured by summing the scores from the Child Behavior Checklist (CBCL). **ALSPAC, BiB and MCS**: Not available

###### Total difficulties (SDQ)

**ALSPAC and MCS**: Measured by summing all the subscales of the strengths and difficulties questionnaire (SDQ), except the prosocial behaviour subscale. **BiB and MoBa:** Not available

###### Total difficulties (SDQ) >=17

**ALSPAC and MCS**: Derived as a binary variable by setting the cutoff for total difficulties scores at 17 (<17=’0’ & ≥17=’1’) according to <https://sdqinfo.org/py/sdqinfo/c0.py>. **BiB and MoBa:** Not available.

###### Naming vocabulary

**MCS**: Measured by a verbal scale of the British Ability Scales (BAS) for children aged 2 years 6 months to 7 years 11 months. It assesses the spoken vocabulary of young children. **ALSPAC, BiB and MoBa**: Not available

###### Number skills

**MCS**: Measured by the number skills test adapted from the NFER Progress in Maths test which is aimed for 7 year olds and was originally developed and nationally UK standardised in 2004. **ALSPAC, BiB and MoBa**: Not available

###### Problem solving

**MCS**: Measured using the picture similarities subtest from the British Ability Scales Second Edition (BAS II). This evaluation occurs during the age 5 sweep. **ALSPAC, BiB and MoBa**: Not available.

###### Reading skills

**MCS**: Measured using the word reading assessment from the British Ability Scales Second Edition (BAS II), for the age 7 sweep. **ALSPAC, BiB and MoBa**: Comparable measure not available.

###### School readiness

**MCS**: Measured using the school readiness assessment of the Bracken Basic Concept Scale –Revised (BBCS-R), for the age 3 sweep. **ALSPAC, BiB and MoBa**: Comparable measure not available.

###### Spatial awareness

**MCS**: Measured using the pattern construction subtest from the British Ability Scales Second Edition (BAS II), for the age 5 and 7 sweep. **ALSPAC, BiB and MoBa**: Not available.

###### Verbal intelligence

**ALSPAC**: Measured by the verbal subsets in the Wechsler Preschool and Primary Scale of Intelligence – Revised (WPPSI-R) for age 4 children (stage 2) and the Wechsler Intelligence Scale for Children – Third Edition (WISC-III) for age 8 years (stage 4). **BiB, MCS and MoBa**: Not available.

###### Performance intelligence

**ALSPAC**: Measured by the performance (non-verbal) subsets of the Wechsler Preschool and Primary Scale of Intelligence – Revised (WPPSI-R) for age 4 children (stage 2) and the Wechsler Intelligence Scale for Children – Third Edition (WISC-III) for age 8 years (stage 4). **BiB, MCS and MoBa**: Not available.

###### Total intelligence

**ALSPAC**: Measured by the combination of verbal and performance scales of the Wechsler Preschool and Primary Scale of Intelligence – Revised (WPPSI-R) for age 4 children (stage 2) and the Wechsler Intelligence Scale for Children – Third Edition (WISC-III) for age 8 years (stage 4). **BiB, MCS and MoBa**: Not available.

##### Serum biomarkers

###### Apolipoprotein A1

**ALSPAC**: Measured using an automated analyser and derived as a continuous variable in g/L. **BiB, MCS, and MoBa**: Not available.

###### Apolipoprotein B

**ALSPAC**: Measured using an automated analyser and derived as a continuous variable in g/L. **BiB, MCS, and MoBa**: Not available.

###### C-Reactive Protein (CRP)

**ALSPAC**: Measured using an automated analyser and derived as a continuous variable in mg/L. **BiB:** Measured using a high-sensitivity immunoturbidimetric assay and derived as a continuous variable in mg/L. **MCS, and MoBa**: Not available.

###### HDL cholesterol

**ALSPAC**: Measured using an automated analyser and derived as a continuous variable in mmol/L. **BiB:** Measured by lipid profile analyzing using nuclear magnetic resonance (NMR) spectroscopy. It is derived as a continuous variable in mmol/L. **MCS, and MoBa**: Not available.

###### Interleukin-6 (IL6)

**ALSPAC**: Measured using enzyme-linked immunosorbent assay (ELISA) and derived as a continuous variable in µU/mL. **BiB, MCS, and MoBa**: Not available.

###### Insulin

**ALSPAC**: Measured using an automated immunoassay analyser and derived as a continuous variable in µU/mL. **BiB:** Insulin levels are measured as part of gestational diabetes (GDM) screening. It is derived as a continuous variable in µU/mL. **MCS and MoBa**: Not available

###### LDL cholesterol

**ALSPAC**: Measured using an automated analyser and derived as a continuous variable in mmol/L. **BiB:** Measured by lipid profile analyzing using nuclear magnetic resonance (NMR) spectroscopy. It is derived as a continuous variable in mmol/L. **MCS and MoBa**: Not available

###### Total cholesterol

**ALSPAC**: Measured using an automated analyser and derived as a continuous variable in mmol/L. **BiB:** Measured by lipid profile analyzing using nuclear magnetic resonance (NMR) spectroscopy. It is derived as a continuous variable in mmol/L. **MCS and MoBa**: Not available

###### Triglycerides

**ALSPAC**: Measured using an automated analyser and derived as a continuous variable in mmol/L. **BiB:** Measured by lipid profile analyzing using nuclear magnetic resonance (NMR) spectroscopy. It is derived as a continuous variable in mmol/L. **MCS and MoBa**: Not available.

##### Blood pressure

**ALSPAC and BiB:** During clinic visits of children at all stages, their blood pressure was measured twice for both systolic and diastolic, and the mean of both the readings were taken. **MCS and MoBa**: not available.

#### Covariates

Child’s sex

Derived as a binary variable and categorized as ‘female’ and ‘male based on sex registered at birth for all the cohorts.

Child’s age

Derived as a continuous variable across all the cohorts. Measured in days, months and years depending on the stage of the child.

Parents’ ethnicity

**ALSPAC, BiB and MCS:** Derived both as a categorical variable (‘Asian’, ‘white’, ‘Black’, and ‘mixed or other’) and binary variable (‘white’ and ‘not white’). **MoBa**: not available.

Parents’ age

Derived as continuous variable for mother’s age at conception (for **ALSPAC and BiB**) and at delivery (for **MCS, and MoBa**), and father’s age during the pregnancy of the mother.

Mother’s parity

**ALSPAC, BiB and MoBa:** Derived as a binary variable and categorized as ‘0’ for nulliparity (i.e. no previous births) and ‘1’ for one or more previous births. **MCS**: not available.

Parents’ education

Defined as described above for SEP as an exposure. When SEP is a covariate, we included the more granular ordered categorical variable rather than the binary (lowest vs anything higher) variable.

Parents’ occupation

Defined as described above for SEP as an exposure. When SEP is a covariate, we included the more granular ordered categorical variable rather than the binary (lowest vs anything higher) variable.

Parents’ other health behaviours (smoking, caffeine, alcohol)

Defined as described above. We used the ‘ever in pregnancy’ variable if the exposure was during pregnancy, and the ‘ever in life’ variable if it was not.

Parents’ health behaviours in previous timepoints (smoking, caffeine, alcohol)

Defined as described above in the Exposures section. Second trimester exposures were adjusted for exposures in the first trimester; third trimester exposures were adjusted for exposures in the first and second trimester; postnatal exposures were adjusted for exposures at any time in pregnancy.

Gestational age at delivery

Derived as a continuous variable, measured in weeks, for all the cohorts. Data were extracted from maternity records where available or self-report.

Birthweight

As described under Outcomes.

Child’s passive smoking before age 2

**ALSPAC: BiB: MoBa and MCS:** derived as a binary variable describing any amount of passive smoke exposure vs none in the first two postnatal years according to parental report. **BiB:** not available.

Child’s caffeine intake before age 2.

**ALSPAC:** derived as a binary variable describing any amount of caffeine from tea, coffee, and cola vs none in the first two postnatal years according to parental report.

**BiB, MoBa and MCS:** not available.

### Genetic risk scores and genetic covariates

##### ALSPAC genotype data

ALSPAC mothers were genotyped using Illumina human660K quad single nucleotide

polymorphism (SNP) chip, and ALSPAC children were genotyped using Illumina HumanHap550 quad genome-wide SNP genotyping platform. Genotype data for both ALSPAC mothers and children were imputed against the Haplotype Reference Consortium v1.1 reference panel, after performing the QC procedure (minor allele frequency (MAF) ≥1%, a call rate ≥95%, in Hardy-Weinberg equilibrium (HWE), correct sex assignment, no evidence of cryptic relatedness, and of European descent).

##### BiB genotype data

BiB mothers and offspring were genotyped at Bristol Bioresource Laboratories, Bristol, UK using four different Illumina arrays: HumanCoreExome12v1.0, HumanCoreExome12v1.1, HumanCoreExome24v1.0 and Infinium Global Screening Array-24 v1.0 (GSA). Genotypes were called with Illumina GenomeStudio. Individuals with high genotype missingness and

SNPs with low call rate were removed using an iterative procedure, resulting in a final sample of individuals with missingness <0.5% and SNPs with call rate >99.5%. Other typically used QC metrics such as deviation from HWE and excess heterozygosity are not appropriate here given the population structure and consanguinity known to be present. Genotype data were imputed to the HRC r1.1 reference panel. Reference genome build version 37 (GRCh37/hg19) was used for genomic positions for BiB genotype data.

##### MoBa genotype data

Genotyping and QC is described in detail in genotyping and QC is described in detail in <https://doi.org/10.1101/2022.06.23.496289> [(7)](https://doi.org/10.1101/2022.06.23.496289).

#### Deriving Genetic Risk Scores (GRS)

Polygenic risk scores (GRS) for alcohol, smoking, and caffeine consumption were constructed using genome-wide significant (P < 5 × 10⁻⁸), LD-independent SNPs identified in large-scale genome-wide association studies (GWAS). SNPs were weighted by their effect estimates as reported in the original GWAS.

The alcohol and smoking GRS were based on summary statistics from (8) which investigated the number of alcoholic drinks consumed per week and four smoking-related traits: age at initiation of regular smoking, ever vs never smoking (“initiation”), cigarettes per day, and smoking cessation (current vs former smoker). For alcohol, consumption was aggregated across beverage types; where categorical data (e.g., 1–5 drinks/week) were used in contributing studies, midpoints were applied (e.g., 2.5 drinks/week). To minimise the influence of outliers, the phenotype was left-anchored at 1 and log-transformed in the original GWAS.

The caffeine GRS was based on a GWAS of the number of coffee cups consumed per day (9).

#### SNPs used to derive the GRS

Alcohol (drinks per week)

| rs705687 | rs62250685 | rs17029090 | rs10978550 | rs2472297 |
| --- | --- | --- | --- | --- |
| rs58107686 | rs74664784 | rs79139602 | rs7074871 | rs12907323 |
| rs12088813 | rs13066454 | rs4699791 | rs17665139 | rs2764771 |
| rs5024204 | rs9838144 | rs13107325 | rs7950166 | rs17177078 |
| rs10753661 | rs2011092 | rs4690727 | rs11030084 | rs378421 |
| rs28680958 | rs60654199 | rs10004020 | rs56030824 | rs113443718 |
| rs823114 | rs6787172 | rs12651313 | rs10750025 | rs62044525 |
| rs77165542 | rs3748034 | rs4916723 | rs1713676 | rs7185555 |
| rs1260326 | rs7682824 | rs12655091 | rs4938230 | rs79616692 |
| rs2178197 | rs11940694 | rs55872084 | rs682011 | rs1104608 |
| rs13383034 | rs35538052 | rs11739827 | rs12795042 | rs4548913 |
| rs1004787 | rs4501255 | rs10085696 | rs10876188 | rs3803800 |
| rs13032049 | rs12499107 | rs6460047 | rs3809162 | rs2854334 |
| rs828867 | rs144198753 | rs10236149 | rs10506274 | rs2532276 |
| rs11692435 | rs1154414 | rs35034355 | rs4842786 | rs10438820 |
| rs13024996 | rs1229984 | rs6951574 | rs500321 | rs9950000 |
| rs72859280 | rs10028756 | rs13250583 | rs1123285 | rs4092465 |
| rs56337305 | rs561222871 | rs1217091 | rs2180870 | rs281379 |
| rs13094887 | rs36052336 | rs28601761 | rs28929474 | rs4815364 |
| rs2165670 | rs55932213 | rs11625650 | rs9607814 |  |

Smoking cessation (regular current or former smoker)

| rs112187834 | rs3025327 | rs6011779 |
| --- | --- | --- |
| rs7617480 | rs10821523 | rs4809543 |
| rs12203592 | rs1611124 | rs6089904 |
| rs707968 | rs7109376 | rs9607805 |
| rs7778443 | rs591143 |  |
| rs1565735 | rs3866543 |  |
| rs60749569 | rs518425 |  |
| rs12378015 | rs145580088 |  |
| rs9409844 | rs56113850 |  |
| rs59586387 | rs117824460 |  |

| Smoking initiation (regular current or non-smoker) | | | |  |  |
| --- | --- | --- | --- | --- | --- |
| rs12130857 | rs10873871 | rs1004787 | rs34399632 | rs10446419 | rs57153235 |
| rs301807 | rs11162019 | rs7598402 | rs74697736 | rs13319205 | rs6437769 |
| rs3820277 | rs1008078 | rs10490159 | rs6756212 | rs3172494 | rs9288999 |
| rs1889571 | rs1935571 | rs1518393 | rs3076896 | rs2526390 | rs6438436 |
| rs10914684 | rs12027999 | rs17616642 | rs16826827 | rs2276825 | rs12053870 |
| rs2637869 | rs45444697 | rs6730325 | rs1445649 | rs2306866 | rs9826984 |
| rs12755632 | rs2901785 | rs2539706 | rs1722666 | rs73831818 | rs2279829 |
| rs951740 | rs147052174 | rs7585579 | rs11678980 | rs1910236 | rs2319545 |
| rs925524 | rs35656245 | rs1863161 | rs12474587 | rs7640107 | rs10935779 |
| rs12022778 | rs12739243 | rs359247 | rs357304 | rs2734390 | rs963354 |
| rs11587399 | rs12563365 | rs62180324 | rs13007361 | rs221988 | rs1714521 |
| rs4912332 | rs876793 | rs6750107 | rs7600835 | rs2196356 | rs1449012 |
| rs1937443 | rs114976176 | rs12714017 | rs6750529 | rs11128203 | rs9850597 |
| rs1022528 | rs62106258 | rs56208390 | rs17229285 | rs62246017 | rs1187820 |
| rs12740789 | rs6731872 | rs11692435 | rs3115418 | rs4543050 | rs16828799 |
| rs80054503 | rs1022376 | rs13392222 | rs62193862 | rs6782116 | rs9841807 |
| rs10789369 | rs61533748 | rs1901477 | rs4674916 | rs13066050 | rs7631379 |
| rs1514176 | rs72790288 | rs11889814 | rs4674993 | rs12633090 | rs4140932 |
| rs2028269 | rs2710634 | rs3811038 | rs11713899 | rs1549979 | rs12642744 |
| rs6874731 | rs62137126 | rs75210106 | rs748832 | rs74664784 | rs59537158 |
| rs6452785 | rs329124 | rs4044321 | rs3218116 | rs12195240 | rs1737329 |
| rs10805858 | rs1385108 | rs2173019 | rs160631 | rs6936160 | rs10272990 |
| rs181508347 | rs1173461 | rs10042827 | rs7743165 | rs12530388 | rs6948707 |
| rs42417 | rs11956866 | rs359431 | rs79180767 | rs3800227 | rs10259715 |
| rs72780746 | rs3909281 | rs1059490 | rs10945141 | rs118202 | rs13237637 |
| rs10060196 | rs3843905 | rs6932350 | rs17554906 | rs73008357 | rs79631993 |
| rs72789626 | rs79476395 | rs1150668 | rs619087 | rs9331343 | rs7809303 |
| rs17165769 | rs6890961 | rs1632941 | rs6568832 | rs10698713 | rs7802996 |
| rs1930371 | rs11594623 | rs7943721 | rs7333559 | rs6497840 | rs17692129 |
| rs2378662 | rs11191269 | rs7929518 | rs1108130 | rs4785187 | rs75919030 |
| rs1927901 | rs28408682 | rs586699 | rs12855717 | rs8050598 | rs2938134 |
| rs4837631 | rs12244388 | rs76460663 | rs12878369 | rs12918191 | rs2587507 |
| rs1759433 | rs111842178 | rs2155646 | rs2145451 | rs9302604 | rs34342129 |
| rs34553878 | rs34970111 | rs78239456 | rs9323328 | rs9936784 | rs4476253 |
| rs7026534 | rs9787523 | rs1713676 | rs1811739 | rs62052916 | rs7505855 |
| rs10858334 | rs11192347 | rs238896 | rs8005334 | rs4788676 | rs8096225 |
| rs10905461 | rs10885480 | rs540860 | rs34940743 | rs61537885 | rs67050670 |
| rs7920501 | rs4752018 | rs1944689 | rs2925128 | rs117657830 | rs2359180 |
| rs1291821 | rs9423279 | rs1834306 | rs1381287 | rs1050847 | rs72898831 |
| rs11258417 | rs6265 | rs1106363 | rs55913542 | rs11642231 | rs8083764 |
| rs7072776 | rs4275621 | rs2010921 | rs1435672 | rs4790874 | rs1373178 |
| rs2796793 | rs62618693 | rs11057005 | rs281296 | rs11078713 | rs62098013 |
| rs1733760 | rs2939756 | rs13906 | rs1435741 | rs28441558 | rs72938304 |
| rs7921378 | rs1381775 | rs4759229 | rs56902655 | rs11651955 | rs11872397 |
| rs7901883 | rs2959084 | rs7969559 | rs2289791 | rs67777803 | rs71367544 |
| rs644740 | rs3740977 | rs7134009 | rs60833441 | rs2344976 | rs76608582 |
| rs61884449 | rs61886926 | rs77215829 | rs62007780 | rs3764351 | rs10853981 |
| rs113230003 | rs9545155 | rs62340589 | rs6993429 | rs6050446 | rs11076962 |
| rs1389171 | rs112913817 | rs12517438 | rs6986430 | rs6058782 | rs7192140 |
| rs55944129 | rs10233018 | rs35375873 | rs9987376 | rs1555445 | rs9922607 |
| rs58400863 | rs10953957 | rs986714 | rs290601 | rs6073075 | rs9941217 |
| rs7657022 | rs77283305 | rs71592686 | rs3847244 | rs910912 | rs7188873 |
| rs55900829 | rs10279261 | rs1030015 | rs11791671 | rs6011779 | rs11611651 |
| rs112725451 | rs1561112 | rs4727189 | rs7024924 | rs3810496 | rs17197663 |
| rs1160685 | rs2952251 | rs76841737 | rs6474609 | rs4818005 | rs4264267 |
| rs1435479 | rs4326350 | rs11768481 | rs1931431 | rs139896 | rs61959481 |
| rs3934797 | rs11780471 | rs1799068 | rs7867822 | rs4822102 | rs3098272 |
| rs71602617 | rs11783093 | rs13437771 | rs10966092 | rs9627272 | rs9538162 |
| rs7696257 | rs1565735 | rs11766326 | rs10969352 | rs1413119 | rs1772572 |
| rs13109980 | rs7836565 | rs6968380 | rs4877285 | rs56367474 | rs75674569 |
| rs1116690 | rs13261666 | rs8103660 | rs4310804 | rs55786907 | rs1109480 |
| rs13110073 | rs3850736 | rs117734003 | rs8027457 | rs4886207 | rs12442563 |
| rs28717373 | rs2063976 | rs1126757 | rs1139897 | rs9540731 | rs72836318 |

Age of smoking initiation (age 1^st^ regularly smoked)

rs72853300

rs12611472

rs7559982

rs11915747

rs13136239

rs2471711

rs624833

rs7682598

rs1403174

rs11780471

| Cigarettes per day | |  |
| --- | --- | --- |
| rs11264100 | rs10519203 | rs75596189 |
| rs2072659 | rs28438420 | rs3025383 |
| rs34973462 | rs72740955 | rs7951365 |
| rs7599488 | rs146009840 | rs10742683 |
| rs78408772 | rs28681284 | rs113001570 |
| rs10204824 | rs8040868 | rs7125588 |
| rs2084533 | rs3743063 | rs11846838 |
| rs7431710 | rs182317 | rs1115019 |
| rs2236951 | rs1592485 | rs632811 |
| rs699165 | rs12924872 | rs4886550 |
| rs28813180 | rs258321 | rs12438181 |
| rs1024323 | rs4144686 | rs56113850 |
| rs11940255 | rs4485470 | rs8192726 |
| rs10454798 | rs59208569 | rs117824460 |
| rs7766641 | rs143200968 | rs6078373 |
| rs215600 | rs73229090 | rs1737894 |
| rs62447179 | rs13253502 | rs2273500 |
| rs2741351 | rs4236926 | rs7281463 |
| rs790564 |  |  |

Coffee consumption

rs1260326

rs1481012

rs6968554

rs7800944

rs17685

rs6265

rs2472297

rs9902453

#### F-statistics

F-statistics for the association between each genetic risk score (GRS) and corresponding health behaviour. F<10 (dark pink) indicates a weak instrument.

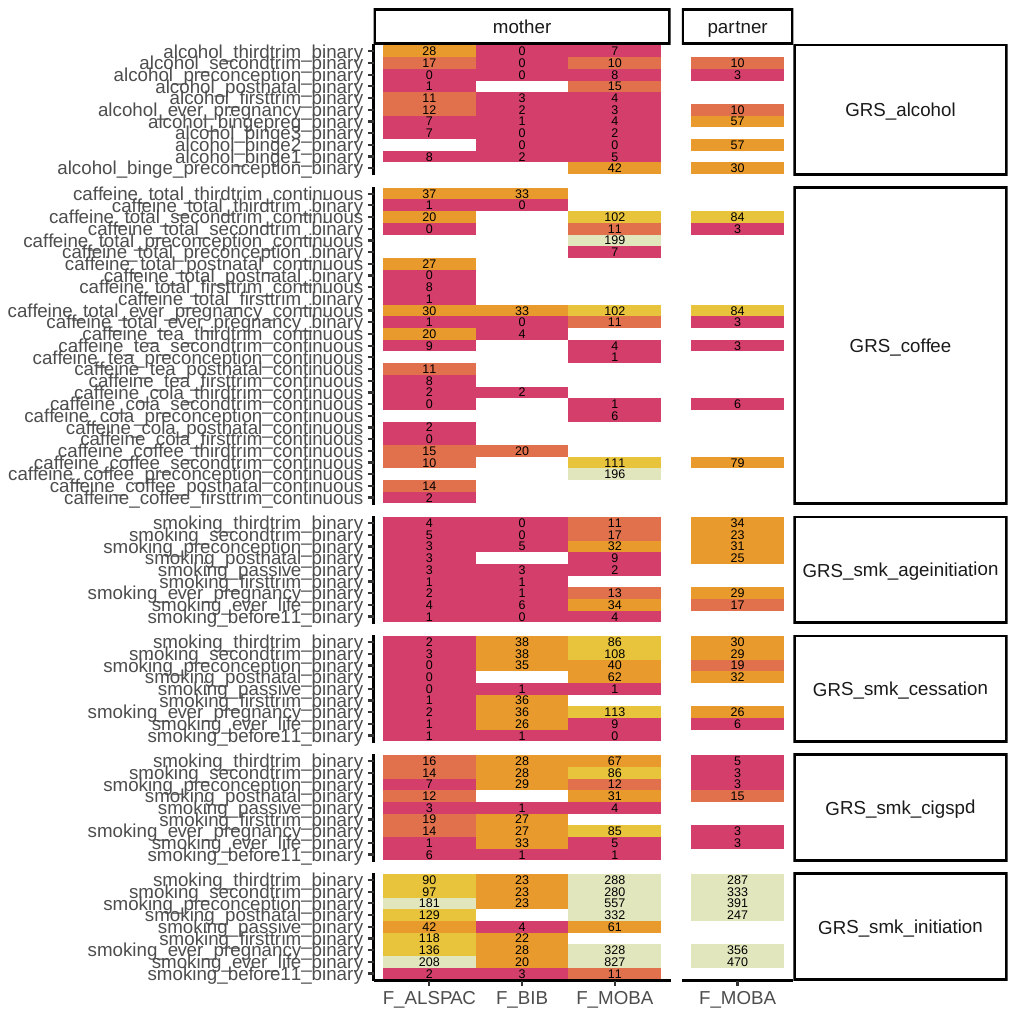

### References

1.	Boyd A, Golding J, Macleod J, Lawlor DA, Fraser A, Henderson J, et al. Cohort Profile: The ‘Children of the 90s’—the index offspring of the Avon Longitudinal Study of Parents and Children. Int J Epidemiol. 2013 Feb;42(1):111–27.

2.	Fraser A, Macdonald-Wallis C, Tilling K, Boyd A, Golding J, Davey Smith G, et al. Cohort Profile: The Avon Longitudinal Study of Parents and Children: ALSPAC mothers cohort. Int J Epidemiol. 2013 Feb 1;42(1):97–110.

3.	Wright J, Small N, Raynor P, Tuffnell D, Bhopal R, Cameron N, et al. Cohort Profile: The Born in Bradford multi-ethnic family cohort study. Int J Epidemiol. 2013 Aug 1;42(4):978–91.

4.	Connelly R, Platt L. Cohort Profile: UK Millennium Cohort Study (MCS). Int J Epidemiol. 2014 Dec 1;43(6):1719–25.

5.	Magnus P, Birke C, Vejrup K, Haugan A, Alsaker E, Daltveit AK, et al. Cohort Profile Update: The Norwegian Mother and Child Cohort Study (MoBa). Int J Epidemiol. 2016 Apr;45(2):382–8.

6.	Rønningen KS, Paltiel L, Meltzer HM, Nordhagen R, Lie KK, Hovengen R, et al. The biobank of the Norwegian Mother and Child Cohort Study: a resource for the next 100 years. Eur J Epidemiol. 2006;21(8):619–25.

7.	Corfield EC, Shadrin AA, Frei O, Rahman Z, Lin A, Athanasiu L, et al. The Norwegian Mother, Father, and Child cohort study (MoBa) genotyping data resource: MoBaPsychGen pipeline v.1 [Internet]. 2022 [cited 2025 Aug 10]. Available from: http://biorxiv.org/lookup/doi/10.1101/2022.06.23.496289

8.	Liu M, 23andMe Research Team, HUNT All-In Psychiatry, Jiang Y, Wedow R, Li Y, et al. Association studies of up to 1.2 million individuals yield new insights into the genetic etiology of tobacco and alcohol use. Nat Genet. 2019 Feb;51(2):237–44.

9.	The Coffee and Caffeine Genetics Consortium, Cornelis MC, Byrne EM, Esko T, Nalls MA, Ganna A, et al. Genome-wide meta-analysis identifies six novel loci associated with habitual coffee consumption. Mol Psychiatry. 2015 May;20(5):647–56.
