## Supplementary File S2: Data Availability for "Exploring parental prenatal influences on child health: a multicohort study and data visualisation tool"

These figures show a summary of data available on exposures (**Figure 1**) and outcomes (**Figure 2**) across all four cohorts.

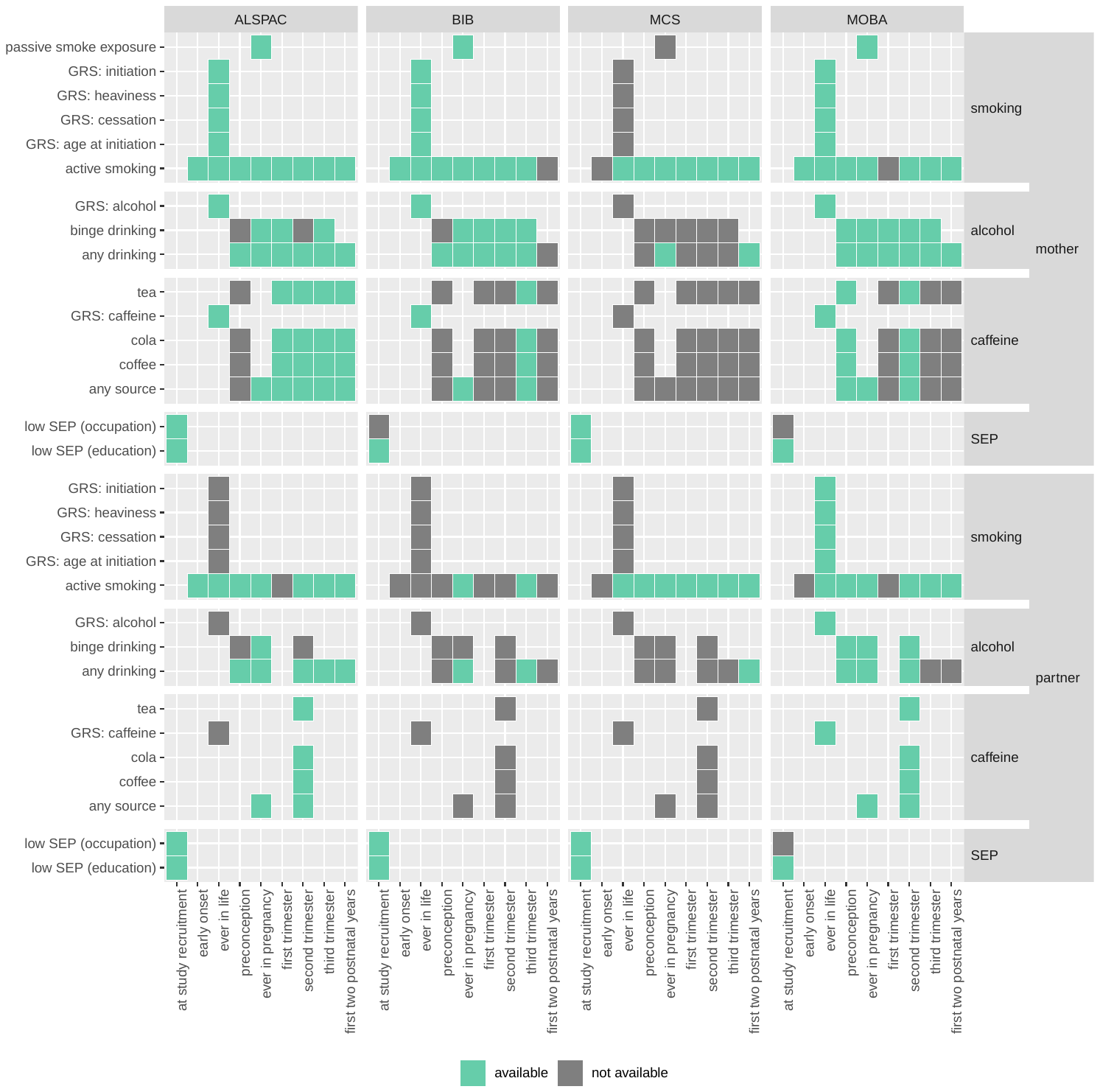

***Figure 1.*** *Availability of data on exposures (light grey, i.e. unfilled squares, mean the data are either not available for any cohort, or the category is not applicable, for example, GRS in the third trimester).*

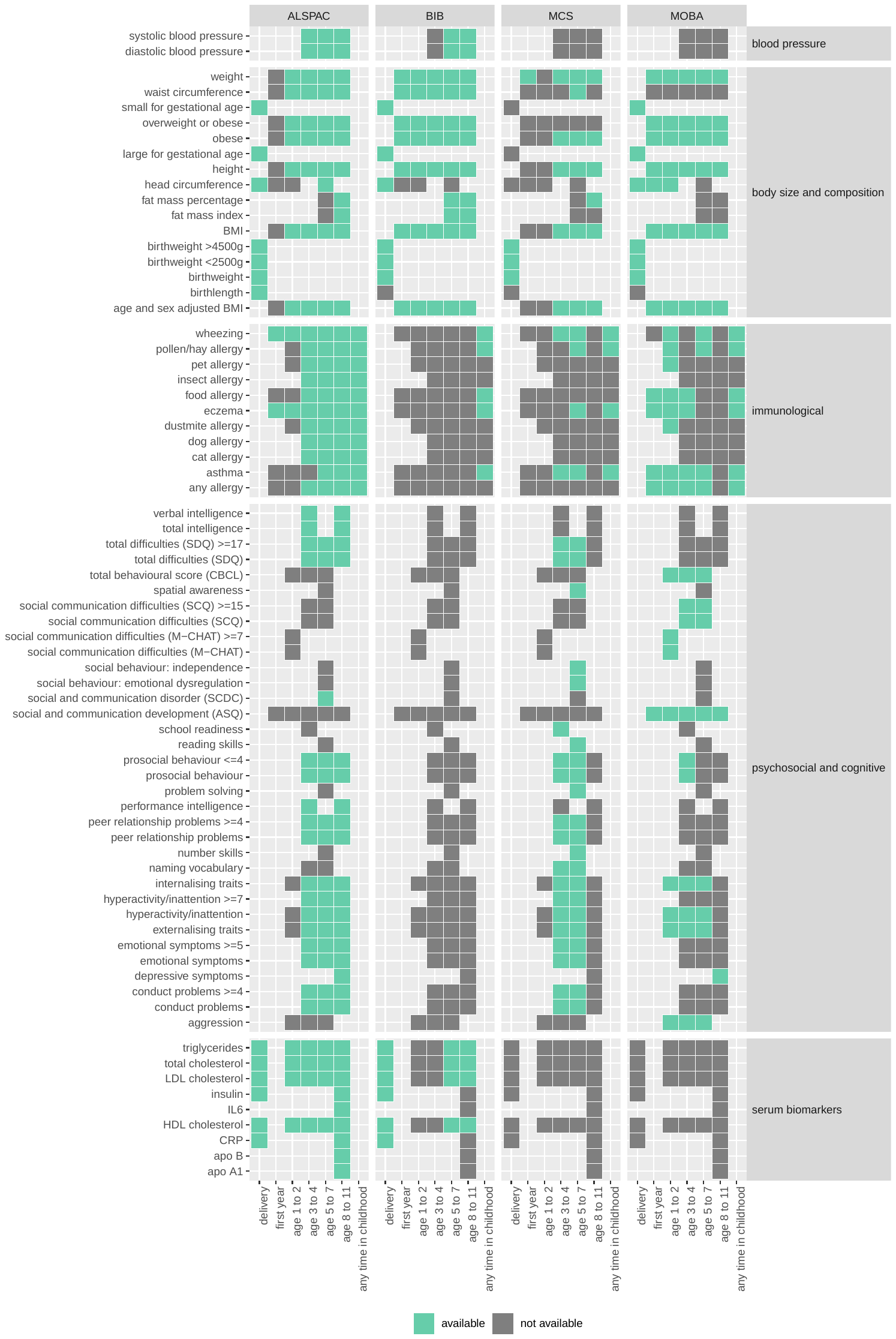

***Figure 2.*** *Availability of data on outcomes (light grey, i.e. unfilled squares, mean the data are either not available for any cohort, or the category is not applicable, for example, birthweight at age 8 to 11).*
