## Supplementary File S3: Statistical analysis details for "Exploring parental prenatal influences on child health: a multicohort study and data visualisation tool"

Selecting covariates

We selected covariates for inclusion in our models based on scientific understanding, and then applied Directed Acyclic Graphs (DAGs) to check the model assumptions. Examples are shown in Figures 1 to X.

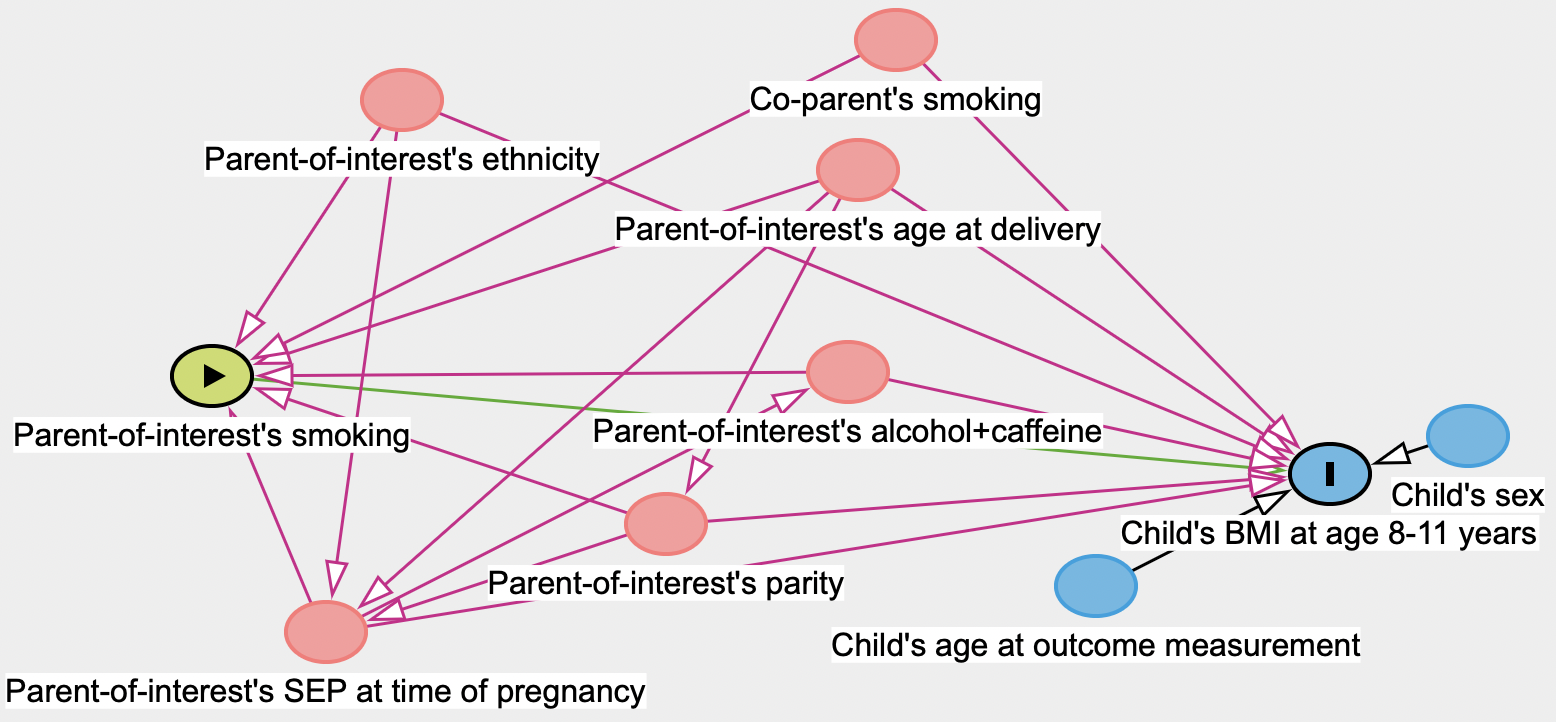

***Figure 1.*** *DAG for models where the exposure is a health behaviour (smoking shown as an example here). Potential confounders and their biasing relationships are shown in pink; green shows the exposure and its relationship with the outcome; blue shows the outcome or ancestors of the outcome. BMI at age 8-11 is given as an example outcome.*

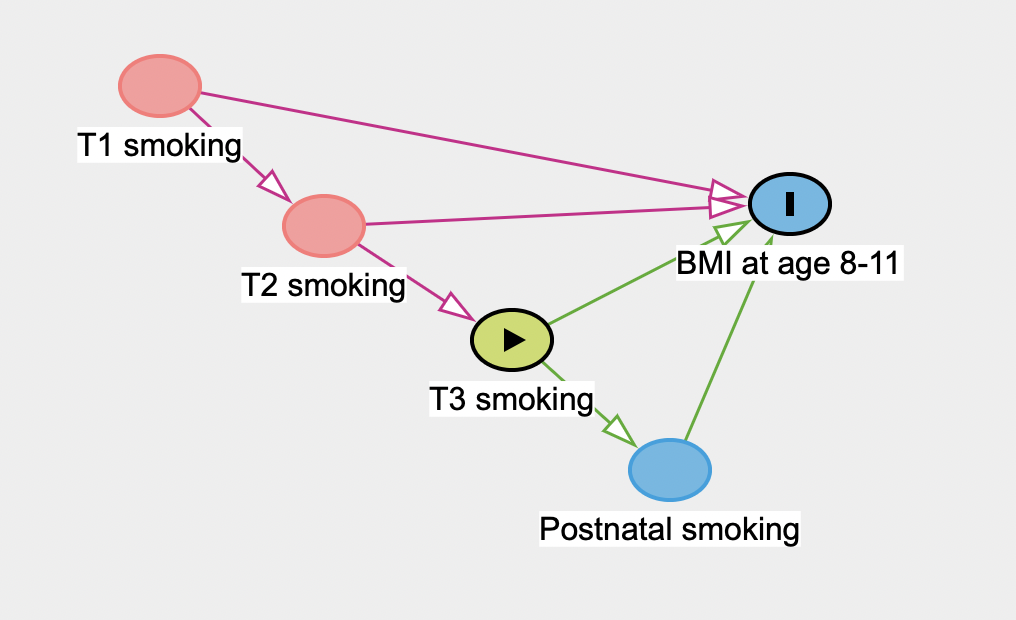

***Figure 2.*** *DAG for models where the exposure is a health behaviour that occurs in a specific time during (trimester 1, 2, 3) or after pregnancy (smoking shown as an example here). Potential confounders and their biasing relationships are shown in pink; green shows the exposure and its relationship with the outcome; blue shows the outcome or ancestors of the outcome. BMI at age 8-11 is given as an example outcome. For simplicity, other confounders are not plotted.*

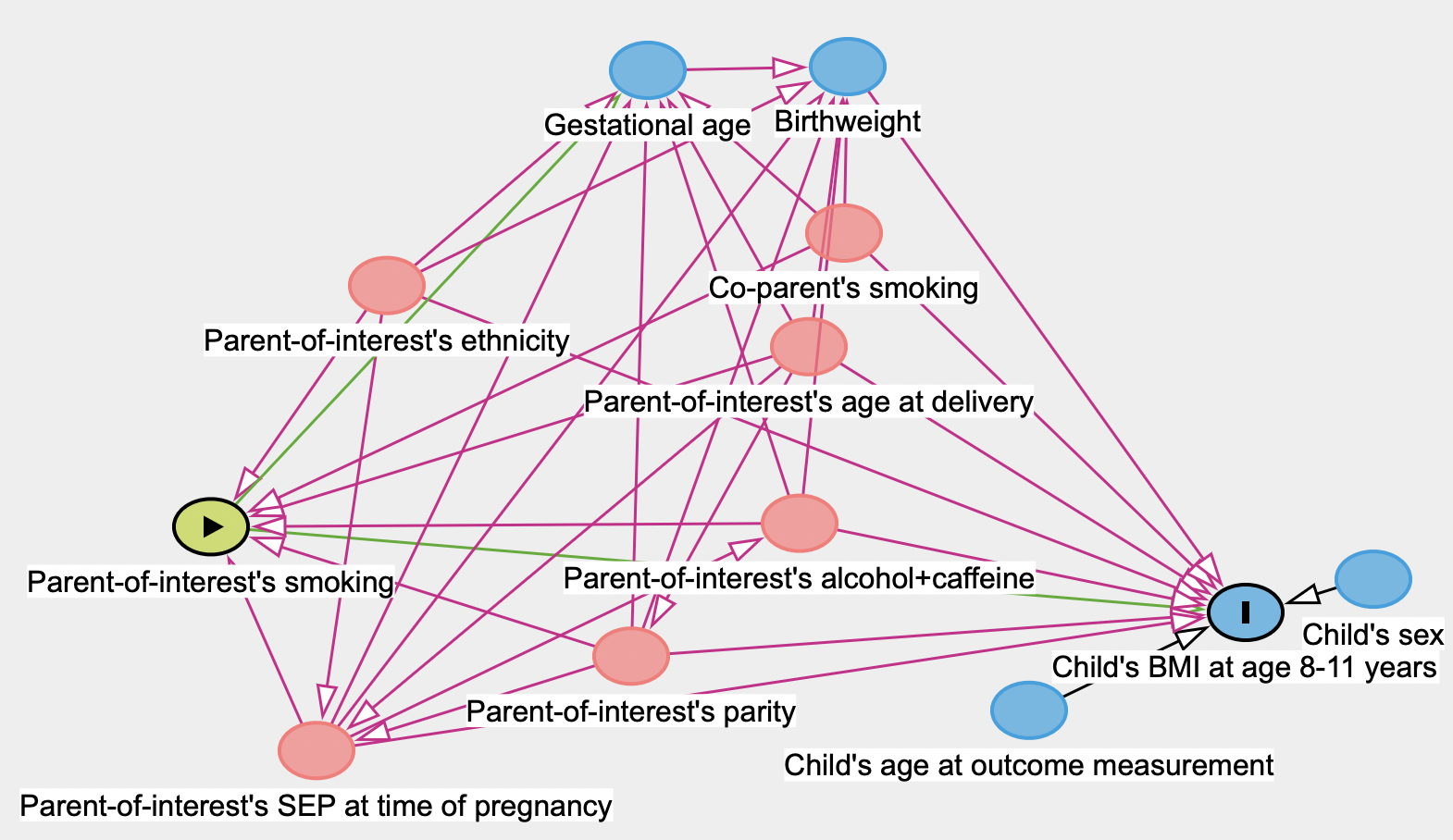

***Figure 3.*** *DAG for models where the exposure is a health behaviour (smoking shown as an example here) and the model includes potential mediators (gestational age and birthweight). Potential confounders and their biasing relationships are shown in pink; green shows the exposure and its relationship with the outcome; blue shows the outcome or ancestors of the outcome. In this case, birthweight and gestational age are mediators sitting on the causal pathway between the exposure (smoking) and the outcome (BMI at age 8-11 is given as an example outcome).*

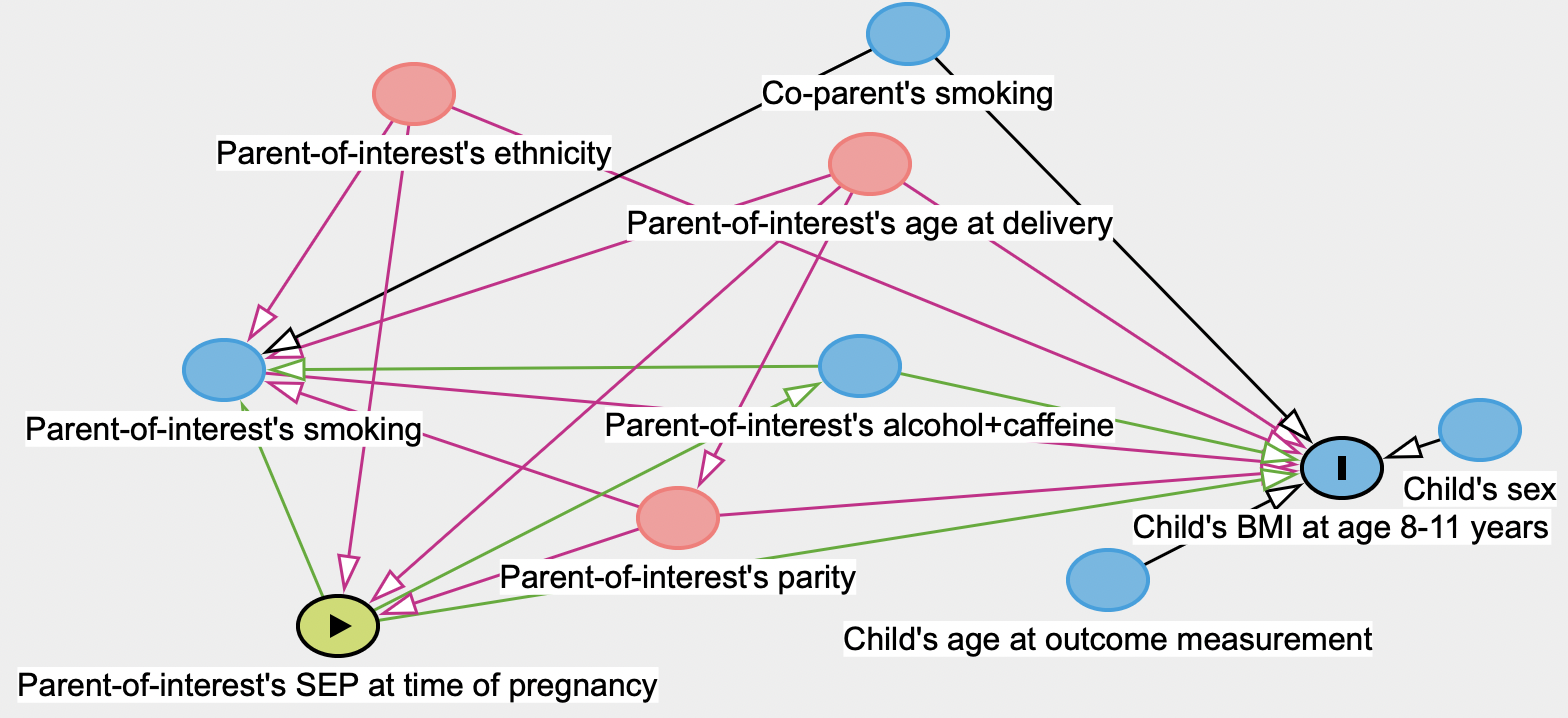

***Figure 4.*** *DAG for models where the exposure is SEP. This is the same DAG as depicted in Figure 1, with the exposure swapped from smoking to SEP. Potential confounders and their biasing relationships are shown in pink; green shows the exposure and its relationship with the outcome; blue shows the outcome or ancestors of the outcome (BMI at age 8-11 is given as an example outcome).*

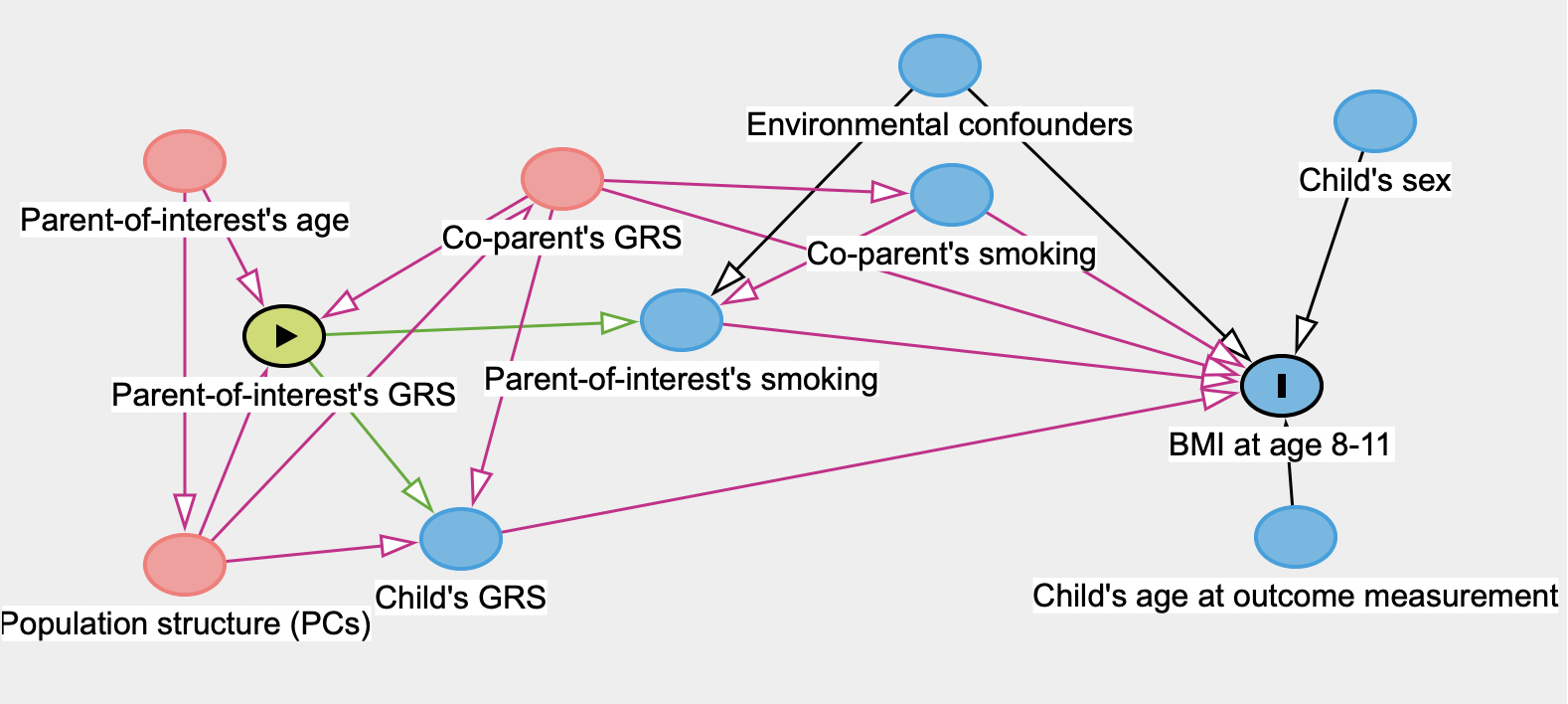

***Figure 5.*** *DAG for models where the exposure is a GRS. Within a mendelian randomization (MR) framework, the GRS acts as an instrument for the exposure (smoking in this example). Potential confounders and their biasing relationships are shown in pink; green shows the exposure and its relationship with the exposure it is instrumenting; blue shows the outcome or ancestors of the outcome (BMI at age 8-11 is given as an example outcome). The GRS is not affected by environmental confounders. Child’s GRS could act as a mediator.*

Models and rationales

All models are numbered and lettered consistently across different exposure types (i.e. health behaviour, SEP, GRS) according to the rationale for the model:

| Model number/letter, name, and rationale | **a. Without adjustment for the exposure in the other partner:**  ***to explore the total effect*** | **b. With adjustment for the exposure in the other partner:**  ***to allow us to estimate the parent-specific effect independent of the effect of co-parent’s exposure*** |
| --- | --- | --- |
| **1. Minimal adjustment:**  ***to explore associations with maximum power and minimum selection, while still controlling for important covariates.*** | Model 1a | Model 1b |
| **2. Standard adjustment for non time-specific exposures:**  ***to control for potential confounders*** | Model 2a | Model 2b |
| **3. Standard adjustment for time-specific exposure:**  ***to control for correlation with previous timepoints in pregnancy by adjusting for the standard adjustment set, plus the exposure in the parent-of-interest at previous timepoints*** | Model 3a | Model 3b |
| **4. Standard adjustment plus potential mediators:**  ***to explore the direct effect of the exposure on the outcome by controlling for factors that (most likely) occur after the exposure and could sit on the causal pathway to the outcome.*** | Model 4a | Model 4b |

Variables included in each model are described in Table 1 of the main text of this paper, and summarised below.

***Self-reported health behaviours:*** we used four different adjustment sets: Model 1 minimal adjustment (parent-of-interest’s ethnicity, child’s sex, and child’s age at outcome measurement); Model 2 standard adjustment (additional adjustment for parent-of-interest’s age at conception/delivery, education, occupation, smoking (where not the main exposure), alcohol consumption (where not the main exposure), and parity (where the mother is the parent-of-interest); Model 3 *standard adjustment plus adjustment for the exposure in previous time points* (this model was run when the main exposure occurred in the second trimester (adjusted for first), third trimester (adjusted for 1^st^ and 2^nd^), or postnatally (adjusted for ever_in_pregnancy)); and Model 4 *standard adjustment plus adjustment for potential mediators* (i.e. gestational age at delivery, birthweight). Models were run without (set a) and with (set b) adjustment for the equivalent health behaviour in the co-parent.

***Genetic risk scores:*** we used two adjustment sets, which we name 2 and 4 for consistency with the observational analyses: Model 2 *standard adjustment* (parent-of-interest’s age at DNA collection, 10 ancestry-informative principal components (PCs), child’s sex and age at outcome measurement); Model 4 *standard adjustment plus adjustment for potential mediators* (standard adjustment plus additional adjustment for the equivalent GRS in the child and 10 PCs derived from the child’s genotype data). Models were run without (set a) and with (set b) adjustment for the equivalent GRS + 10 ancestry-informed principal components (PCs) in the co-parent.

***Socioeconomic position:*** we used three adjustment sets: Model 1 *minimal adjustment* (parent-of-interest’s ethnicity, child’s sex, and child’s age); Model 2 *standard adjustment* (additional adjustment for parent-of-interest’s age at delivery/conception and parity (where the mother is the parent-of-interest); and Model 4 *standard adjustment plus adjustment for potential mediators* (i.e. gestational age at delivery, birthweight). Models were run without (set a) and with (set b) adjustment for the equivalent measure of SEP in the co-parent.

Processing of cohort results

Analyses were run in each individual cohort. To uphold the privacy of participants we did not include analyses where the total sample size was <20, or any binary exposure or outcome group had n<5. We also removed results with an implausible effect estimate, which may have arisen due to the model failing to converge. We set lenient absolute thresholds to identify these outliers: 10 where the effect estimate was a standardised mean difference (i.e. resulting from a linear regression with a continuous outcome variable); and 3 (equating to an odds ratio of ~20 or 2*10^-9^) where the effect estimate was a log odds ratio (i.e. resulting from a logistic regression with a binary outcome variable).

Processing of final results

After meta-analysing across cohorts where possible, we had generated results from 594,326 analyses, which included a mix of regression results from single cohorts (where only one cohort had the required data for that analysis) and meta-analysis results (where two or more cohorts had the required data). P-values for each model were then corrected for multiple testing using a false discovery rate (FDR) of 5%. To aid visual comparisons between binary and continuous outcome results, we calculated Cohen’s D (standardised mean difference) which is a unitless measure of effect. Because we had already standardised continuous outcomes (to Z scores), no adjustment to the resulting effect estimate from linear regression was needed, but for binary outcomes, we converted the logistic regression effect estimates to Cohen’s D by multiplying the log odds ratio by 0.5513 as described elsewhere (1).

Causal inference approaches and triangulation

EPoCH analyses were designed to enable us to garner evidence from multiple causal inference approaches to test the causal hypothesis for health behaviours ***during pregnancy*** affecting child health via an *in-utero* mechanism. The approaches we used were:

***Multivariable regression adjusting for potential confounders:***

**Rationale:** Adjusting for measured potential confounders in multivariable regression models can help to reduce bias from confounding. **Assumptions:** All confounders are measured well and included in the model so that there is no residual confounding. MVR also assumes: no selection of participants that induces spurious associations; any misclassification of exposure is not related to the outcome (and vice versa); misclassification of covariates are not systematically related to the exposure or outcome. **A priori sources of bias:** Measurement error in exposure, measurement error in outcome, unmeasured (or residual) confounding, misclassification of exposure/outcome related to the outcome/exposure, differential missing data between exposure levels due to loss to follow-up. **Expected direction of bias if affected by residual confounding:** Estimates are most likely biased away from the null (i.e., if there is a true effect, the magnitude is likely to be exaggerated in these results). **Assessing causal evidence:**  If the expected direction of bias is correct, estimates will move towards the null as more variables are included in the model (i.e., as confounding is reduced). However, sometimes adjusting for variables that are on the causal pathway between an exposure and an outcome (i.e. mediators) can introduce collider bias that can bias estimates away from the null, so estimates for models adjusted for mediators (models 4a and 4b) may not fit this pattern.

***Instrumenting parental health behaviours with GRS in a Mendelian randomization (MR) framework:***

**Rationale:** A genetic risk score (GRS) contains several genetic variants that have been shown to robustly relate to the exposure of interest. Genetic variation is fixed at conception and therefore cannot be modified by exposures occurring after conception. Therefore, in MR a GRS is used to ‘instrument’ an exposure to assess its potential causal effect on an outcome, while overcoming some types of confounding. MR is analogous to a randomised control trial, whereby alleles are randomly assorted at conception, resulting in groups of individuals who differ with respect to genetic variation and between which environmental confounders are equally distributed. **Assumptions:** There are three main assumptions: (1) Relevance: the genetic variant is robustly associated with the exposure of interest; (2) Independence: there are no unmeasured confounders of the association between the exposure and outcome; (3) Exclusion restriction: the GRS affects the outcome *only* via its effect on the exposure. **A priori sources of bias:** Violation of any of the three assumptions above; population stratification. **Expected direction of bias:** Violation of the relevance assumption would introduce ‘weak instrument bias’, which would bias estimates towards the null. Given that parents transmit 50% of their genetic information to their children, some of the estimated effect may be driven by genetic transmission rather than a causal effect of the exposure. Therefore, without adjustment for child’s GRS (i.e. Model 2a/2b), the effect estimate may be overestimated away from the null (2). However, adjusting for child’s GRS (Model 4a) may introduce collider bias that would also push the effect estimate away from the null (and potentially in a different direction). Additionally adjusting for the other parent’s GRS (i.e Model 4b) may help to avoid collider bias. However, because only one cohort has genetic data for partners, and sample sizes are smaller, full adjustment results in small samples and low power analyses. Therefore, when triangulating evidence, we use Model 2a, but heed caution that these estimates may be biased. **Assessing causal evidence:** Generally, the power for genetic analyses is lower than for observational analyses because the sample size with genetic data is lower and/or the GRS only partially explains the variance in the exposure. Therefore, confidence intervals are likely to be wide. However, where estimates are large and/or confidence intervals do not cross the null, there is some evidence of a causal effect.

***Employing co-parent’s exposure as a negative control to account for shared familial effects:***

**Rationale:** Parents typically share a household and environmental exposures, and often exhibit similar behaviours and demographic profiles due to assortative mating. Biological parents also share 50% of their genomes with their children. Therefore, exposure variables for each parent are likely to be correlated with each other, and with a set of (unmeasured and measured) ‘familial’ confounders. Accordingly, when estimating the effect of one parent’s exposure, independent of the co-parent’s exposure, it is important to adjust for the co-parent’s exposure to help account for this shared confounding. Moreover, the use of the co-parent’s exposure as a negative control rests on the idea that, while both parents may share familial confounders, they differ in terms of their capacity to influence the fetus via a direct intrauterine mechanism. Specifically, maternal exposures during pregnancy may plausibly have a biological intrauterine effect on the developing fetus. In contrast, partner exposures during this period are unlikely to affect the fetus through the same direct mechanism. Instead, any potential influence would likely occur pre-conception (e.g., via sperm epigenetics or germline mutations) or indirectly (e.g., through effects on the mother's behaviour or stress during pregnancy). Therefore, if both maternal and partner exposures show similar associations with offspring outcomes—even after mutual adjustment—it suggests that the maternal association may be due to **shared familial confounding** rather than a causal intrauterine effect. Conversely, if the maternal effect is stronger and persists after adjustment for the paternal exposure, this may support a **true causal maternal effect. It is important to note that this logic does not apply in reverse**—maternal exposures cannot serve as a negative control for paternal effects, because maternal exposures during pregnancy *can* plausibly have **direct biological effects on the fetus**. Therefore, we interpret the use of partner exposures as a negative control as a tool to interrogate potential *maternal* intrauterine effects only. **Assumptions:** The key sources of bias (specific confounders, misclassification bias and other biases) are the same (shared) for both parents’ exposures; there is no real causal effect of the partner’s exposure on the outcome *operating via the same mechanism as the maternal effect*. The maternal and partner exposures are measured on the same/similar scales. **A priori sources of bias:** The sources of bias between the maternal and partner exposure are different (i.e. they aren’t shared). There is a real causal effect of the partner’s exposure on the outcome that *operates via the same (or a similar) mechanism as the mother’s effect.* **Expected direction of bias if confounded by SEP:** Prior to mutual adjustment, estimates are most likely biased *away from* the null (i.e., if there is a true effect, the magnitude is likely to be exaggerated in these results). **Assessing causal evidence:** Where the estimated effect of the mother’s exposure is greater than the partner’s, even after mutual adjustment, this suggests a potential causal effect of the mother’s exposure on the outcome. Where mutually-adjusted estimates move to the null, with confidence intervals that cross the null, this suggests that the association is more likely to be explained by (residual) shared familial confounding than a true causal effect.

***Employing postnatal exposure as a negative control for pregnancy exposures:***

**Rationale:** the biological mechanisms that explain any effect of exposures during pregnancy are likely to be different from those that explain effects of exposures postnatally. For example, while exposures during pregnancy could have intrauterine effects on the developing fetus, post-pregnancy exposures may affect child health via parental behaviours (or a direct effect in the case of passive smoke exposure), or may have a non-causal association with offspring health due to their correlation with other confounding factors. Therefore, the postnatal exposure, which cannot plausibly influence the offspring outcome in pregnancy, can be used as a negative control for the pregnancy exposure. To account for the within-person correlation in health behaviours over time, the postnatal exposure model is adjusted for the same exposure during pregnancy. **Assumptions:** No residual confounding; no selection of participants that induces spurious associations; any misclassification of exposure is not related to the outcome (and vice versa); misclassification of covariates are not systematically related to the exposure or outcome. In addition, these analyses also assume that exposures are measured on the same/similar scales at all included time points. **A priori sources of bias:** Measurement error in exposure; measurement error in outcome; unmeasured (or residual) confounding; misclassification of exposure/outcome related to the outcome/exposure; differential missing data between exposure levels due to loss to follow-up; exposures measured on different scales at different time points. **Assessing causal evidence:** If there is a causal effect of exposure during pregnancy, we would expect estimates of the effect of the exposure in pregnancy to be greater (further from the null) than estimates of the effect of the exposure post-pregnancy (after adjustment for the exposure in pregnancy).

***Exploring dose-response relationships:***

**Rationale:** if there is a causal effect of an exposure on an outcome, greater levels of the exposure should be associated with a stronger effect. **Assumptions:** No residual confounding, no selection of participants that induces spurious associations, any misclassification of exposure is not related to the outcome (and vice versa), misclassification of covariates are not systematically related to the exposure or outcome. **A priori sources of bias:** Measurement error in exposure, measurement error in outcome, unmeasured (or residual) confounding, misclassification of exposure/outcome related to the outcome/exposure, differential missing data between exposure levels due to loss to follow-up. **Assessing causal evidence:** If there is a dose-response effect, effect estimates should move away from the null as the exposure level increases.

Reproducibility and code checks

Code to prepare the data in each cohort was written (in R version 4.2.2 or higher) by one author (GCS or KEE) and checked by the other. KEE wrote and GCS checked the PLINK code for generating GRS. GCS wrote and KEE checked the analysis R code. Analyses were run multiple times, allowing issues to be identified and corrected. EPoCH Explorer was initially coded in RShiny by GCS, optimised by EH, and further developed by GCS. All R code is available on GitHub: <https://github.com/gcsharp/EPoCH_analysis>. Individual level data from each cohort cannot be shared under the terms of each cohort’s data sharing agreement (but is available to researchers directly from the cohorts themselves), but all summary statistics from our analyses are available via EPoCH Explorer. The RShiny code for EPoCH Explorer is also available on GitHub: <https://github.com/gcsharp/EPoCH_results_app>. This paper is written in accordance with the STROBE guidelines for reporting of observational studies (checklist in **Supplementary File 4**).

**References**

1.	Anzures‐Cabrera J, Sarpatwari A, Higgins JP. Expressing findings from meta‐analyses of continuous outcomes in terms of risks. Stat Med. 2011 Nov 10;30(25):2967–85.

2.	Lawlor D, Richmond R, Warrington N, McMahon G, Davey Smith G, Bowden J, et al. Using Mendelian randomization to determine causal effects of maternal pregnancy (intrauterine) exposures on offspring outcomes: Sources of bias and methods for assessing them. Wellcome Open Res. 2017 Feb 14;2:11.
