## Supplementary File S5: Supplementary Results for "Exploring parental prenatal influences on child health: a multicohort study and data visualisation tool"

Sample sizes

**Table 1** summarises the number of cohorts and sample sizes contributing to analyses for each model.

***Table 1.*** *Summary of the number of cohorts and sample sizes contributing to analyses*

| Model name | Model description | Parent of interest | Number of analyses | Number of analyses involving more than one cohort | Percentage of analyses involving more than one cohort | Mean sample size | Minimum sample size | Maximum sample size |
| --- | --- | --- | --- | --- | --- | --- | --- | --- |
| Model 1a | Minimal adjustment - without co-parent's exposure | mother | 17250 | 5283 | 31 | 28937 | 93 | 232139 |
|  |  | partner | 10225 | 3290 | 32 | 25098 | 207 | 172635 |
| Model 1b | Minimal adjustment - with co-parent's exposure | mother | 17019 | 4599 | 27 | 19097 | 62 | 170358 |
|  |  | partner | 10180 | 3217 | 32 | 23772 | 70 | 167217 |
| Model 2a | Standard adjustment - without co-parent's exposure | mother | 17153 | 5155 | 30 | 22644 | 83 | 184324 |
|  |  | partner | 10181 | 3290 | 32 | 14389 | 196 | 135285 |
| Model 2b | Standard adjustment - with co-parent's exposure | mother | 16931 | 4543 | 27 | 15376 | 50 | 136842 |
|  |  | partner | 10124 | 3217 | 32 | 13703 | 68 | 129860 |
| Model 3a | Standard adjustment - without co-parent's exposure - plus exposure at previous timepoint(s) | mother | 9586 | 2824 | 29 | 18316 | 55 | 154726 |
|  |  | partner | 6537 | 2137 | 33 | 12927 | 192 | 74710 |
| Model 3b | Standard adjustment - with co-parent's exposure - plus exposure at previous timepoint(s) | mother | 9428 | 2332 | 25 | 13035 | 35 | 119050 |
|  |  | partner | 6496 | 2105 | 32 | 12202 | 68 | 68822 |
| Model 4a | Standard adjustment - without co-parent's exposure - plus potential mediators | mother | 16960 | 5025 | 30 | 22000 | 81 | 178534 |
|  |  | partner | 10078 | 3195 | 32 | 14004 | 185 | 134200 |
| Model 4b | Standard adjustment - with co-parent's exposure - plus potential mediators | mother | 16744 | 4428 | 26 | 14954 | 50 | 136003 |
|  |  | partner | 10024 | 3136 | 31 | 13352 | 68 | 128797 |
| Model 2a: GRS | Standard adjustment - without co-parent's GRS | mother | 1254 | 510 | 41 | 18934 | 221 | 76990 |
|  |  | partner | 558 | 0 | 0 | 23258 | 1310 | 43681 |
| Model 2b: GRS | Standard adjustment - with co-parent's GRS | mother | 558 | 0 | 0 | 20551 | 1159 | 38354 |
|  |  | partner | 558 | 0 | 0 | 20551 | 1159 | 38354 |
| Model 4a: GRS | Standard adjustment - without co-parent's GRS - plus child's GRS | mother | 1254 | 510 | 41 | 14663 | 182 | 56432 |
|  |  | partner | 558 | 0 | 0 | 18783 | 1058 | 34274 |
| Model 4b: GRS | Standard adjustment - with co-parent's GRS - plus child's GRS | mother | 558 | 0 | 0 | 17022 | 957 | 31021 |
|  |  | partner | 558 | 0 | 0 | 17022 | 957 | 31021 |

Sample characteristics

**Figure 1** shows the Pearson correlation coefficients for associations between measures of socioeconomic position (here we used the ordinal variable, where higher values = higher SEP) and the studied health behaviours, across parents and cohorts.

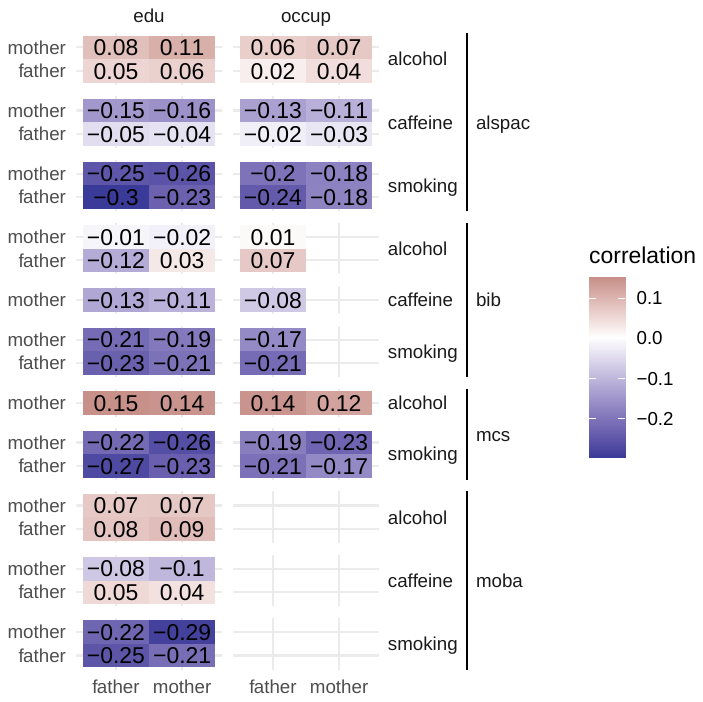

***Figure 1.*** *Correlations between low SEP (education ‘edu’ and occupation ‘occup’) and health behaviours (ever during pregnancy binary for smoking and alcohol, continuous for caffeine).*

General trends

*Methods: To create the following plots, we calculated the proportion of standardised estimates (Cohen’s D) < or > 0.2 with and without FDR-adjusted P-values <0.05, yielding four evidence categories. We then took the direction of the effect estimate and reversed it if higher levels of the outcome were considered beneficial for offspring health. We then calculated the number of associations in each grouping, depending on how the plot is broken down (e.g. direction + exposure class + outcome class + exposure time in Figure 1) [X], and the number of those associations in each of the four evidence categories [Y]. Finally, we calculated the percentage (Y/X *100) and multiplied the result by the direction (-1 or 1).*

**Figure 2** shows the distribution of strength of evidence across exposure classes, exposure times, parent-of-interest, and outcome classes for our main model, model 2a. This shows that associations with the strongest evidence (i.e. absolute Cohen’s D effect estimate >0.2 and FDR-adjusted P-value <0.05; dark green in the figure) exist mostly when the exposure is low SEP or smoking, and the outcome is psychosocial/cognitive or body size/composition. It also shows that, for partners, alcohol consumption tends to be is associated with better child psychosocial outcomes, whereas for mothers, it tends to be associated with worse outcomes (but with small effect sizes). The GRS for alcohol consumption did not follow this trend, lending support for the apparent protective effect of partner alcohol consumption being explained by unmeasured or residual confounding. The results for early onset smoking (<12y) in relation to blood pressure are not discussed in this paper, but the large differences between parents-of-interest are intriguing and warrant further investigation via EPoCH Explorer and in further research.

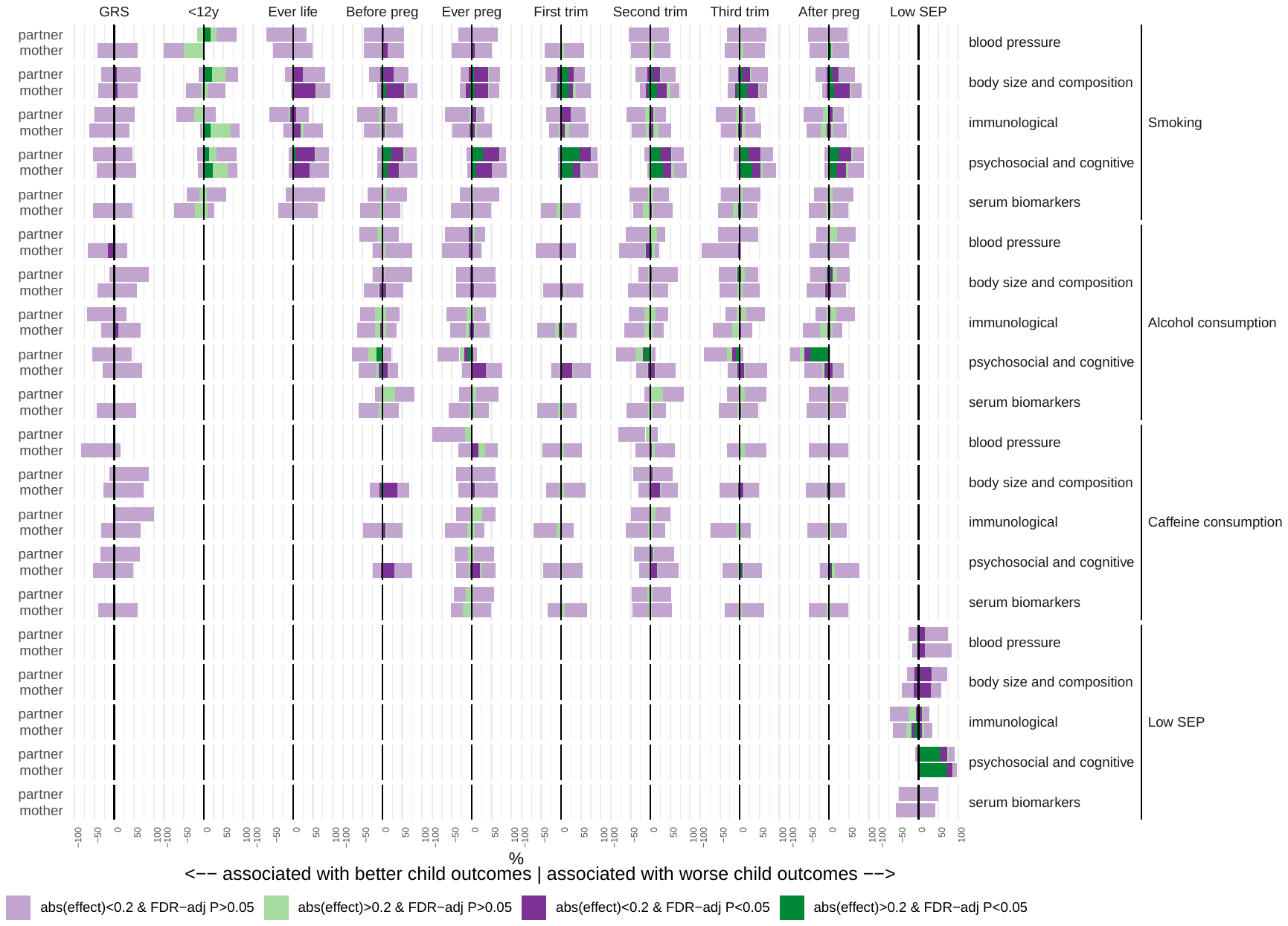

***Figure 2.*** *Results for model 2a broken down by exposure class, exposure time, and outcome class.*

We broke down the distributions in different ways to help visualise these and other trends more clearly. **Figure 3** highlights that there is no clear evidence that exposures during pregnancy (compared to post or prenatal) or in specific trimesters (compared to any time in pregnancy) yielded stronger evidence of association; i.e., no strong evidence that the timing of exposure is important. Similarly, **Figure 4** shows no clear pattern in the of evidence according to child’s age at outcome; i.e. no evidence that associations are stronger for outcomes that occur closer to pregnancy.

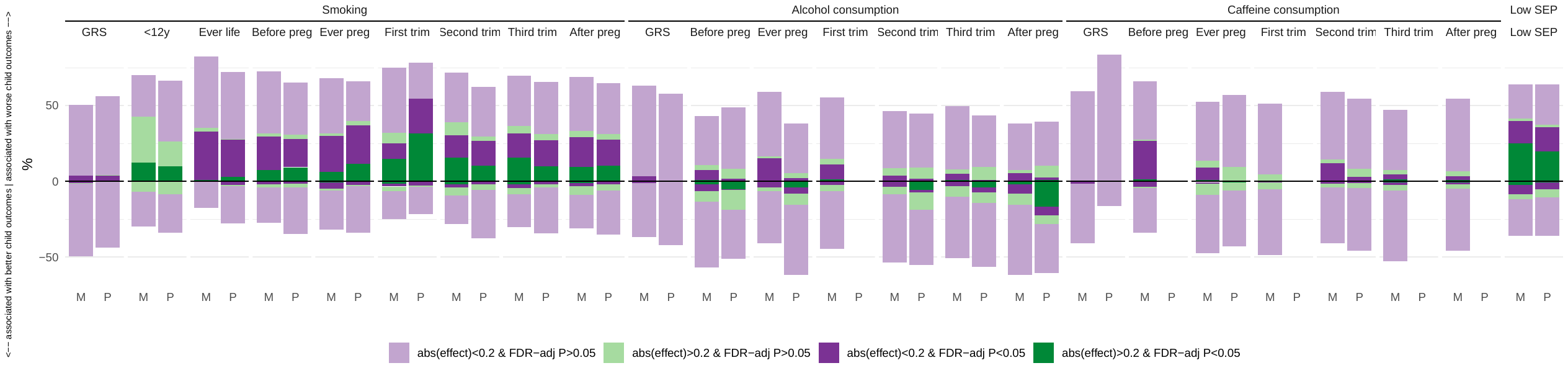

***Figure 3.*** *Results for model 2a broken down by exposure class and time. X axis key: M=mother; P=partner*

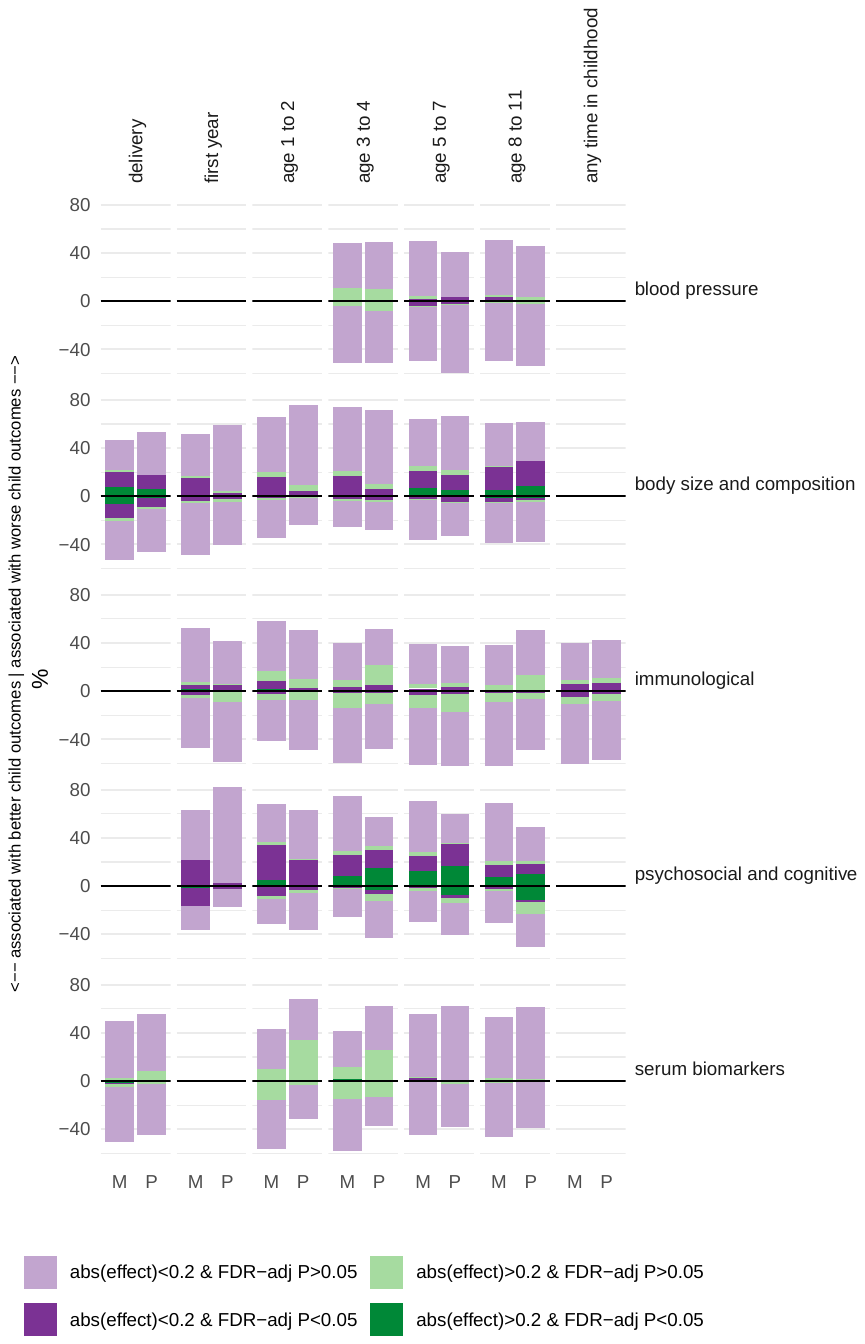

***Figure 4.*** *Results for model 2a broken down by outcome class and time. X axis key: M=mother; P=partner*

Finally, we broke down the distributions by model and exposure class (**Figure 5**), which shows, as would be expected, smaller effect estimates and larger P-values with increased level of adjustment.

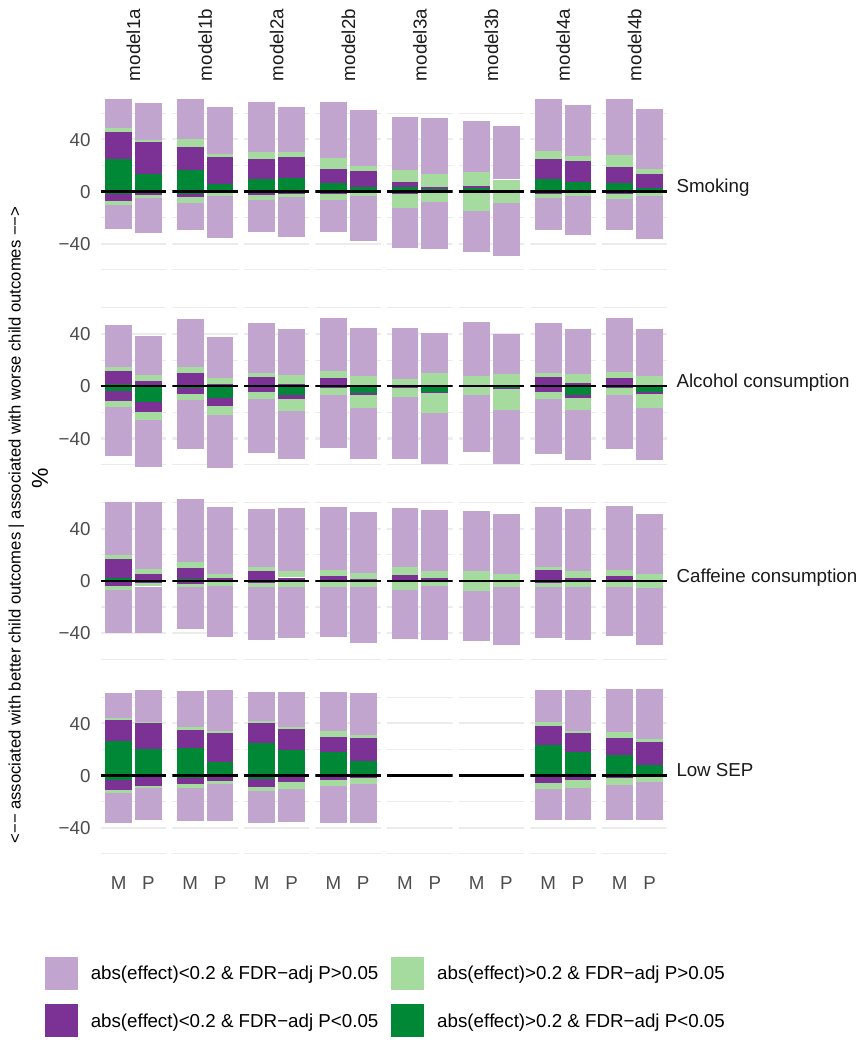

***Figure 5****. All results broken down by model and exposure class. X axis key: M=mother; P=partner*

Positive associations between partner alcohol consumption and psychosocial outcomes

**Figure 6** shows that positive associations exist between partner alcohol and some psychosocial outcomes. This contrasts with what we see for mothers, where higher levels of alcohol are associated with worse outcomes. All associations with partners’ GRS for alcohol are of very small magnitude and null (except for internalising traits, which is still very small but with a 95% CI that does not cross the null), providing some confidence that these positive associations are likely explained by residual confounding (by SEP). Although it should be noted that the F-statistic for the association between the partner GRS and alcohol consumption in pregnancy was 10 (reasonably weak strength instrument).

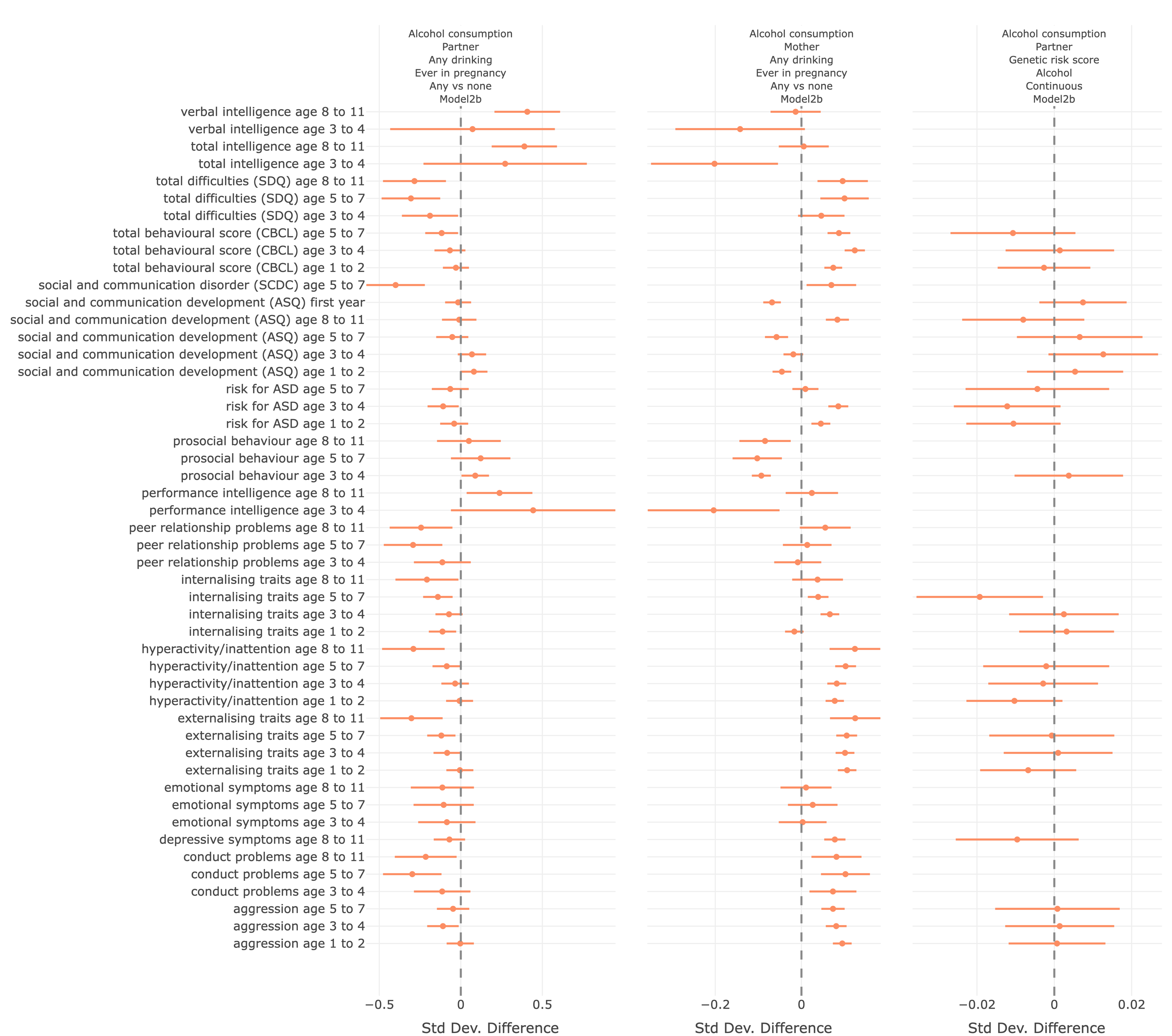

***Figure 6.*** *Coefficient plots (from EPoCH Explorer) comparing results for associations between partner alcohol during pregnancy (left), maternal alcohol during pregnancy (middle), and the partner GRS score for alcohol (right) on child psychosocial/cognitive outcomes. Note the X-axis scales are different.*
