## Supplementary File S7: Example from EPoCH Explorer for "Exploring parental prenatal influences on child health: a multicohort study and data visualisation tool"

**EPoCH Explorer description**

EPoCH Explorer is a web app built in RShiny that enables users to explore and download the results (summary statistics) of the EPoCH study. It is available at [*https://gcsharp.shinyapps.io/EPoCH/*](https://gcsharp.shinyapps.io/EPoCH/) and the code is available at <https://github.com/gcsharp/EPoCH_results_app>. Figure 1 shows the opening screen.

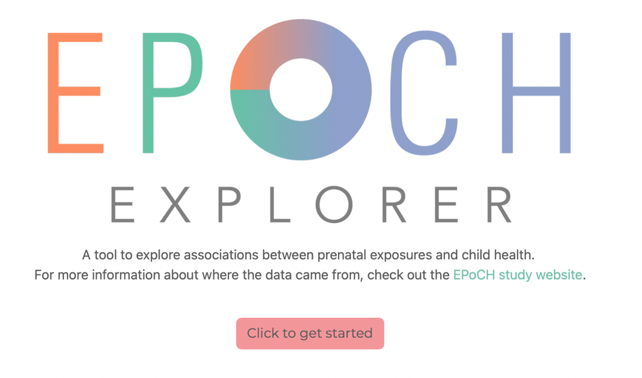

***Figure 1.*** *The opening screen of EPoCH Explorer*

On selecting ‘Click to get started’ the data is loaded, and a modal appears to explain that this will take a few seconds. The main screen of the app then appears with the navigation bar (Figure 2) at the top. The user can click the tabs to gain insights into the EPoCH results.

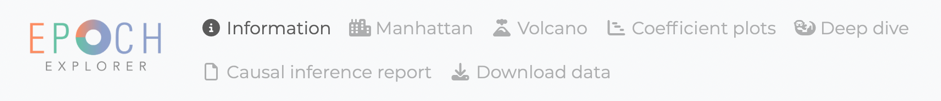

***Figure 2.*** *EPoCH Explorer navigation bar*

Welcome/About

A description of each tab is provided on the welcome screen (Figure 3).

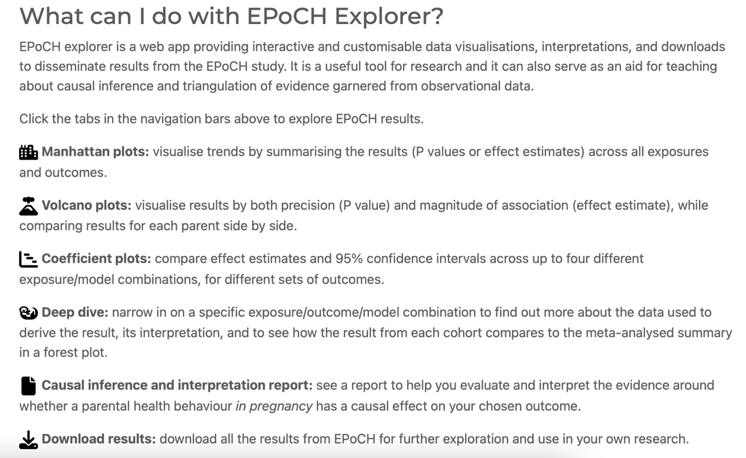

***Figure 3.*** *Information about each tab provided on the welcome screen*

On sub-tabs within the welcome/about page, users can view basic information about each cohort that contributed to EPoCH, followed by an interactive table describing the distribution of variables (the same tables that are used in Supplementary File 6 of this paper). An example for ALSPAC is shown in Figure 4.

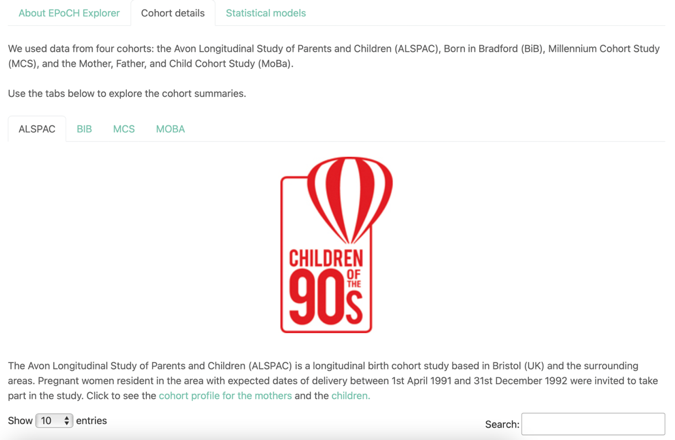

***Figure 4.*** *Users can view basic information about each cohort, followed by a detailed summary of the distribution of variables in an interactive table.*

The statistical models are also described on a separate sub-tab.

Manhattan plots

Manhattan plots (**Figure 5**) allow the user to visualise trends by summarising the results (P values or effect estimates) across all exposures and outcomes. They can select the data to plot using the dropdown boxes and, once generated, they can filter results by parent-of-interest and select whether to view -log10 P-values or standardised effect estimates (Cohen’s D) on the Y axis, and exposures or outcome on X. All plots (created using Plotly) are interactive, so the user can zoom in on certain areas. Hovering the cursor over a point will reveal pertinent information about the association (e.g. exposure, outcome, effect estimate, p-value, sample size, contributing cohorts).

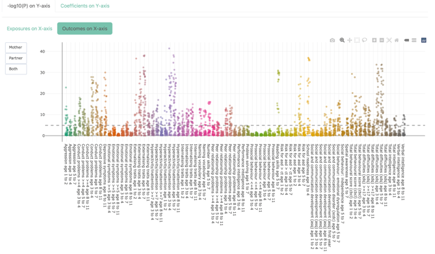

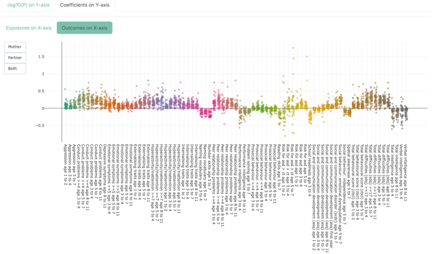

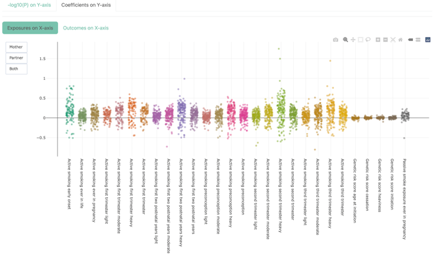

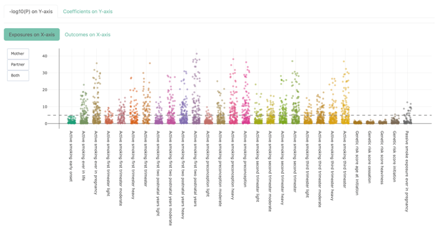

***Figure 5.*** *Example Manhattan plots showing the same set of results (smoking, psychosocial/cognitive outcomes, standard unstratified model without adjustment for the co-parent’s exposure, i.e. model 2a). Top left: Y=-log10P, X=outcomes; top right: Y=effect estimates, X=outcomes; bottom left: Y=-log10P, X=exposures; bottom right: Y=effect estimates, X=exposures.*

Volcano plots

Volcano plots (**Figure 6**) allow the user to visualise results by both precision (-log10 P) and magnitude of association (standardised effect estimate Cohen’s D). Plots for each parent-of-interest are plotted side-by-side. Users can select the data to plots using the dropdown boxes and, once generated, they can select whether the Y axes display the raw -log10 P-values or the ranked versions. Raw values allow users to visualise differences between parents-of-interest more clearly, whereas ranked values spread the data out along the Y-axis more equally, allowing easier identification of points with larger P-values. As with the Manhattan plots, the user can hover over a point to reveal more information (as shown in **Figure 6**).

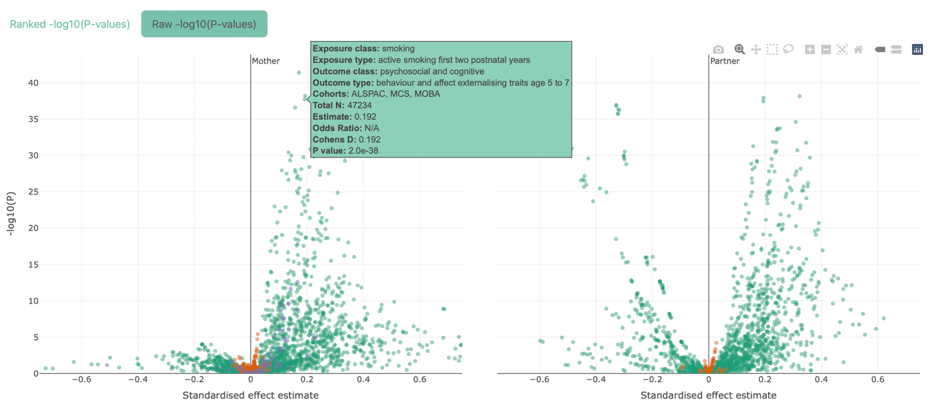

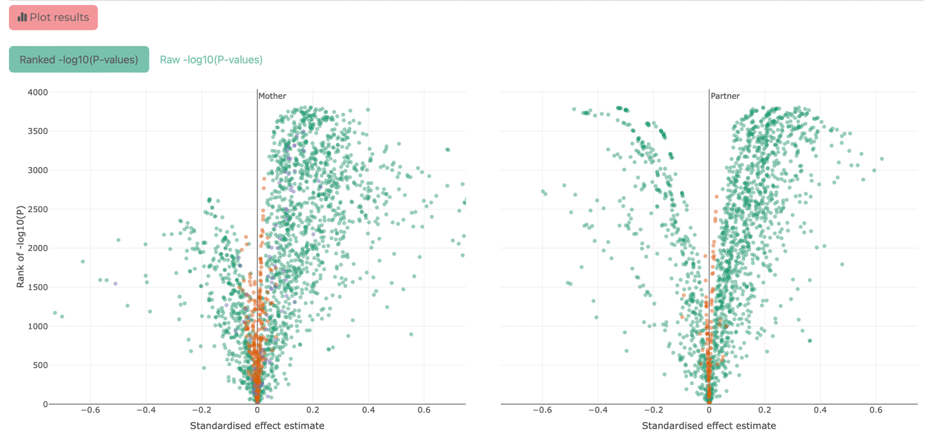

***Figure 6.*** *Example Volcano plots showing the same set of results (smoking, psychosocial/cognitive outcomes, standard unstratified model without adjustment for the co-parent’s exposure, i.e. model 2a). Top: Y= raw -log10P, mother on left, partner on right; bottom: Y= ranked -log10P, mother on left, partner on right. The top panel shows an example of the information displayed on hovering over a point.*

Coefficient plots

Coefficient plots (**Figure 7**) allow the user to compare effect estimates and 95% confidence intervals from up to four different exposure/model combinations side-by-side. The user selects the outcome class and type (i.e. binary or continuous) to view.

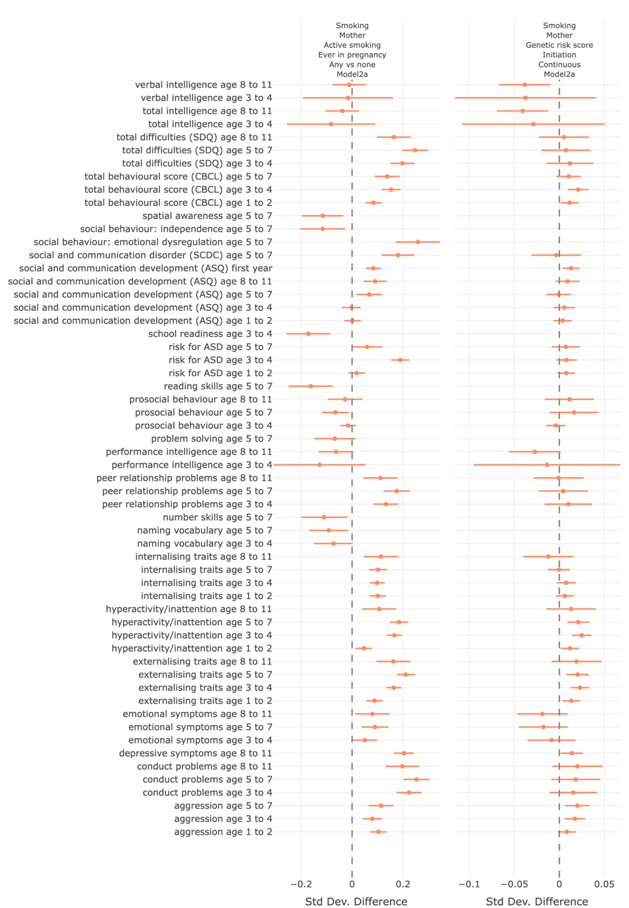

***Figure 7.*** *Example Coefficient plots comparing the observational results (i.e. self-reported maternal smoking ever in pregnancy) vs results from the genetic risk score for smoking initiation.*

Deep Dive

On the ‘Deep Dive’ tab, users can select a specific association (i.e. select the exposure subclass and timing, the outcome subclass and timing, and the model) and generate a detailed description of the result, including an interactive forest plot displaying the effect estimates and 95% CIs for each cohort along with the meta-analysis result (**Figure 8**). The verbal interpretation summarises the same size, number and % exposed, effect estimate and 95% CIs, number of contributing cohorts, and (where n cohorts >1) the heterogeneity statistics I2 and heterogeneity P-value. These are integrated with a verbal summary of the results that attempts to translate the results into terms that can be more easily understood by students or other interested non-experts.

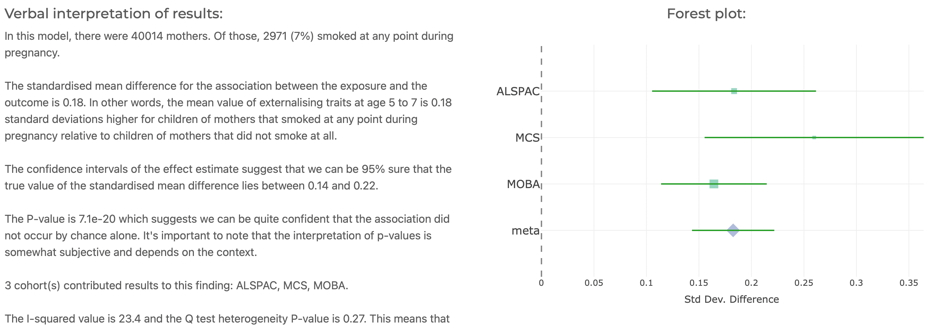

***Figure 8.*** *Example extract of a result from a ‘Deep Dive’*

Causal inference report

This tab allows users to select a health behaviour and parent of interest, and a specific outcome, and generate a series of plots designed to help them evaluate and interpret the evidence around whether their exposure *in pregnancy* has a causal effect on their outcome. An example of a causal inference report is provided in the next section of this file.

Download

The user can download all results by first agreeing to three conditions (around appropriately interpreting and citing our work), and then clicking the download button that will appear (**Figure 9**).

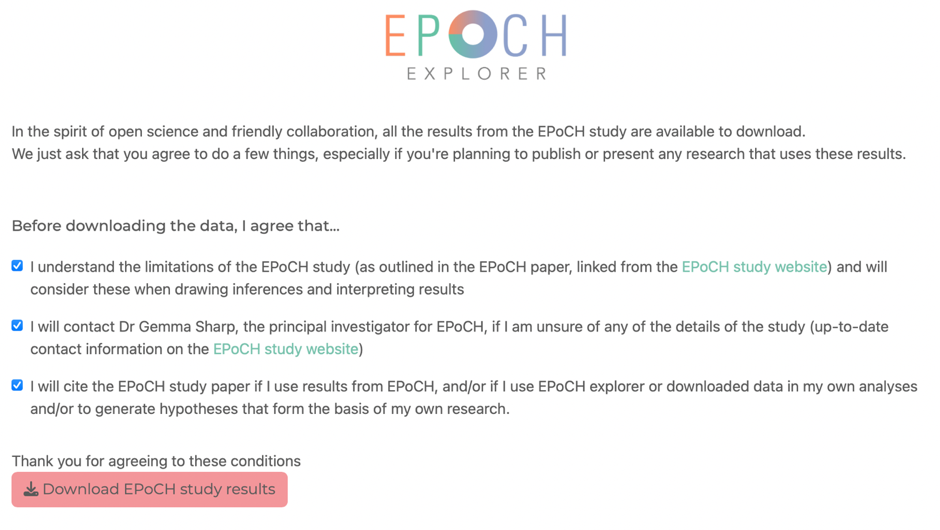

***Figure 9.*** *The download tab*

**Example of a causal inference report generated in EPoCH Explorer**

The following screenshots show the various sections of a causal inference report generated to evaluate evidence for an effect of maternal smoking in pregnancy on externalising traits at age 5 to 7.

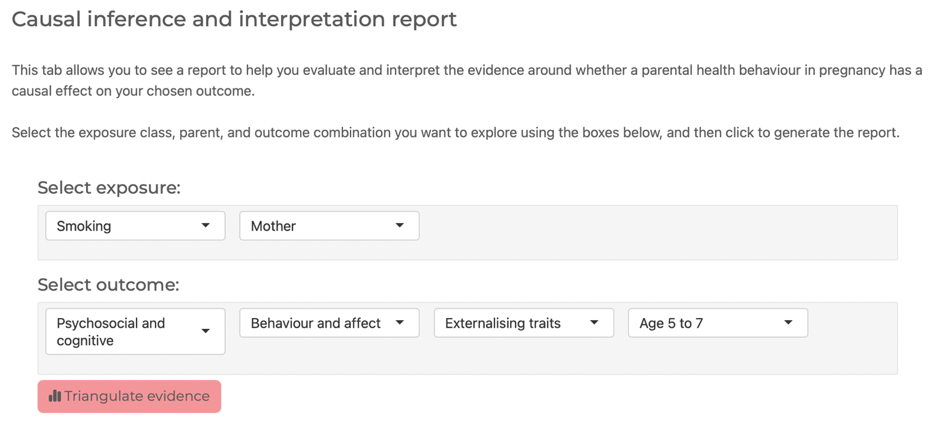

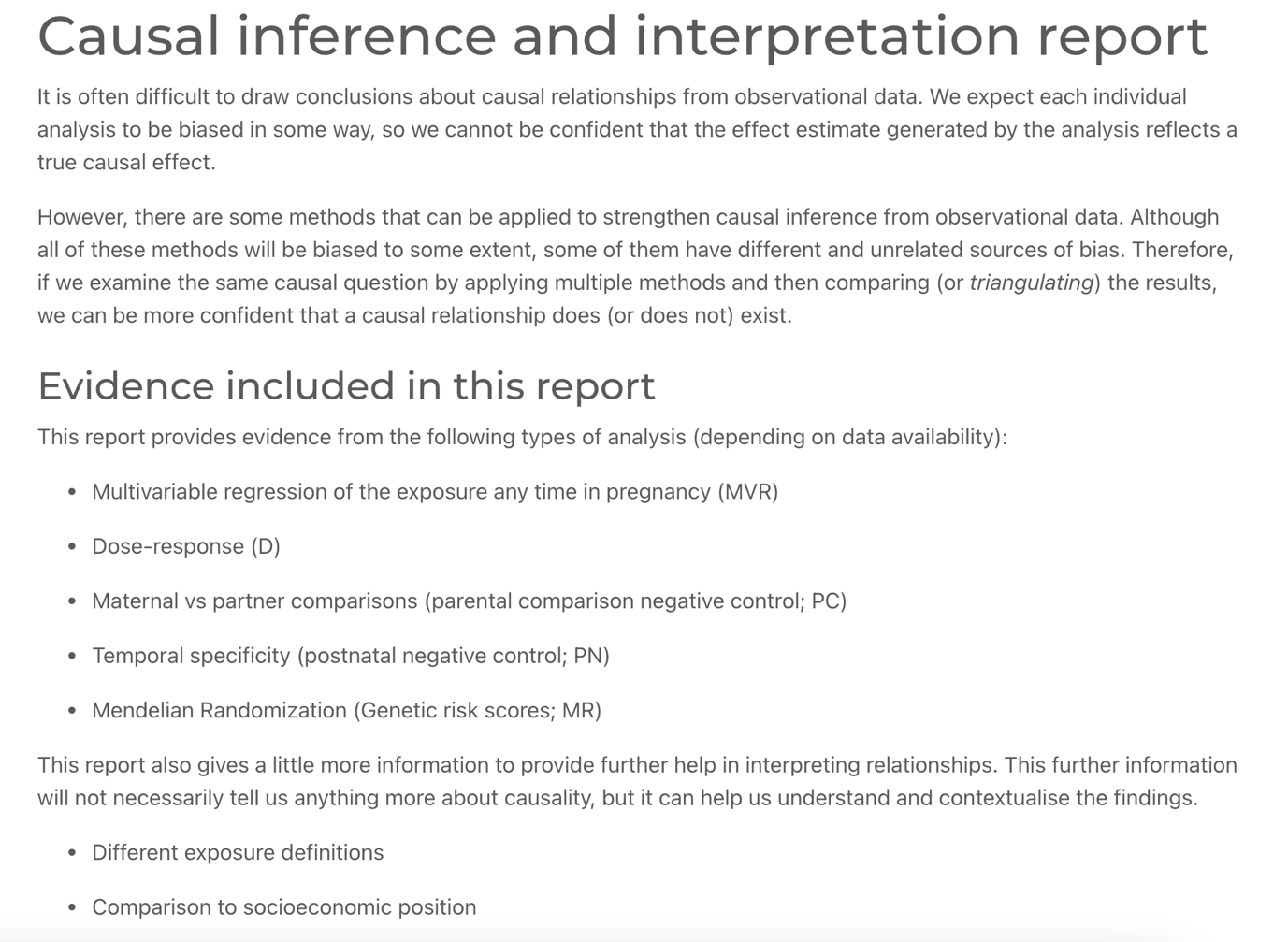

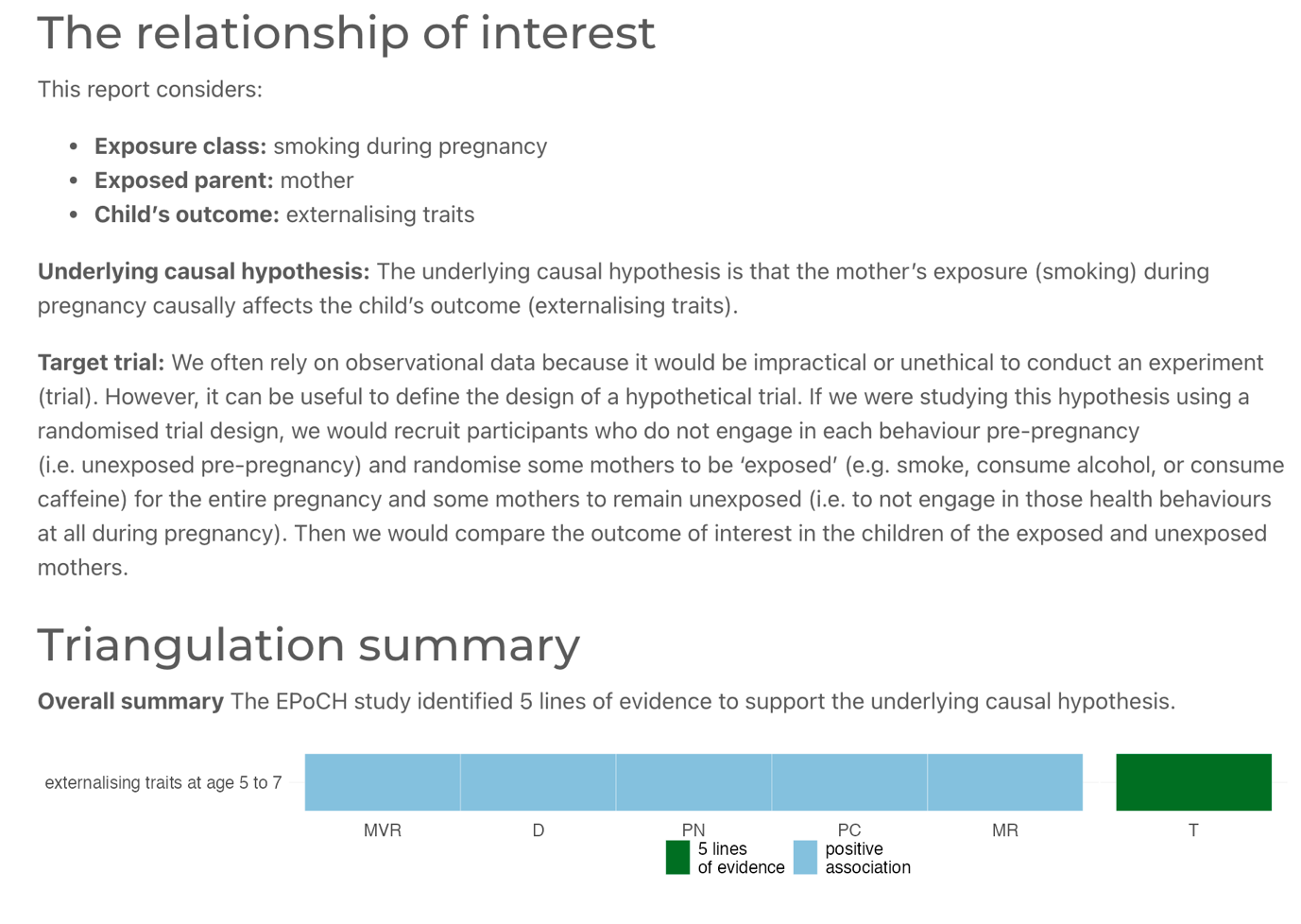

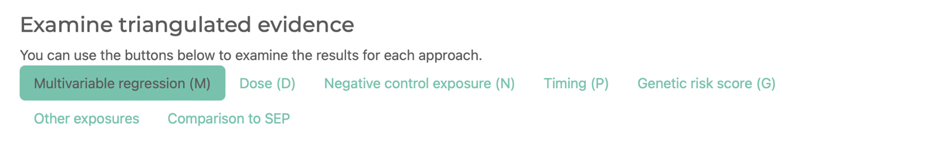

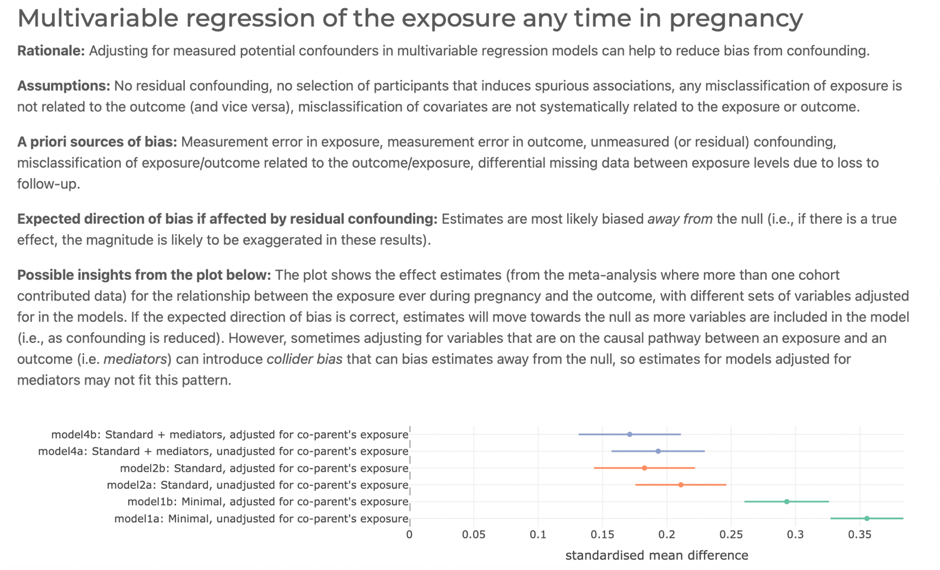

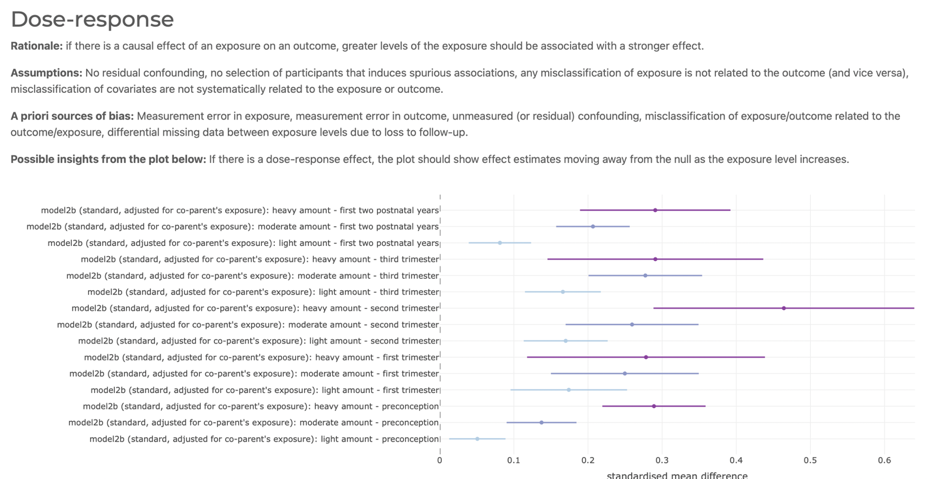

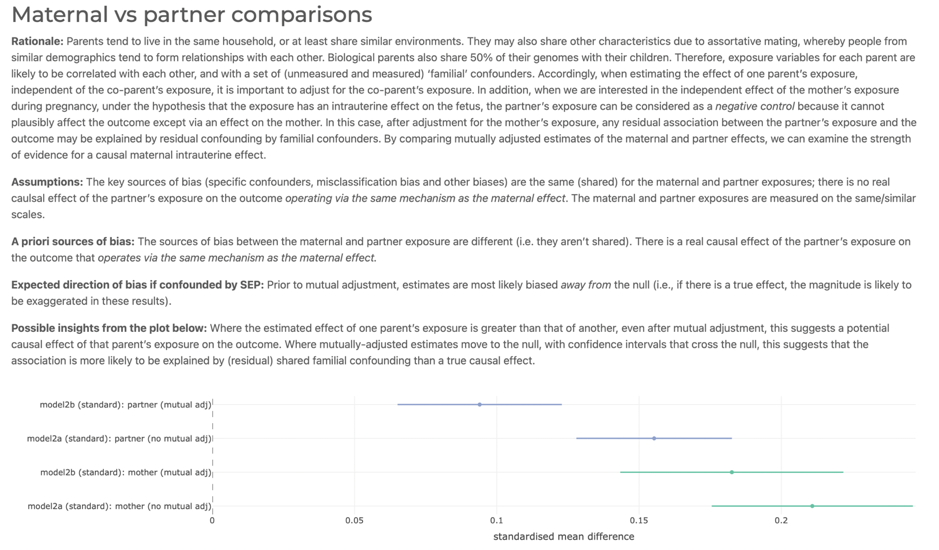

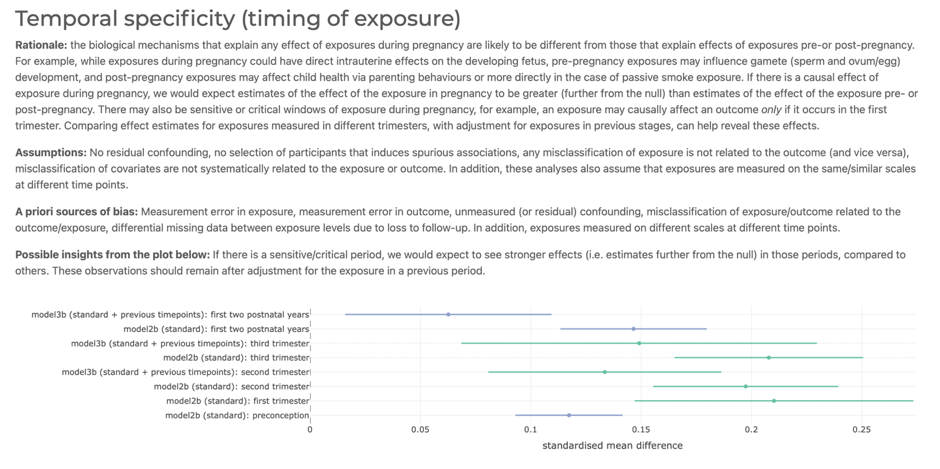

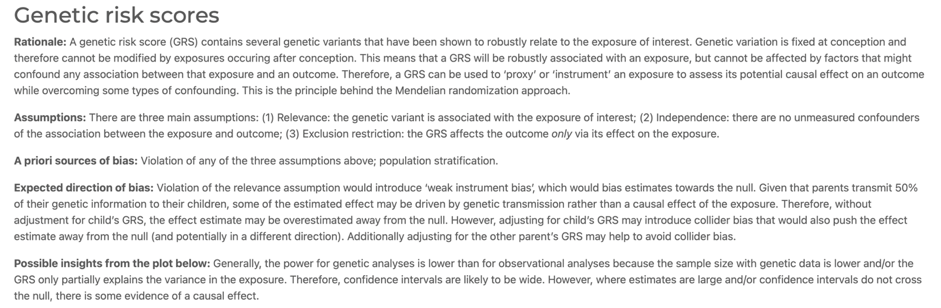

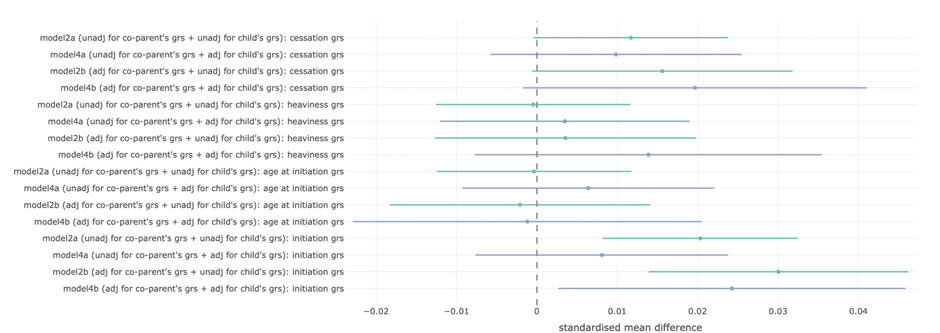

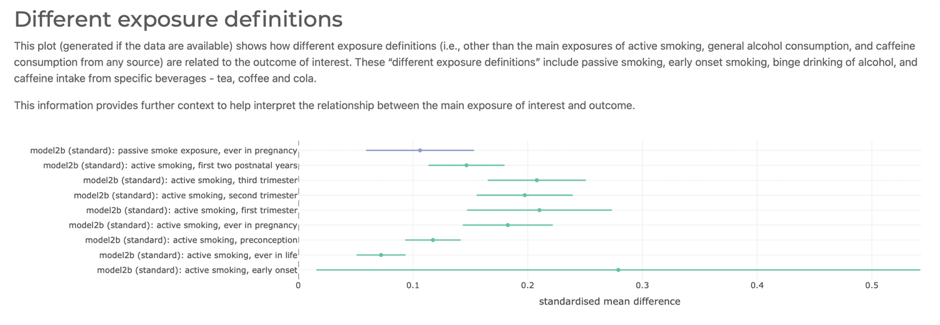

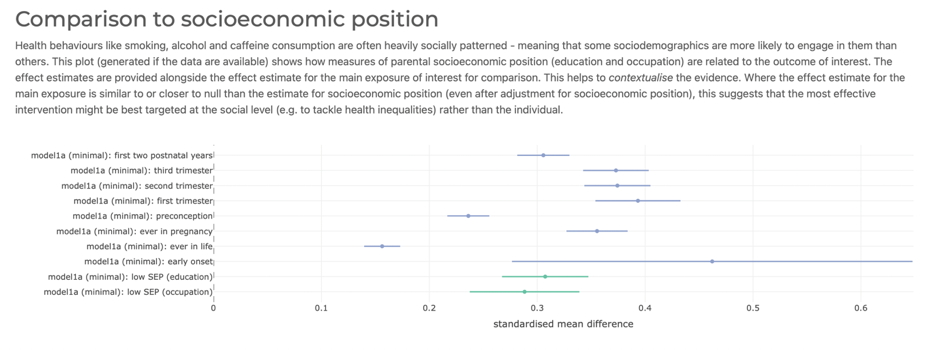
